# Measuring and increasing rates of self-isolation in the context of infectious diseases: A systematic review with narrative synthesis

**DOI:** 10.1101/2023.09.29.23296339

**Authors:** Louise E Smith, Alex F Martin, Samantha K Brooks, Rachel Davies, Madeline V Stein, Richard Amlôt, Theresa M Marteau, G James Rubin

**Affiliations:** King’s College London, Institute of Psychiatry, Psychology and Neuroscience; NIHR Health Protection Research Unit in Emergency Preparedness and Response; UK Health Security Agency, Behavioural Science and Insights Unit; Department of Psychology, King’s College London; University of Cambridge, Department of Public Health and Primary Care

**Keywords:** COVID-19, isolation, quarantine, definitions, measures, adherence, factors associated

## Abstract

**Background:** Self-isolation was used to prevent the spread of COVID-19 and will likely be used in future infectious disease outbreaks.

**Method:** We conducted a systematic review following PRISMA and SWiM guidelines. MEDLINE, PsycINFO, Embase, Web of Science, PsyArXiv, medRxiv, and grey literature sources were searched (1 January 2020 to 13 December 2022) using terms related to COVID-19, isolation, and adherence. Studies were included if they contained original, quantitative data of self-isolation adherence during the COVID-19 pandemic. We extracted definitions of self-isolation, measures used to quantify adherence, adherence rates, and factors associated with adherence. The review was registered on PROSPERO (CRD42022377820).

**Findings:** We included 45 studies. Self-isolation was inconsistently defined. Only four studies did not use self-report to measure adherence. Of 41 studies using self-report measures, only one reported reliability; another gave indirect evidence for a lack of validity of the measure. Rates of adherence to self-isolation ranged from 0% to 100%. There was little evidence that self-isolation adherence was associated with socio-demographic or psychological factors.

**Interpretation:** There was no consensus in defining, operationalising, or measuring self-isolation. Only one study presented evidence of the psychometric properties of the measure highlighting the significant risk of bias in included studies. This, and the dearth of scientifically rigorous studies evaluating the effectiveness of interventions to increase self-isolation adherence, is a fundamental gap in the literature.

**Funding:** This study was funded by Research England Policy Support Fund 2022-23; authors were supported by the NIHR Health Protection Research Unit in Emergency Preparedness and Response.

## Introduction

Isolation is the separation of those who are ill from those who are well, while quarantine is the separation of those at risk of developing an illness from those who are well. [1] During the COVID-19 pandemic, many countries implemented isolation and quarantine orders which required people to stay at home (or in supported isolation such as a hotel) and not leave except for very few reasons (e.g., to take or return a COVID-19 test). These differed to population-wide “stay-at-home” or “lockdown” orders (also known as “mass quarantine”), where people were required to stay at home, but could leave to buy essentials (groceries / pharmacy) and for exercise. In this paper we refer to both isolation and quarantine as “self-isolation”.

Self-isolation of suspected COVID-19 cases and their household members could substantially reduce transmission. [2] However, self-isolation only succeeds when people adhere to it. A rapid review of adherence to self-isolation in 14 studies (all conducted before the COVID-19 pandemic) found rates of 0% to 93% adherence. [3] Low adherence may be partly explained by self-isolation’s significant financial, psychological and practical implications. [3] The COVID-19 pandemic caused researchers, public health practitioners, and policy-makers to focus on self-isolation as a prevention measure for infection, [4] revealing several uncertainties. [5]

First, it remains unclear how adherence should be defined and operationalised. Should the denominator be everyone in the population with relevant symptoms who has not tested negative, or only the subset who are in contact with public health teams (e.g., who have a received a positive test)? Similarly, which forms of non-adherence should we be concerned about? For research and surveillance purposes, it may be easiest to have a clear-cut definition in which any infraction of the guidance counts as non-adherence. However, qualitative research suggests that while people may break self-isolation rules, they often do so in ways that do not pose a substantial transmission risk. [6] Definitions of non-adherence need to be clear as to whether they are measuring complete adherence to guidelines, or avoidance of behaviours that pose a more than trivial risk of transmission.

Second, adherence can be measured in different ways. During the pandemic, online surveys were a common way to collect data quickly, cheaply, and safely, despite limitations in sampling strategies and representativeness of participants. [7] Self-report measures are subject to response bias, and rates of self-reported and observed behaviour often differ. [8] Other measures of self-isolation included geofencing, [9] in-person spot checks, [10] and assessment of contact tracing data for contacts who later became cases. Each has advantages and disadvantages in terms of cost, acceptability, reliability, and validity.

Third, improving adherence to self-isolation while also reducing its burden on individuals and society is a scientific and policy priority. [11] Research into how best to do this depends upon reducing the uncertainties described above.

This systematic review describes and appraises 1) definitions of self-isolation used in studies during the COVID-19 pandemic, 2) measures used to quantify adherence and their reliability, validity, and acceptability, 3) rates of self-isolation adherence, and 4) factors associated with adherence.

## Methods

We conducted a systematic review in accordance with the Preferred Reporting Items for Systematic Reviews and Meta-Analyses (PRISMA) checklist (see Supplement S1) and guidance by Cochrane. [12] A protocol was registered on PROSPERO (CRD42022377820). Deviations from the protocol are outlined in Supplement S2.

### Search strategy and selection criteria

Databases (MEDLINE, PsycINFO, Embase, Web of Science, PsyArXiv, medRxiv) were searched from 1 January 2020 to 13 December 2022. The search strategy included terms for COVID-19, isolation and quarantine (combined with NOT social isolation), and adherence and compliance. The search was also used for another systematic review carried out in parallel, investigating the effect of self-isolation on wellbeing (full search terms in Supplement S3). [13] Screening for both reviews was conducted together until the full-text screening stage.

We conducted grey literature searches, including a) five grey literature databases, b) relevant UK Government public health agency and statistical agency pages, and other behavioural studies, c) Google searches, and d) making enquiries with relevant UK Government agencies (Supplement S3). References of included citations were also searched.

LES, AFM, SKB, RD, MVS, and GJR screened citations. To ensure consistency, all authors initially screened the same 300 citations, discussing queries and discrepancies until agreement on included studies was reached. Authors then independently screened citations, meeting weekly to discuss queries and reaching group agreement through discussion.

Studies were included in the review if they contained original, quantitative data of adherence to self-isolation during the COVID-19 pandemic. For this review, we defined self-isolation as anyone advised (either directly by a public health team, or via widely disseminated public health guidance) to avoid contact with others because they were known or suspected to have COVID-19 or because they were suspected to be incubating COVID-19. Studies that reported attitudes or intentions to self-isolate (vs completed behaviour), or adherence to other social mobility rules (e.g., lockdown, physical distancing) were excluded. Studies were excluded if more than 5% of the sample were reported to be isolating in hospital. No exclusions were made based on participant characteristics or language of publication. Where studies were not published in English, relevant full texts were translated. Where queries around inclusion were not solved by discussion, the corresponding author of the study was contacted for clarification. If no response was received, the study was excluded.

### Data analysis

Study characteristics (country, dates of data collection, study design, inclusion criteria, response rate), self-isolation characteristics (reason, duration, location), participant characteristics (gender, age), definition of self-isolation, adherence measure (reliability, validity, acceptability), rates of adherence, and factors associated with adherence were extracted by LES. Where studies reported different methods to measure adherence, we treated each as a separate study.

Risk of bias for rates of adherence to self-isolation was assessed using the Joanna Briggs Institute Prevalence Critical Appraisal Tool, [14, 15] a recommended tool for prevalence estimates. [16, 17] We made some amendments to the tool (Supplement S4). Associations with adherence to self-isolation were appraised using the Risk Of Bias In Non-randomized Studies - of Exposure (ROBINS-E) tool [18] or the Risk Of Bias In Non-randomized Studies - of Interventions (ROBINS-I) tool, depending on study design. [19] For each study, we assessed the most rigorous analysis conducted (e.g., multivariable regressions, over unadjusted regressions). Studies were rated as having low risk of bias (ROBINS-I “low”), some concerns (ROBINS-I “moderate”), high risk of bias (ROBINS-I “serious”), or very high risk of bias (ROBINS-I “critical”) based on the algorithms provided by the tools. LES completed risk of bias ratings.

Results were narratively synthesised for each of the review aims separately. Results are reported in line with synthesis without meta-analysis (SWiM) reporting guidelines. [20] A meta-analysis was not planned due to likely heterogeneity between studies (methods and materials) and definitions of self-isolation used. For brevity, details of analysis for each outcome are reported in Supplement S5.

### Role of the funding source

The funders of the study had no role in study design, data collection, data analysis, data interpretation, or writing of the report.

## Results

The search identified 15,277 reports (citations). Sixty-five reports were included; 35 from database and register searches and 30 from other searches (Figure 1). Two manuscripts each described two separate methods to measure adherence to self-isolation (Hood et al 2022 [21]: case interview, survey. Rubio et al 2021 [22]: survey, random home visits and calls). We treated these as separate studies. In total, reports described results from 45 studies.

**Figure 1.**
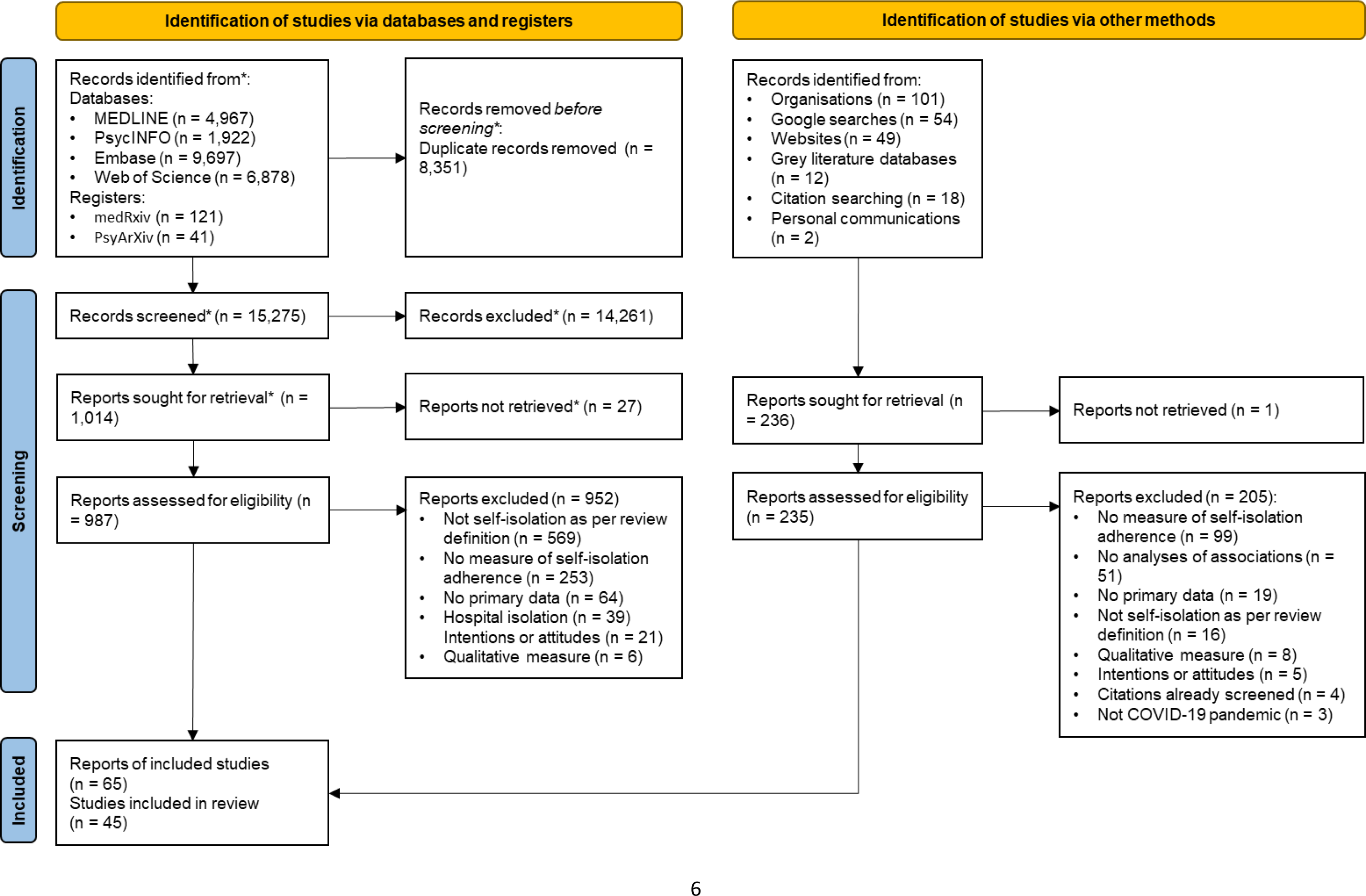
Flowchart of study selection. *At this stage, citation screening was completed for this systematic review and the systematic review investigating the effect of self-isolation on wellbeing. Therefore, these totals include citations screened for the systematic review investigating the effect of self-isolation on wellbeing as well as for this systematic review.

### Study characteristics

Thirty studies used a cross-sectional design, with a further four using a series of cross-sectional studies (Table 1). Seven studies used a longitudinal design, one was a non-randomised comparative design, one a case series, one a retrospective chart review, and one was secondary data analysis. In 18 studies participants were COVID-19 cases (or suspected cases), five studies investigated contacts of cases, six studies investigated cases and contacts, five studies investigated people with COVID-19-like symptoms, and three studies investigated people returning from travel (Table 1). Other studies used a combination of self-isolation reasons (Table 1). Seventeen studies were conducted in Europe, eight in the Middle East, seven in North America, three each in Africa, South Asia and East Asia, two in West Asia, and one each in Australasia and Southeast Asia (Table 1).

**Table 1.**
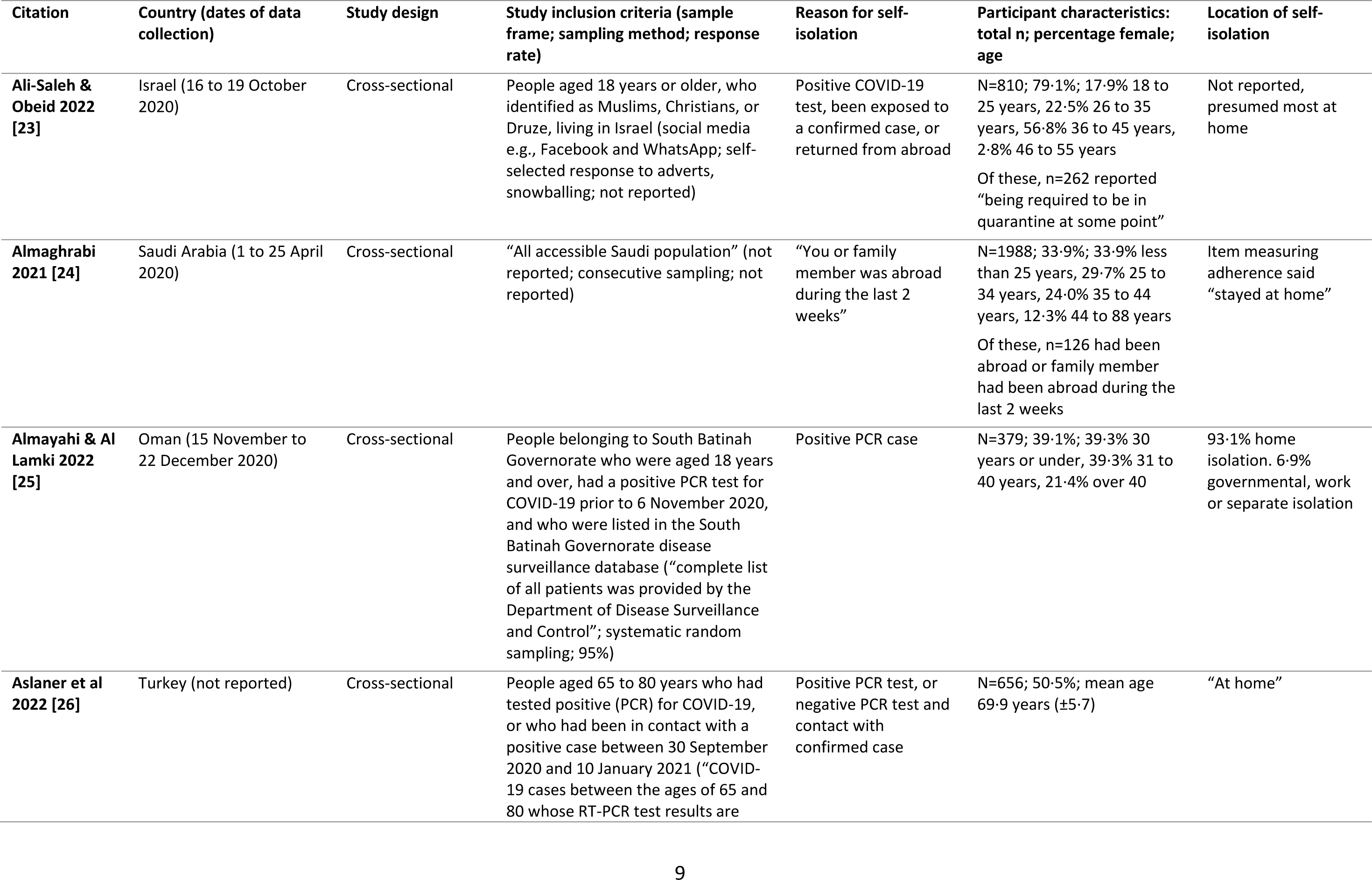
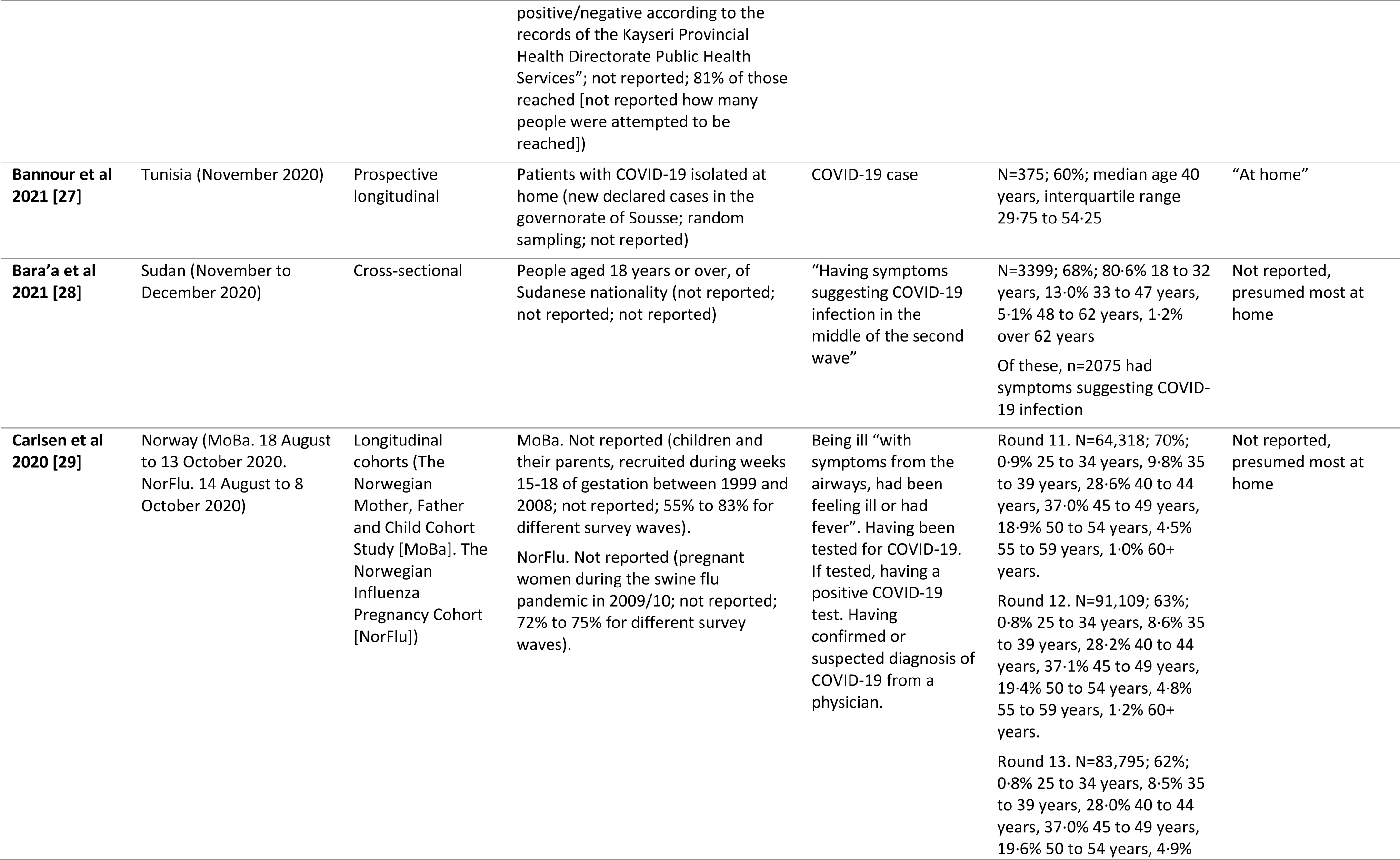
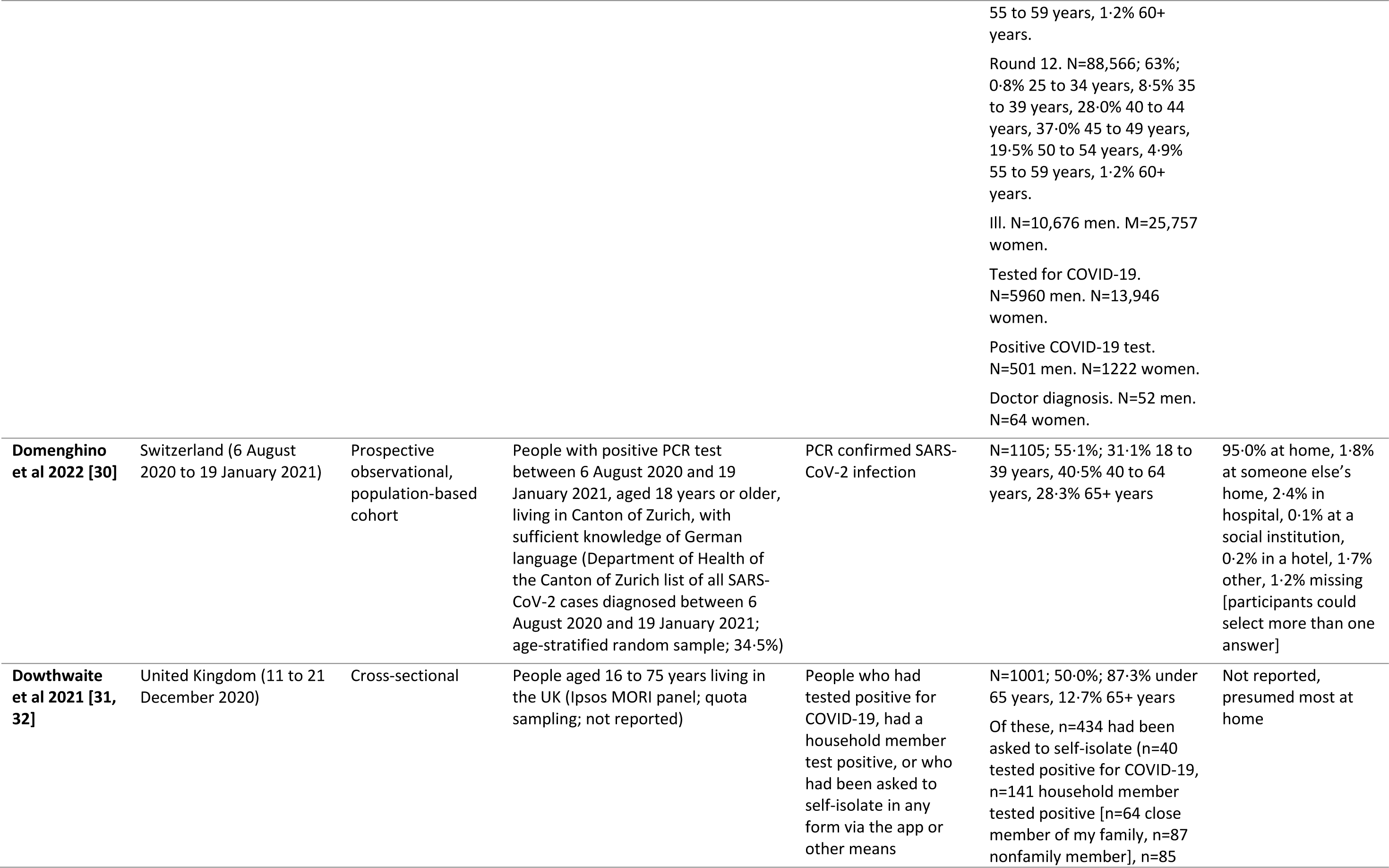
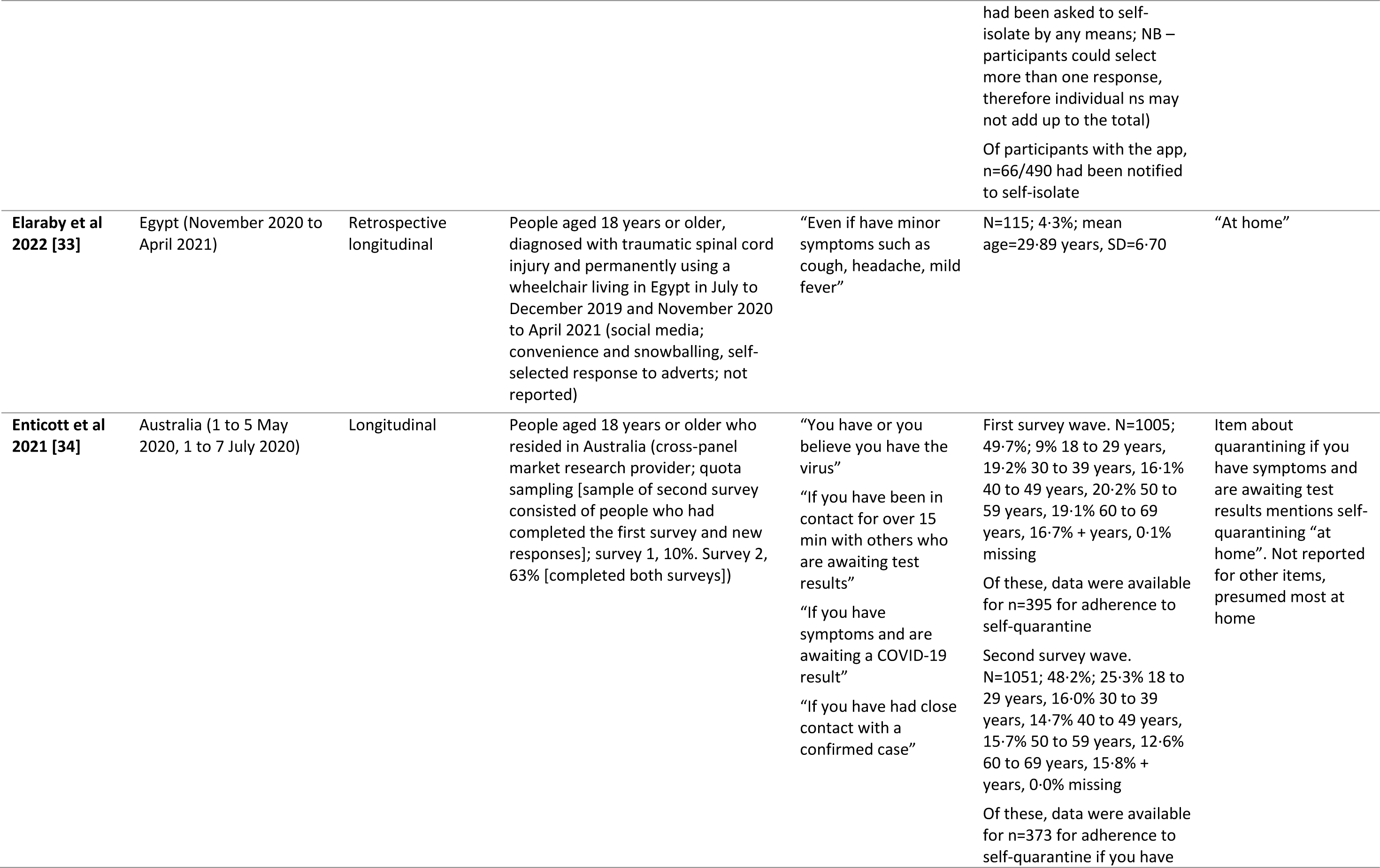
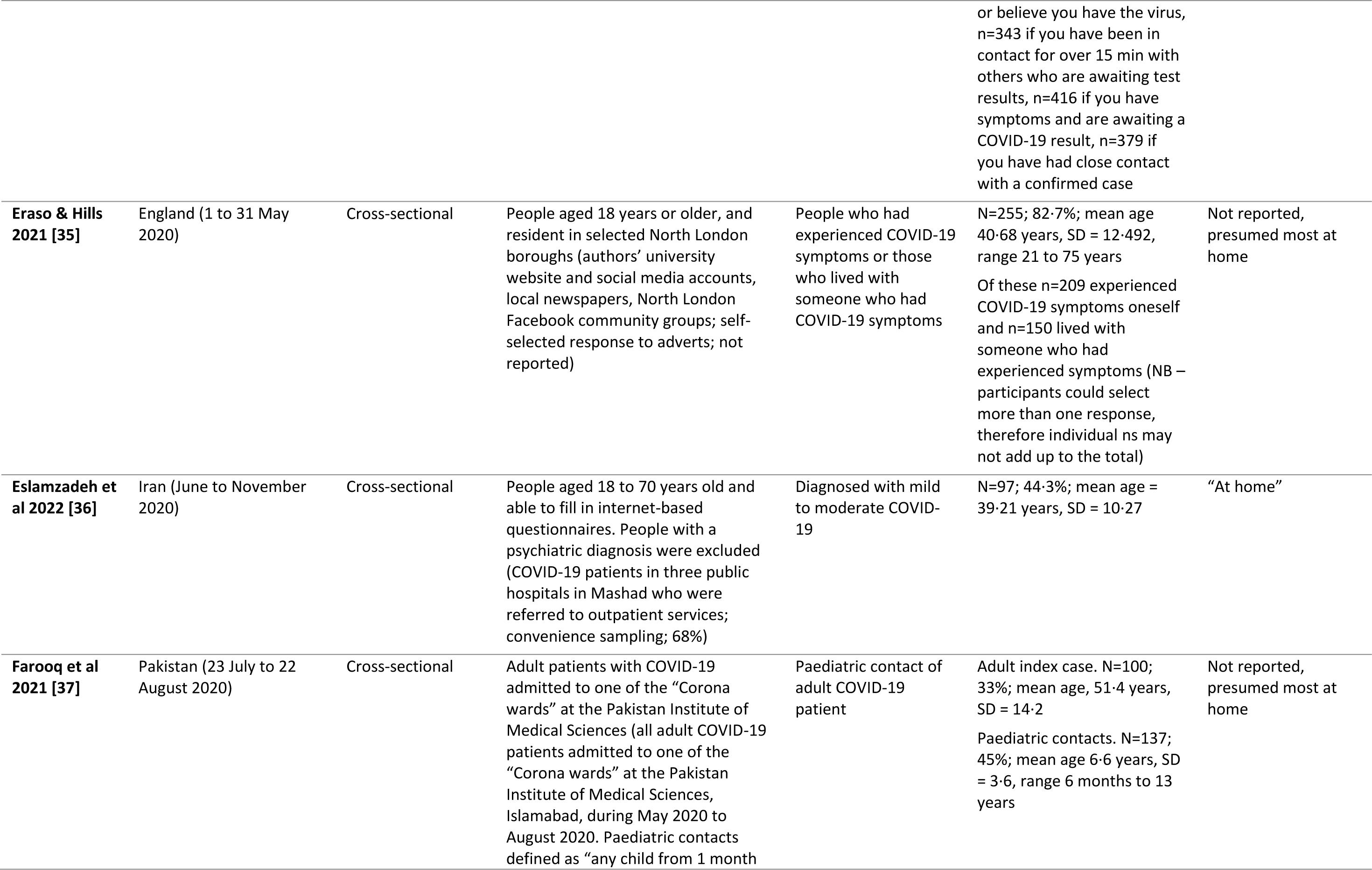
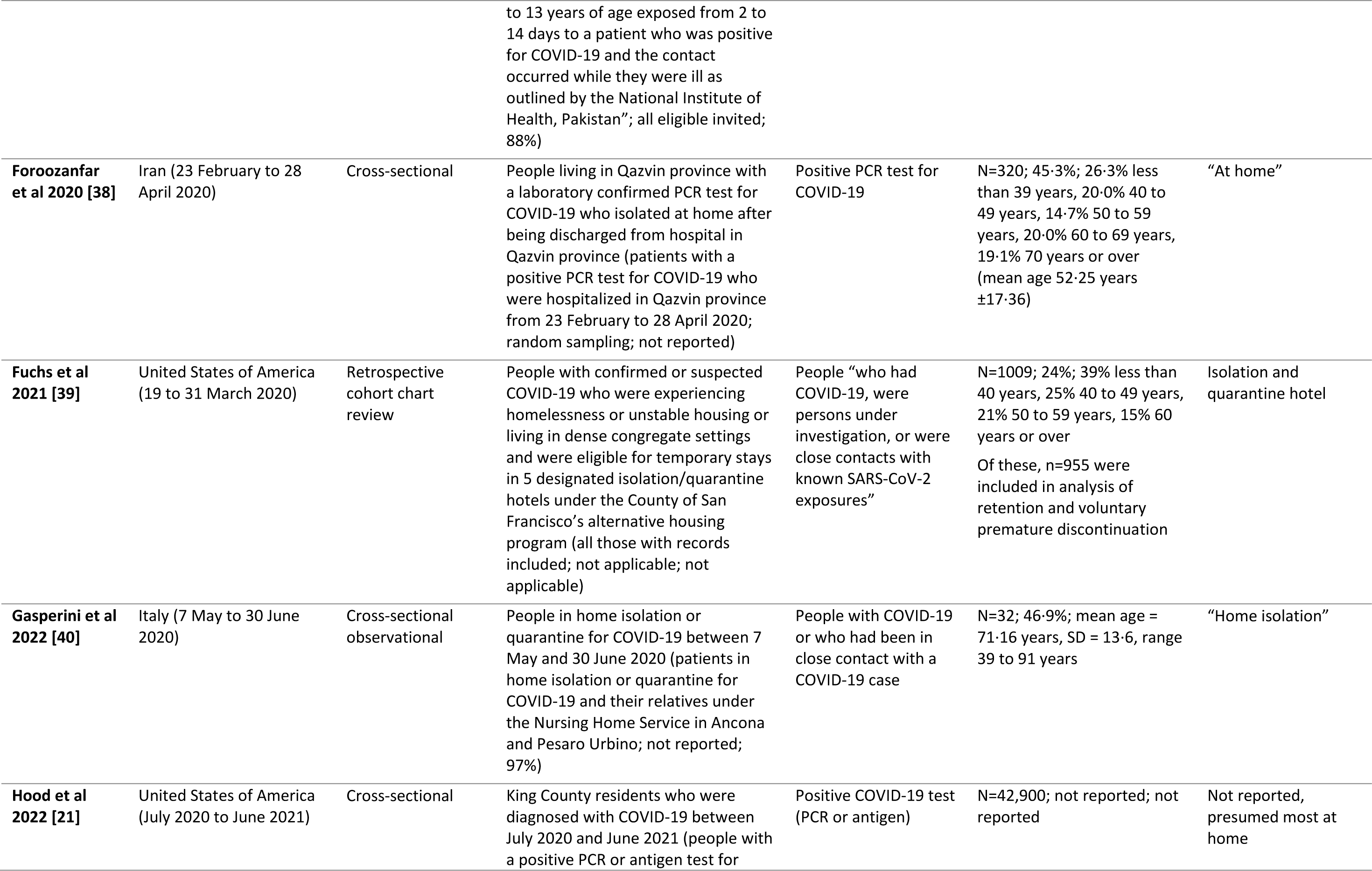
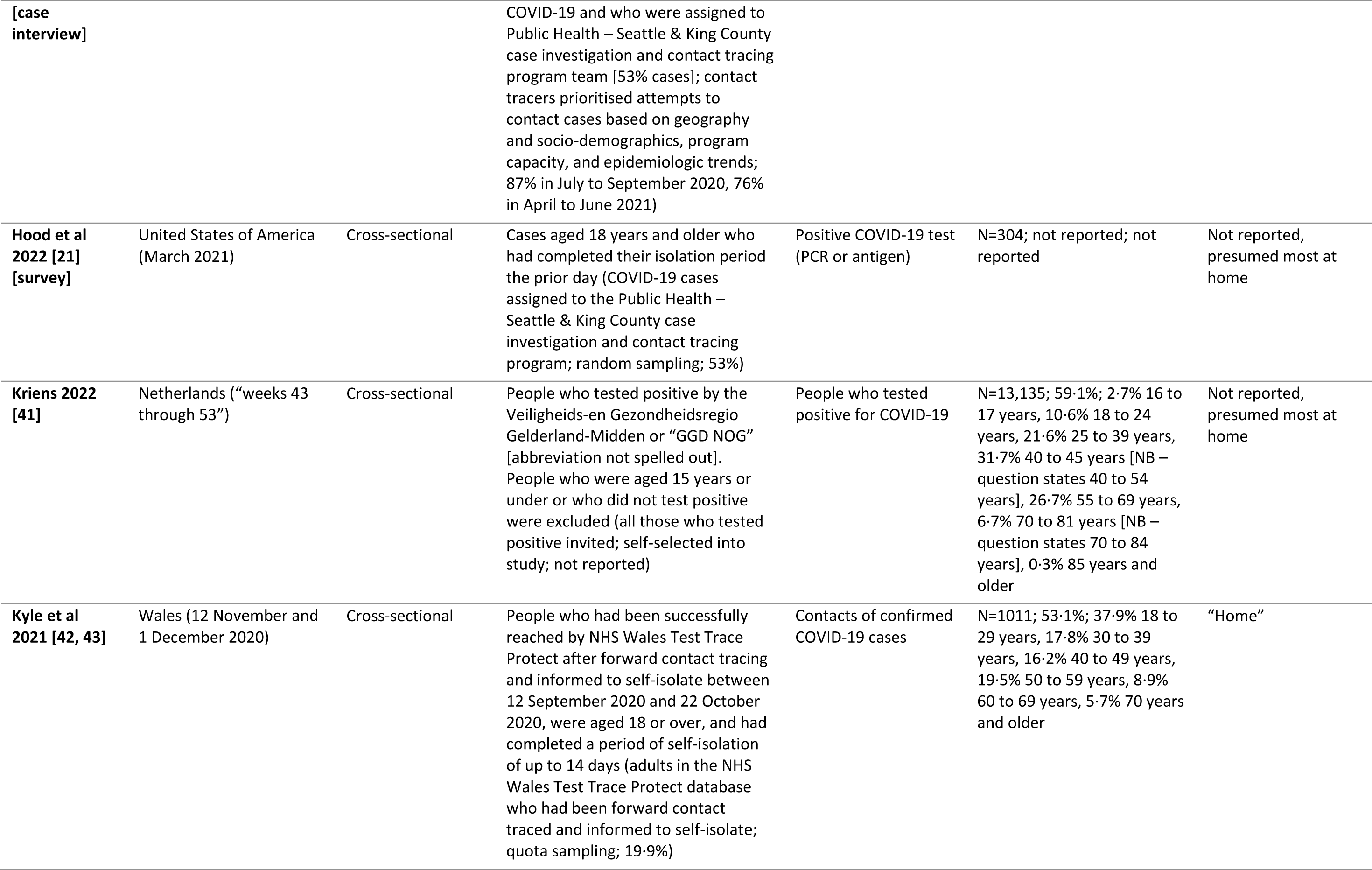
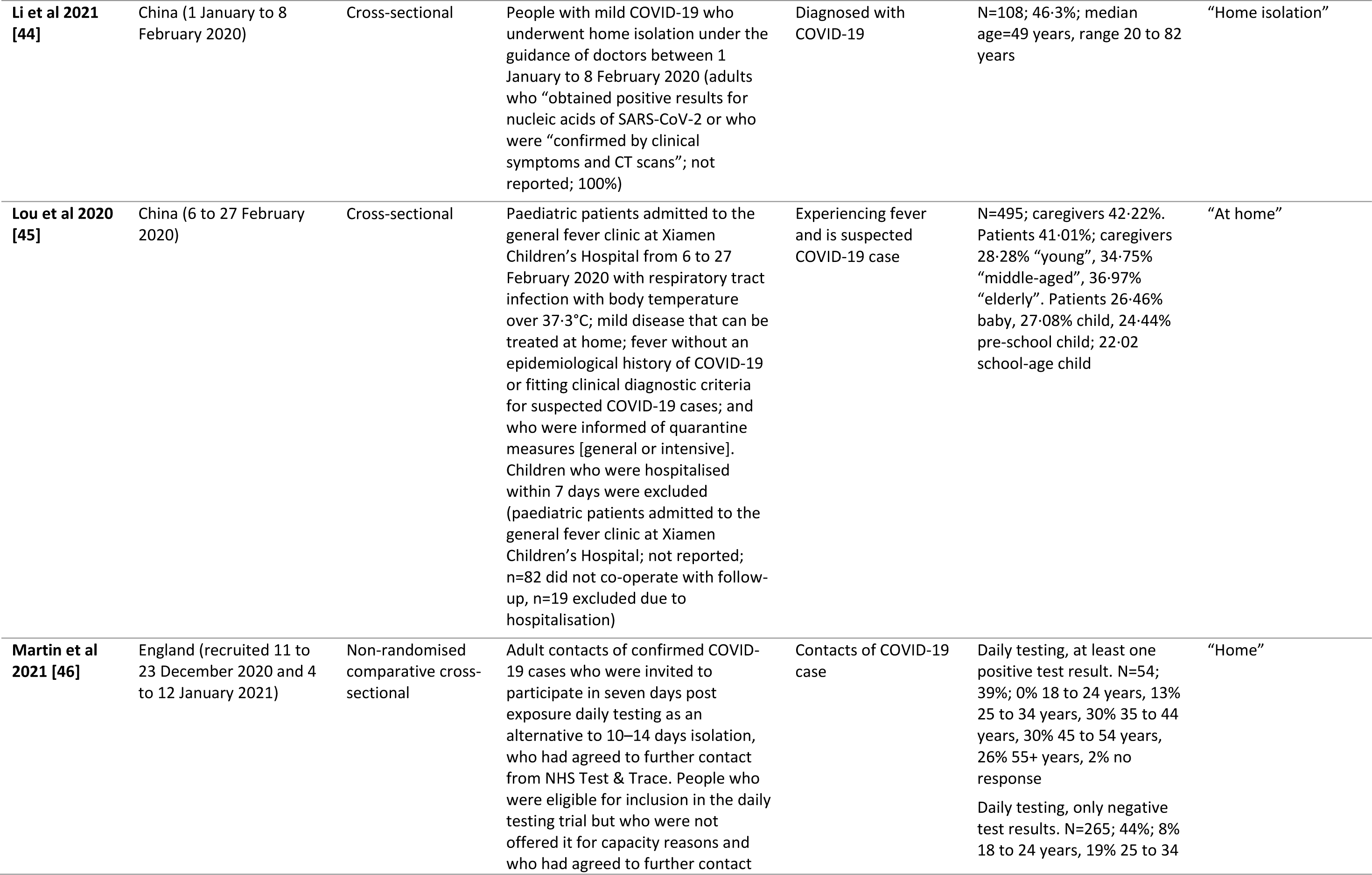
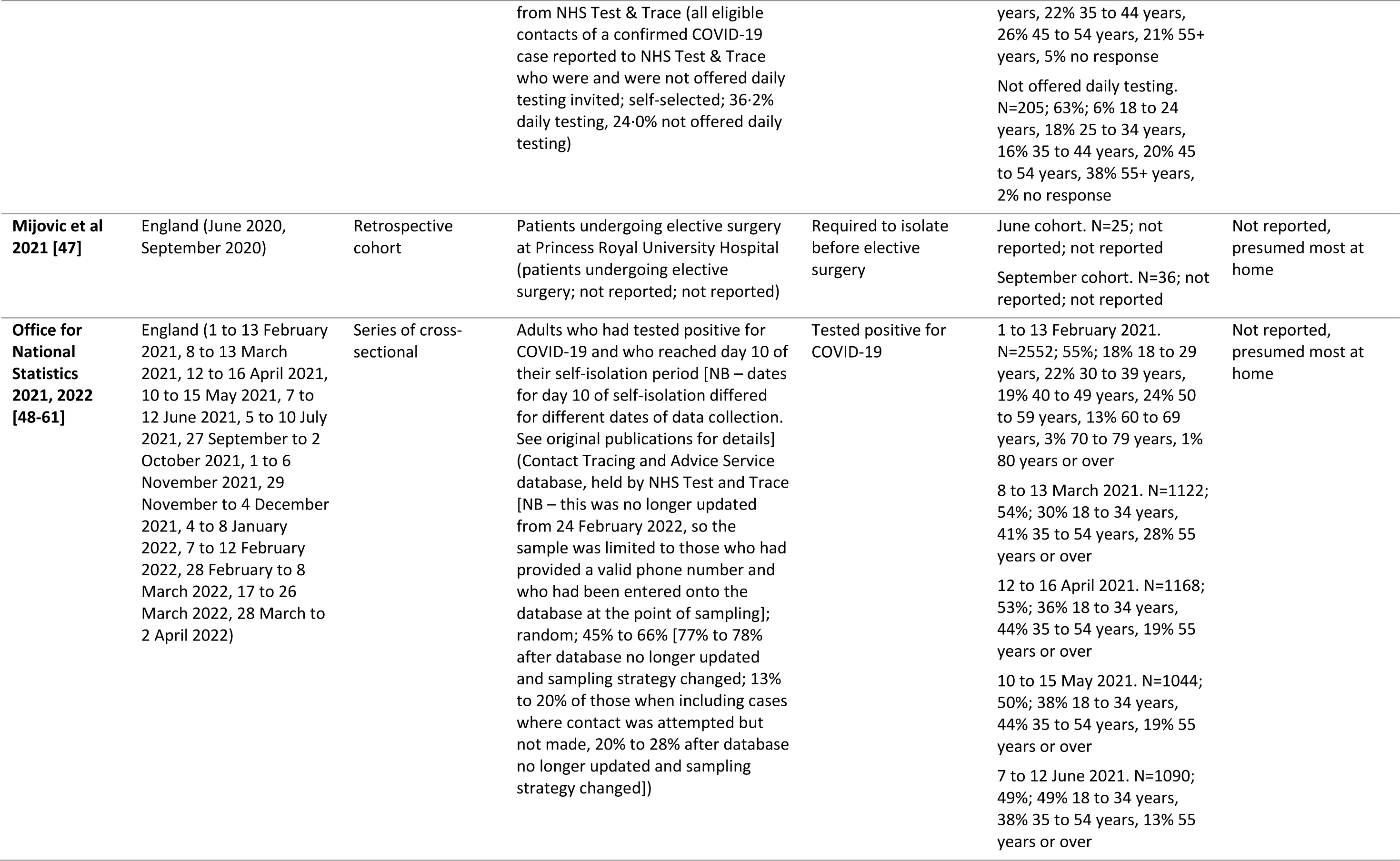
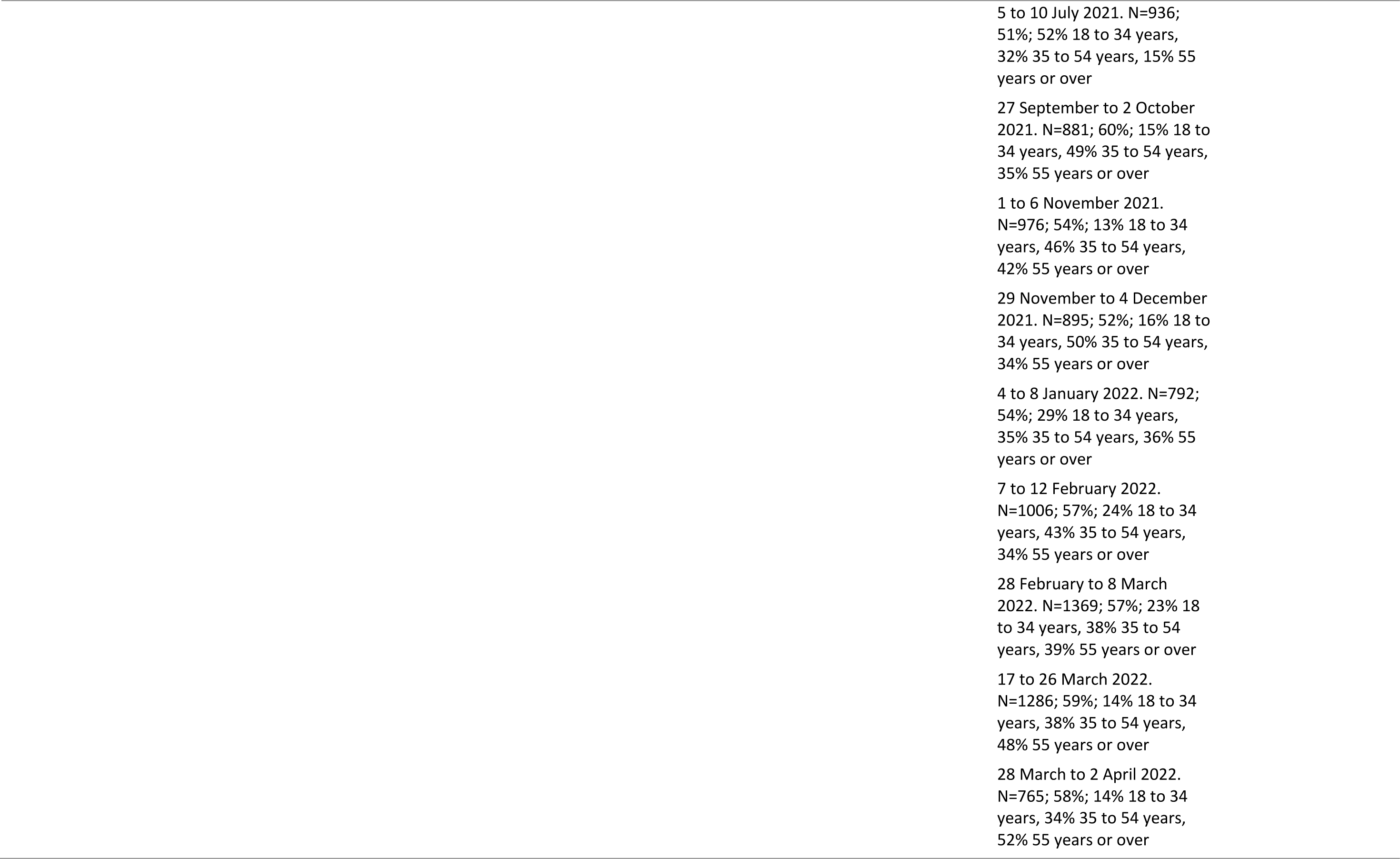
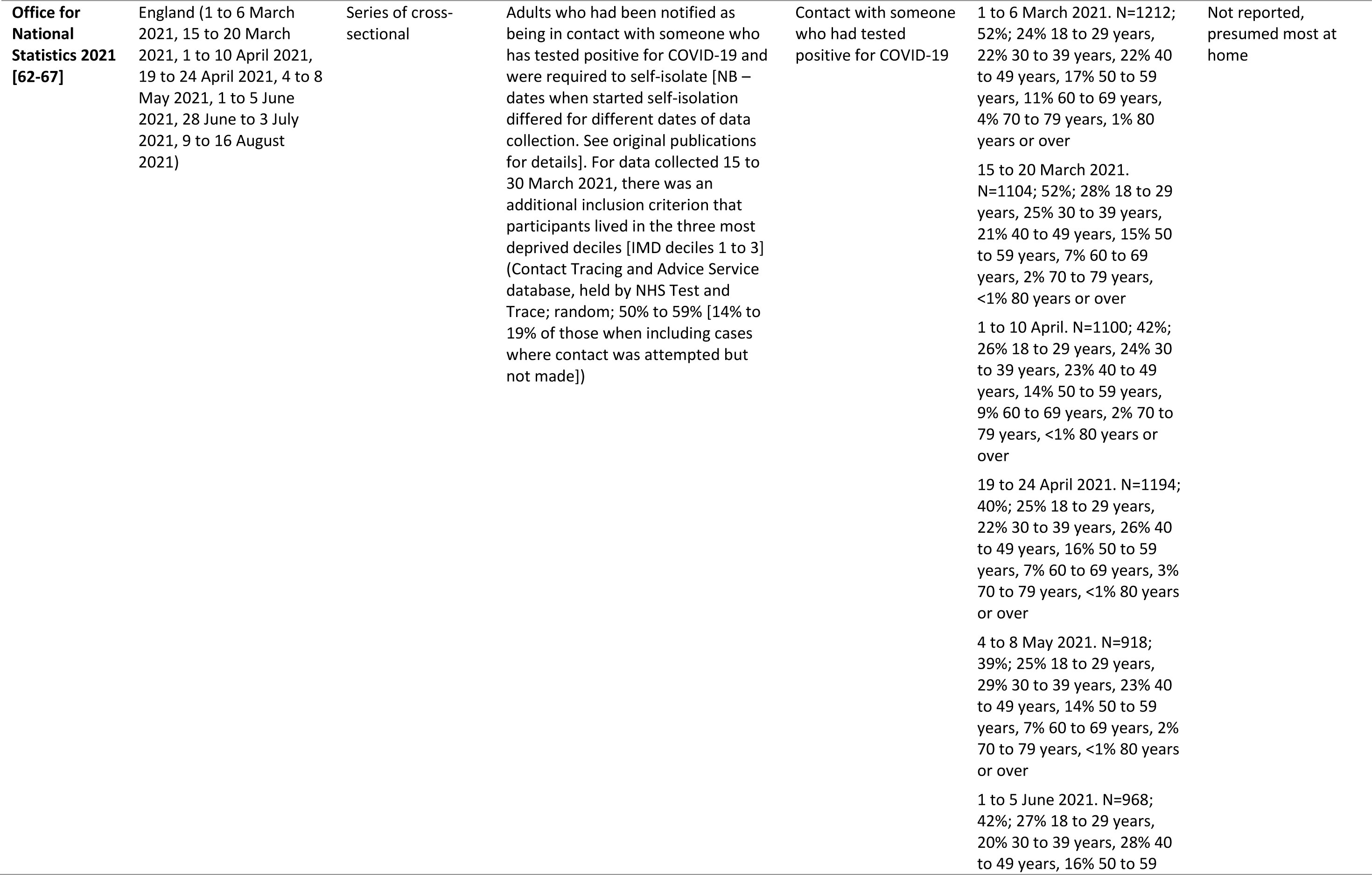
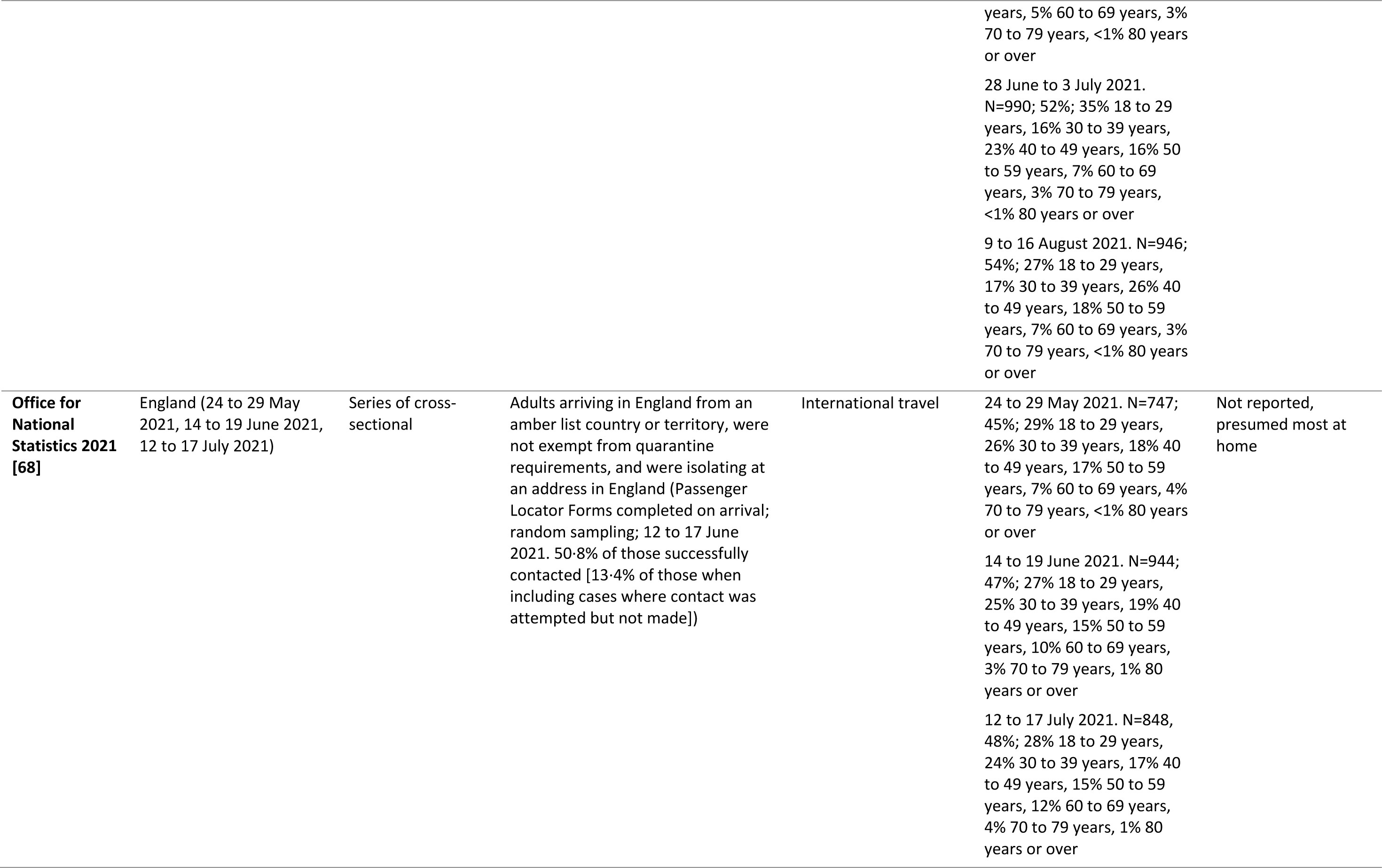
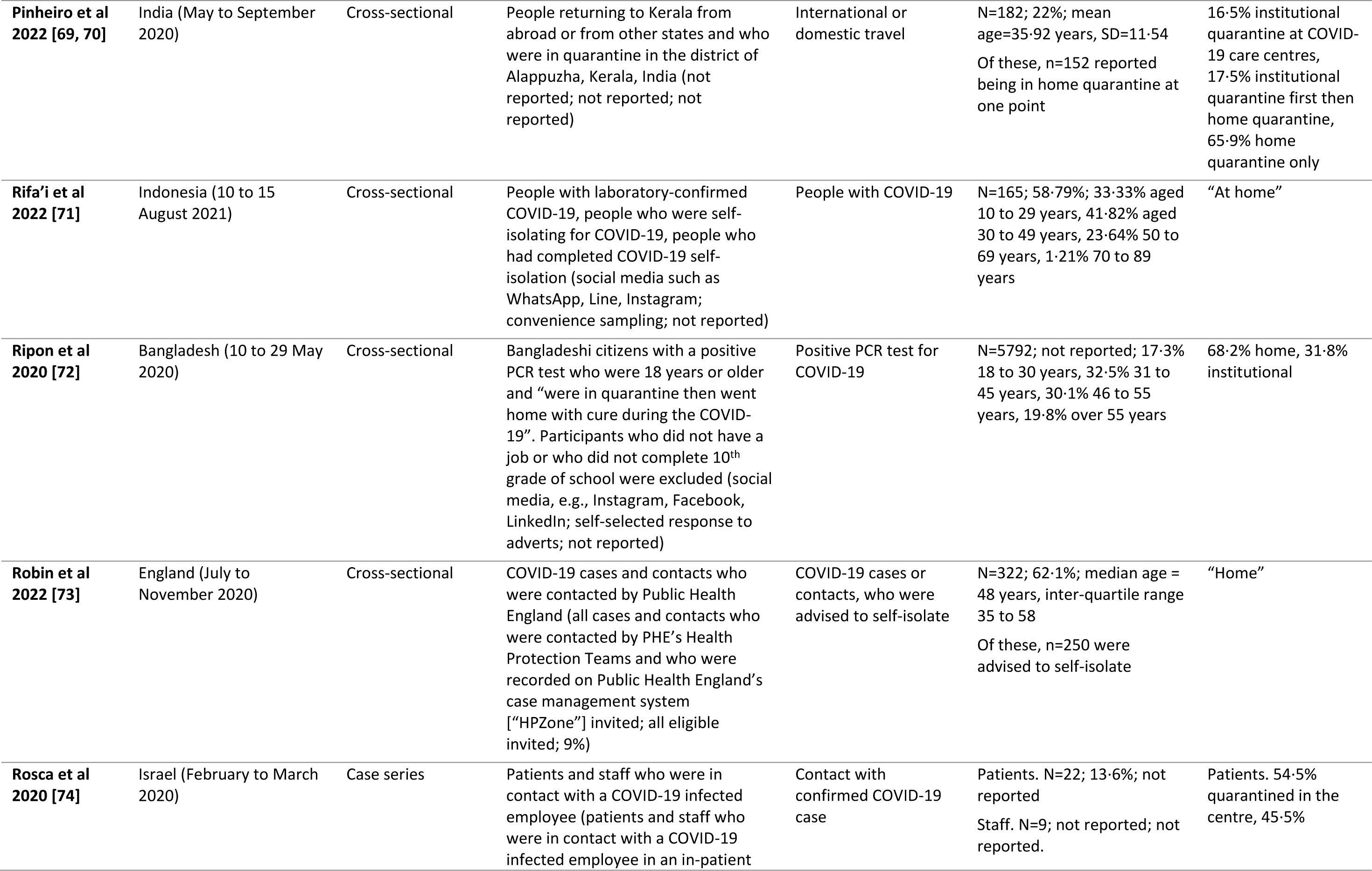
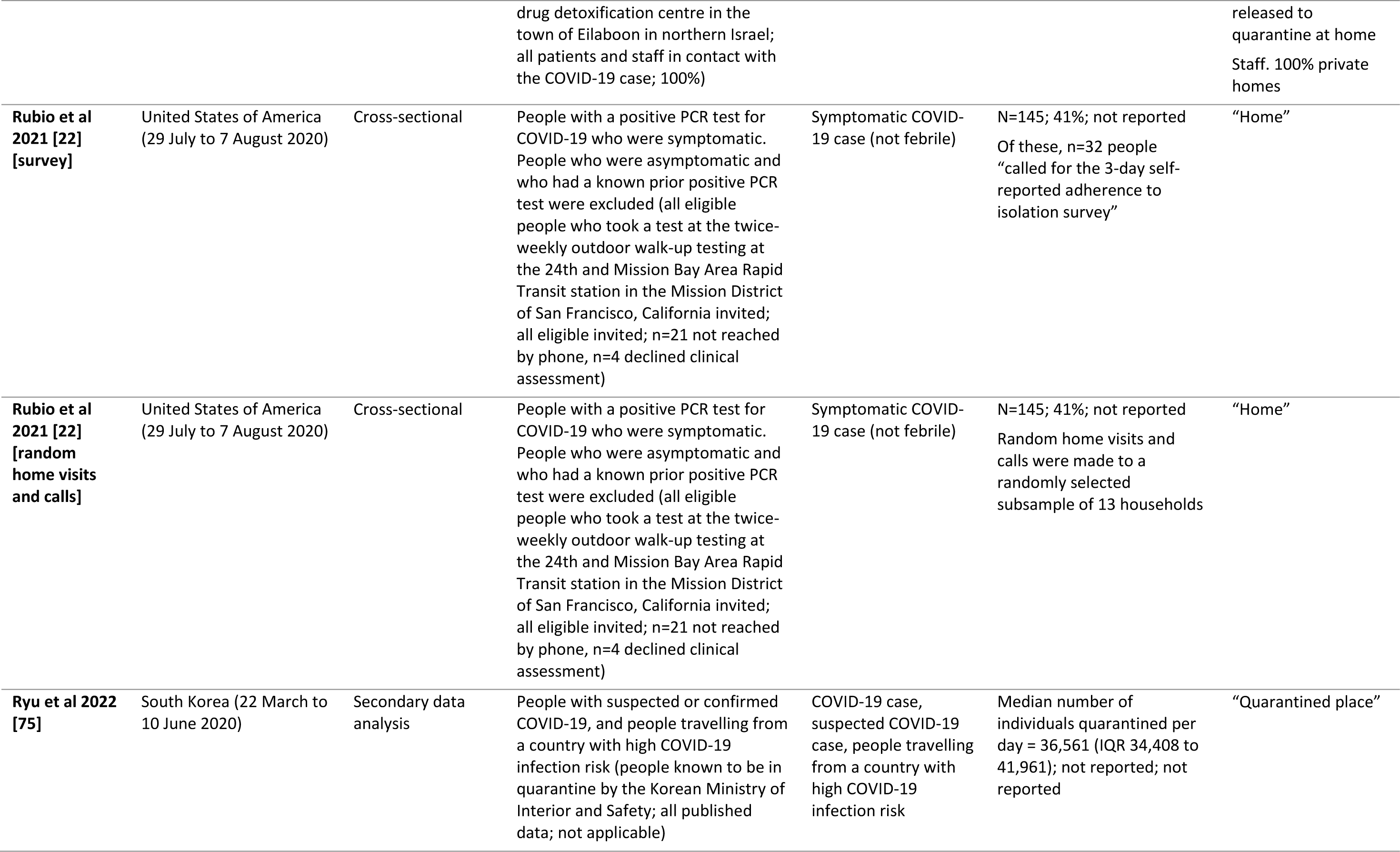
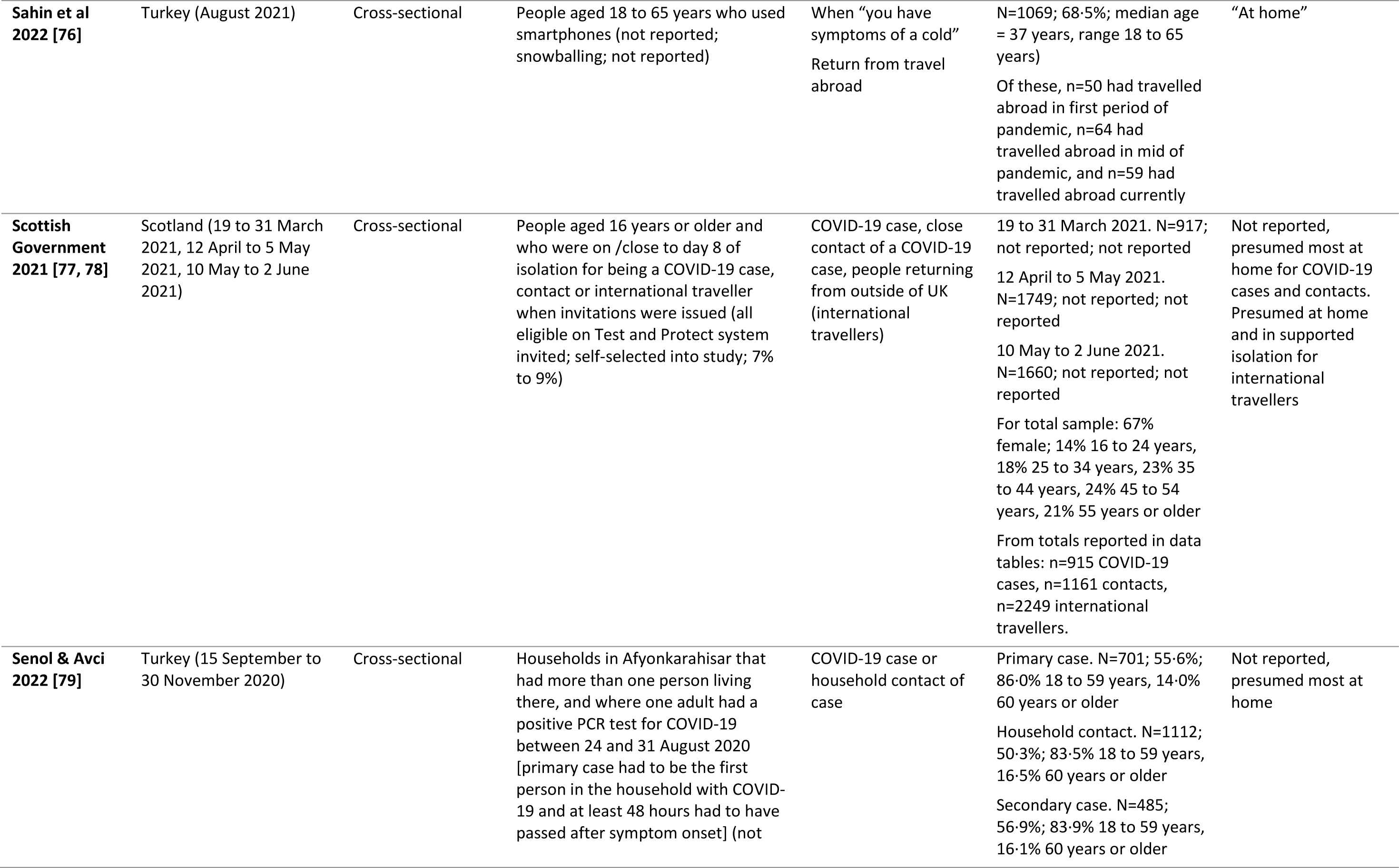
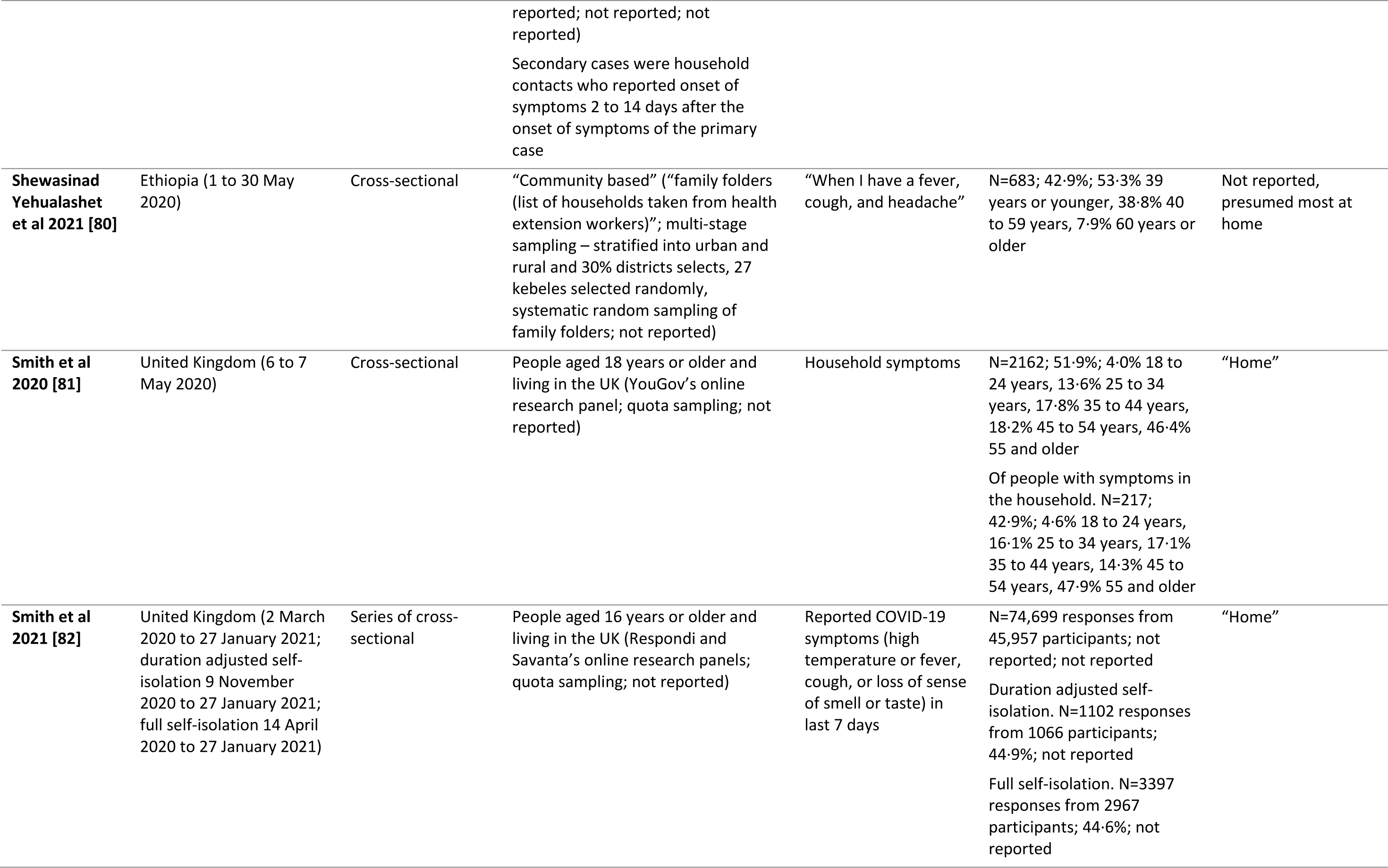
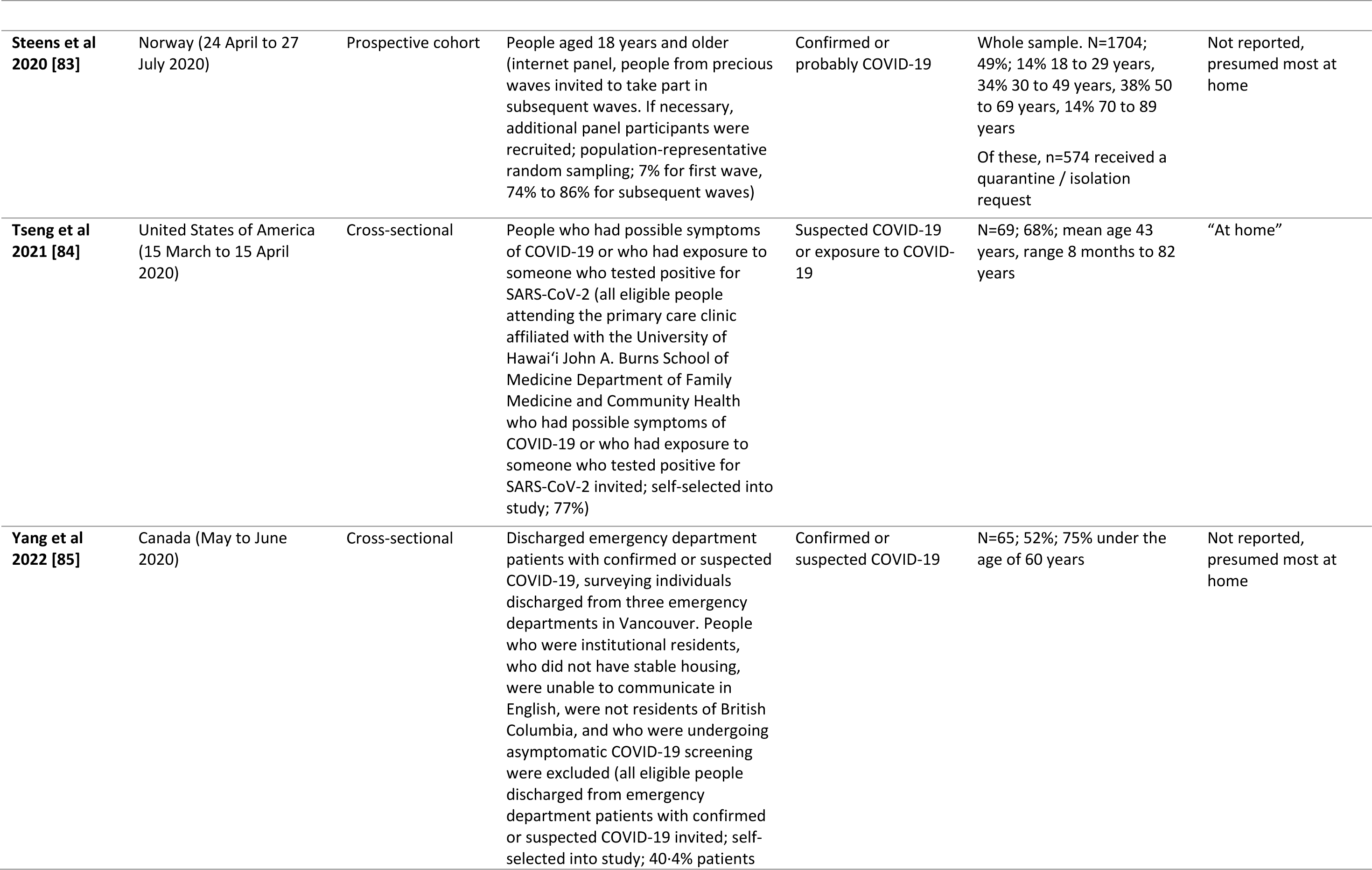
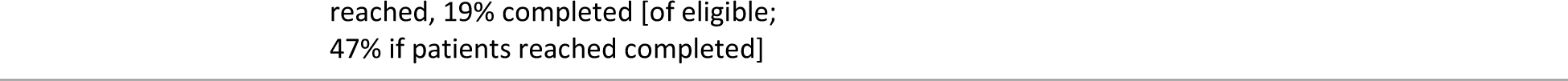
Study characteristics.

### Risk of bias

All but one study reported a prevalence estimate for rate of adherence to self-isolation. [41] One study reported two prevalence estimates. [76] Twenty prevalence estimates were at very high risk of bias, [21, 23, 24, 28, 29, 31–37, 40, 47, 69–72, 74, 76–78] 14 were at high risk, [21, 22, 27, 38, 44–46, 73, 79, 81–83, 85] ten had some concerns, [25, 26, 30, 39, 42, 43, 48–68, 80, 84] and one was at low risk. [75] Full details for each study are reported in Supplement S6.

### Definitions of self-isolation

Self-isolation was inconsistently defined (Supplementary Table S7). Seven studies defined self-isolation solely as staying at home, [22, 41, 68, 73, 81, 82] with another two studies specifying that self-isolation involved staying at home apart from a limited number of reasons. [46, 48–61] Where studies defined self-isolation using multiple behaviours, these are summarised in Table 2. In conjunction with other behaviours, the most mentioned behaviour was avoiding contact with others, followed by wearing a mask, and maintaining good hand hygiene. Five studies reported following official guidance in the country where the study was conducted, but did not specify what the guidance was. [21, 36, 37, 39] One study defined self-isolation as following “isolation measures (e.g., avoiding contact with others, hygiene measures, wearing a mask)”. [30] Twelve studies reported “self-isolate” or “quarantine” (or some such variant), but did not define required behaviours. [25, 33, 34, 42, 43, 47, 74, 75, 79, 80, 83–85] There was no mention of a definition of self-isolation in eight studies. [23, 24, 26–29, 31, 32]

**Table 2.** Summary table of definitions of self-isolation where studies specified multiple behaviours.

| Citation | Stay at home / isolation location | Not have visitors | Avoid contact with others | Wear a mask | Wash hands / hand hygiene | Stay in a separate room | Ventilate room | Disinfect room | Monitor condition / contact hospital if symptoms worsen |
| --- | --- | --- | --- | --- | --- | --- | --- | --- | --- |
| Eraso & Hills 2021 [35] | ✓ |  | ✓ |  |  |  |  |  |  |
| Foroozanfar et al 2020 [38] |  |  | ✓ |  |  | ✓ |  |  |  |
| Gasperini et al 2022 [40] |  |  | ✓ | ✓ | ✓ | ✓ |  |  |  |
| Li et al 2021 [44] |  |  | ✓ |  |  | ✓ | ✓ | ✓ | ✓ |
| Lou et al 2020 [45] |  |  | ✓ | ✓ | ✓ |  | ✓ | ✓ |  |
| Office for National Statistics 2021 [62-67] | ✓ | ✓ |  |  |  |  |  |  |  |
| Pinheiro et al 2022 [69, 70] |  |  | ✓ | ✓ | ✓ |  |  |  |  |
| Rifa'i et al 2022 [71] |  |  | ✓ |  |  |  | ✓ |  |  |
| Ripon et al 2020 [72] | ✓ |  |  | ✓ | ✓ |  |  |  | ✓ |
| Sahin et al 2022 [76] | ✓ |  | ✓ | ✓ |  |  |  |  |  |
| Scottish Government 2021 [77, 78] | ✓ | ✓ | ✓ |  |  |  |  |  |  |

### Measures of self-isolation

Only four studies did not use self-report to measure adherence to self-isolation. One study used a retrospective cohort chart review, with data about self-isolation adherence (reasons for leaving self-isolation early) extracted from clinical data. [39] One study used publicly available violation data (for self-isolation) from the Korean Ministry of Interior and Safety records. [75] Violation data were ascertained by actively monitoring mobile phone location data or phone calls twice a day, together with random visits to the self-isolating person. Another study used random home visits and calls to measure adherence to self-isolation. [22] People were categorised as non-adherent if they did not answer the door after three attempts or if external noise was heard on the telephone. Data were reported for seven participants, though the methods indicated that thirteen households were selected for this measure. One study did not report how they measured adherence to self-isolation. [74]

Thirty-nine studies used self-report measures of adherence to self-isolation (19 online surveys, 12 telephone surveys or interviews, 2 online and telephone, 6 did not report mode; Supplementary Table S7). One study used an in-person survey conducted by a nurse, the self-isolation adherence measure was “the general opinion of the home care nurses about the adherence to isolation/quarantine rules”. [40] Another study reported that data were collected by health care workers using an online survey platform. [80] Only one study reported psychometric characteristics of the adherence measure (Cronbach’s α; Table 3). [23] One other study commented that authors were “uncertain” whether their adherence measure had been correctly understood, indicating indirect evidence for a lack of validity of this measure. [83] Generally, studies reported little to no missing data (Table 3; Supplementary Table S7). Exceptions to this were online surveys which reported 13% to 67% missing data, [24, 30, 34] and a case interview that reported 23% missing data. [21] Only eleven studies reported verbatim the items and response options used to measure adherence to self-isolation, allowing reproduction in future studies. [29, 31, 32, 34, 35, 44, 68, 76, 79–81, 84] Measures often relied on presumed knowledge to be able to answer an item accurately (e.g., knowledge of self-isolation guidance) and used subjective scales (Table 3).

**Table 3.**
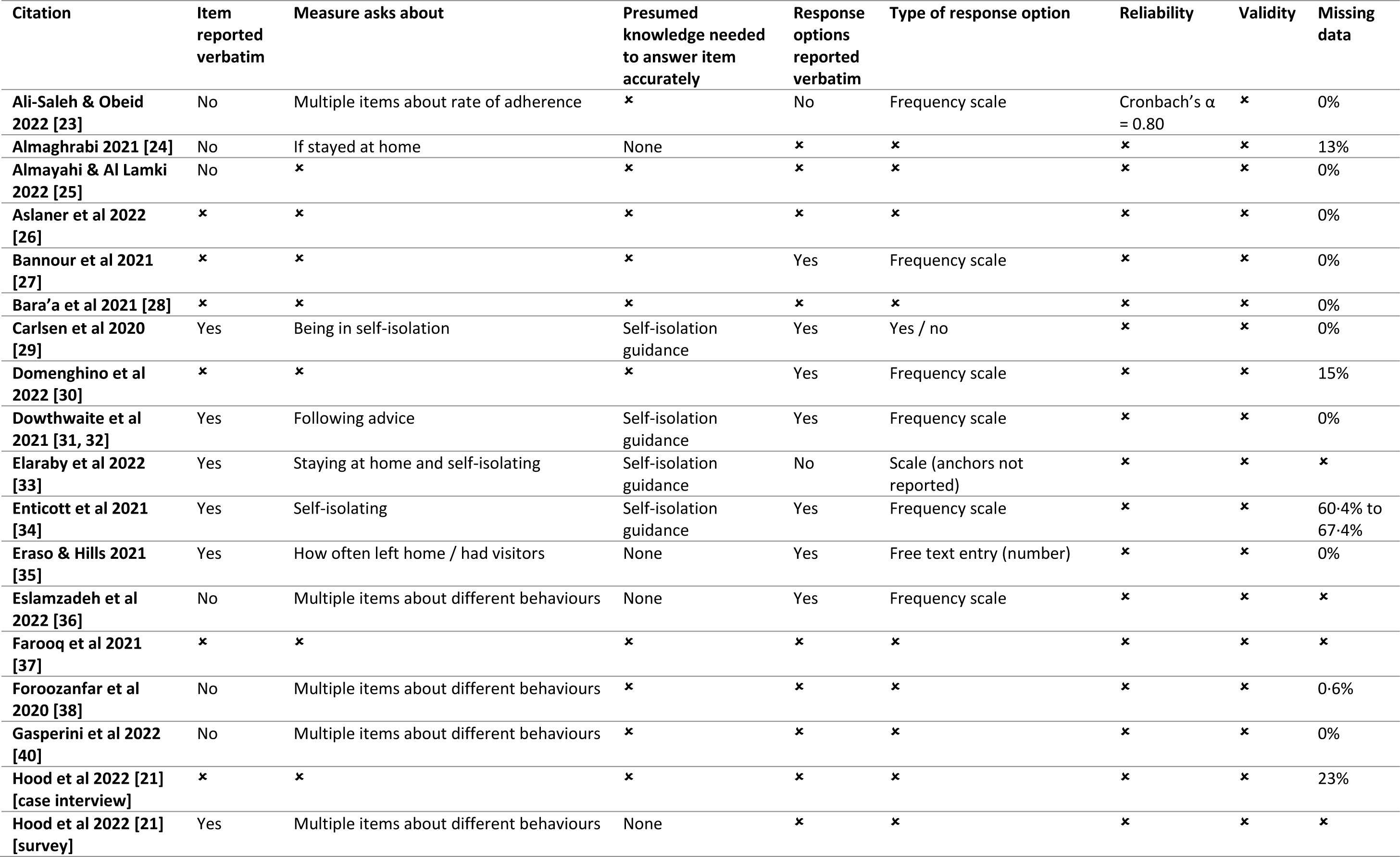
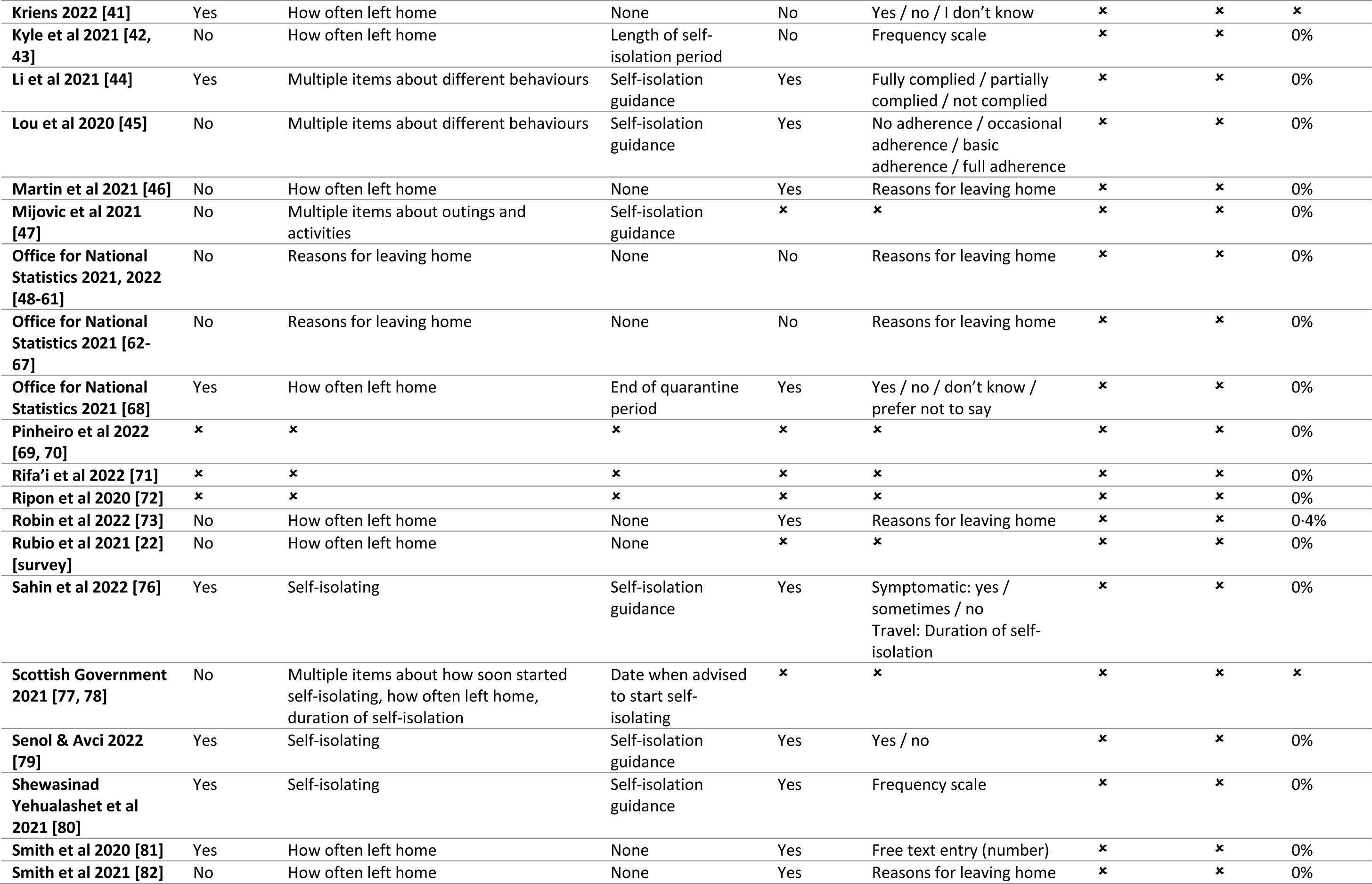
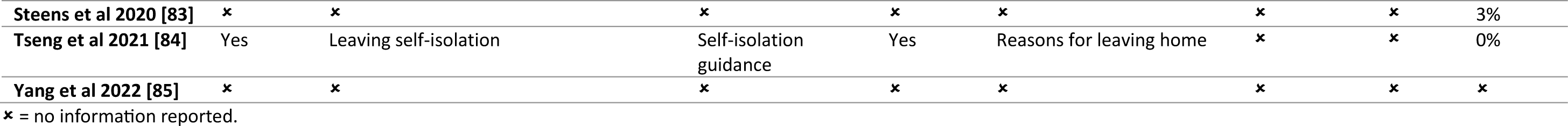
Properties of self-report measures for self-isolation adherence. Crosses indicate that no information was reported.

### Adherence to self-isolation

For COVID-19 cases, rates of adherence to self-isolation ranged between 0% and 100% (Figure 2, Supplementary Table S8). Studies with some concerns of bias reported rates between 51% and 86%; these studies generally had small confidence intervals (<5%). Studies with high or very high risk of bias had wider ranging estimates of adherence, and often had wider confidence intervals often due to smaller sample sizes. Self-isolation adherence for people with COVID-19-like symptoms ranged between 6·2% and 91·2% (Figure 2, Supplementary Table S8). The only study rated at some concerns of bias reported 16·4% adherence. [80]

**Figure 2.**
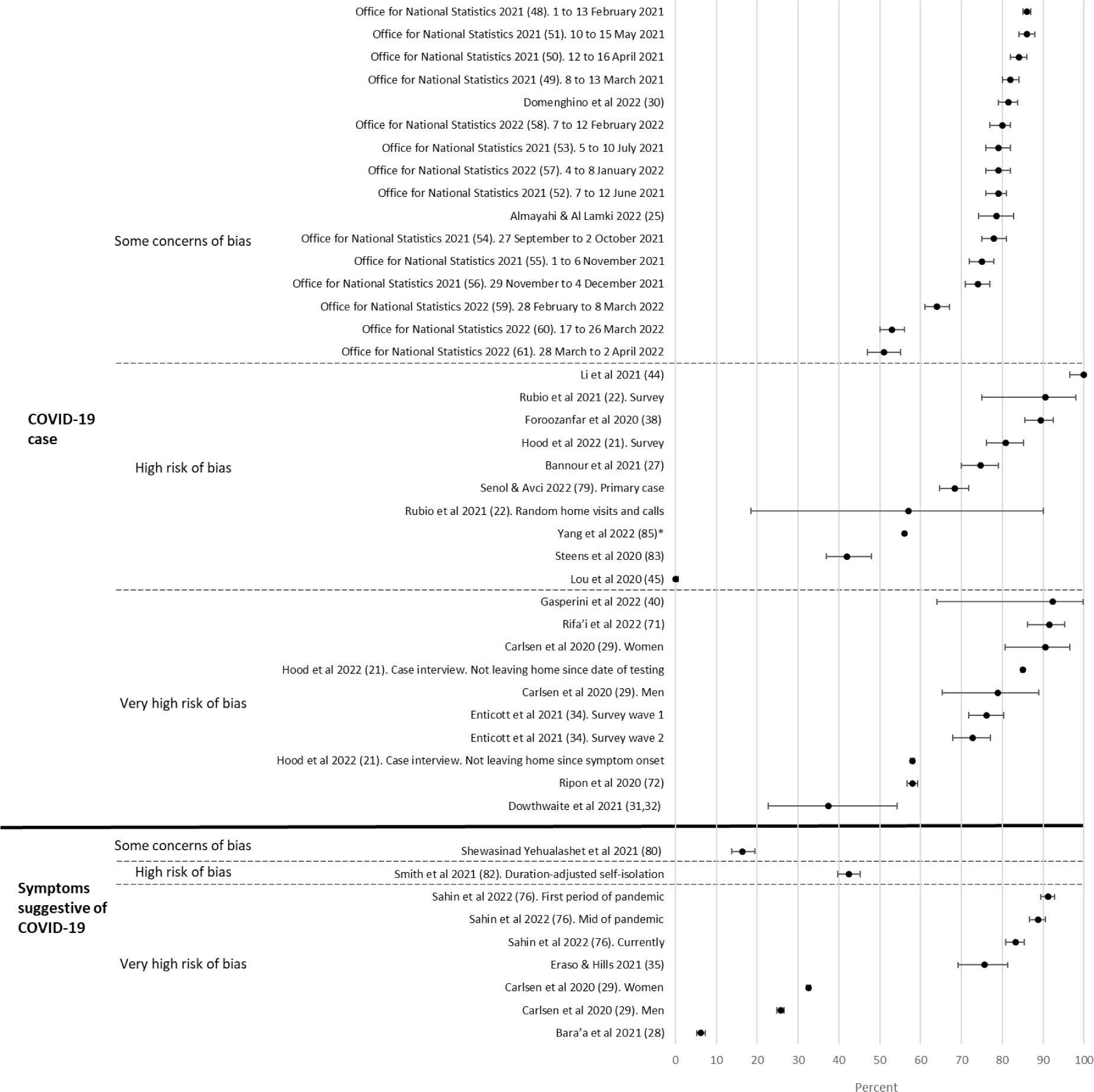
Forest plot showing rates of adherence to self-isolation in COVID-19 cases and people with symptoms suggestive of COVID-19 by risk of bias rating. Error bars are 95% confidence intervals. *95% confidence intervals could not be calculated accurately from the information given in the study

Self-isolation adherence in contacts of COVID-19 cases ranged from 26·6% to 94%. Studies at some concerns of bias reported adherence between 77·8% to 94% (Figure 3, Supplementary Table S8). Studies at high and very high risk of bias reported wider-ranging adherence rates and wider confidence intervals. Where cases and contacts were analysed together, adherence rates were between 24·9% and 97·4%; for studies rated as some concerns of bias, this was 53·6% to 97·4%. Self-isolation adherence for people returning from travel ranged between 15·3% to 86%, with rates in studies at some concerns of bias being between 78% and 86%. Other studies investigating this outcome were all at very high risk of bias and estimates had wide confidence intervals.

**Figure 3.**
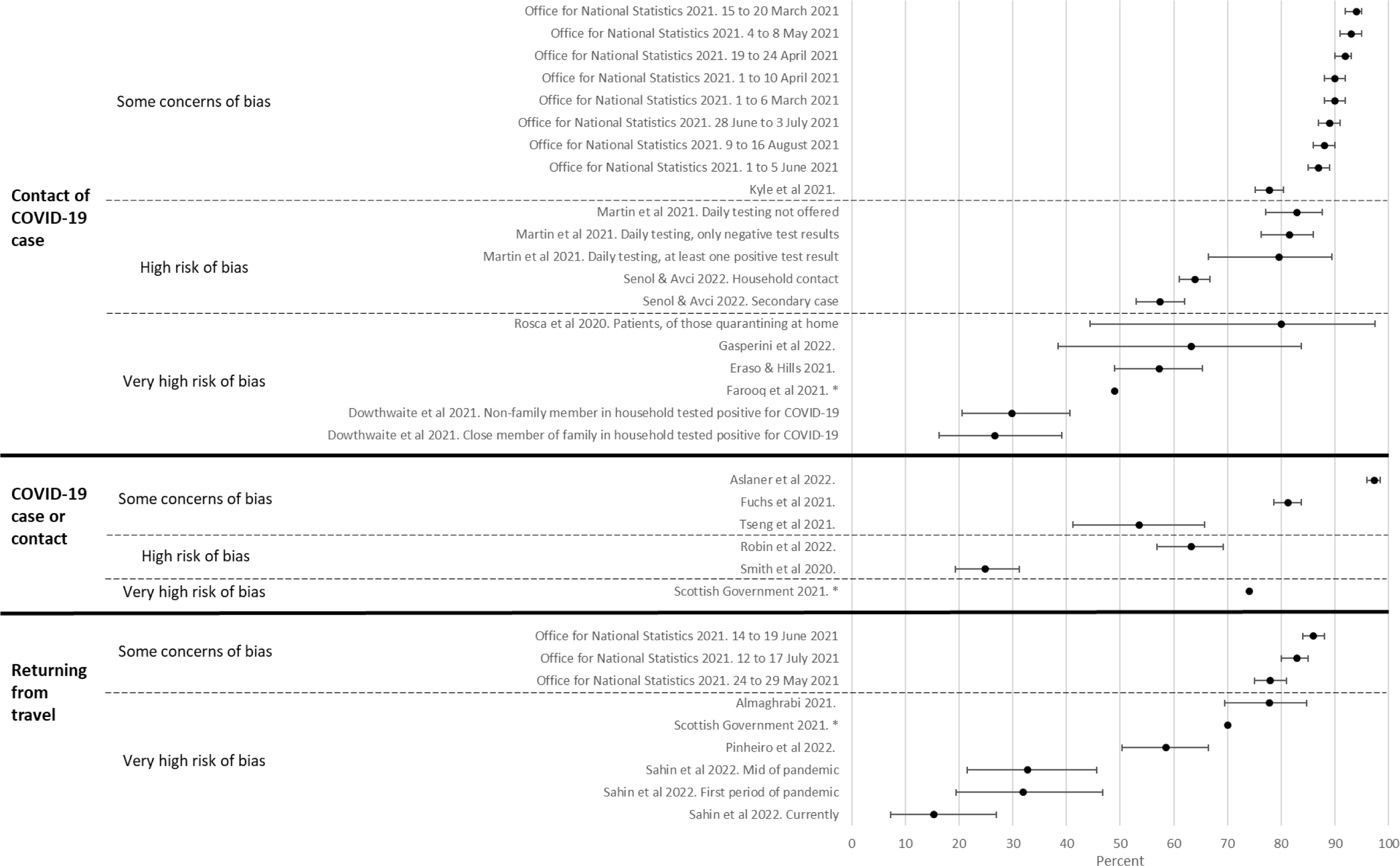
Forest plot showing rates of adherence to self-isolation in contacts of COVID-19 cases, COVID-19 cases and contacts together, and people returning from travel by risk of bias rating. Error bars are 95% confidence intervals. *95% confidence intervals could not be calculated accurately from the information given in the study

Studies investigating adherence to self-isolation for other reasons are reported in Supplementary Figure S9.

### Factors associated with self-isolation

Twenty-five studies investigated factors associated with adherence to self-isolation. [23, 29, 30, 34–43, 45–68, 73, 75–78, 81–83] Of these, 24 were assessed using the ROBINS-E. Analyses of associations from seven studies were at very high risk of bias, [30, 34, 37, 38, 73, 77, 78, 83], 15 were at high risk of bias, [23, 29, 35, 36, 40–43, 47–68, 75, 76, 81, 82] and two had some concerns (Supplement S6). [39, 45] One study was assessed using the ROBINS-I and was rated as very high risk of bias. [46] No analyses were at low risk of bias.

For brevity, few results are narratively summarised here; full details are in Supplement S10. Unless stated, all studies were high or very high risk of bias.

Overall, there was mixed evidence that socio-demographic factors were associated with adherence to self-isolation (Table 4, Supplementary Table S8). There was mixed evidence for an association between age and adherence. Five analyses found an association between adherence and older age (one at some concerns of bias), [34, 39, 41, 77, 78] three found an association with younger age, [29, 48–61, 83] and 12 found no evidence for an association. [23, 29, 30, 35, 36, 42, 43, 62–68, 73, 81, 82]

**Table 4.**
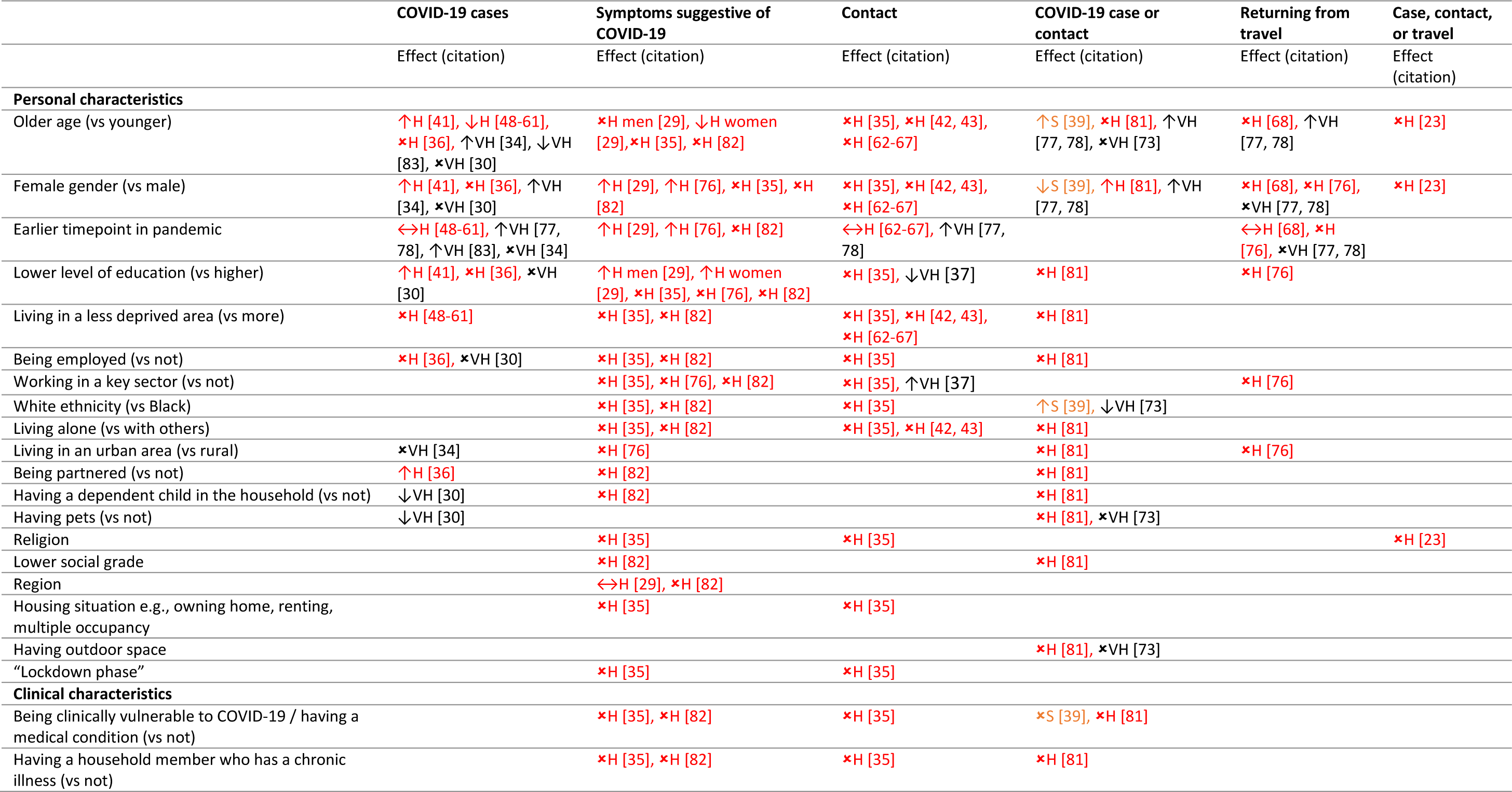
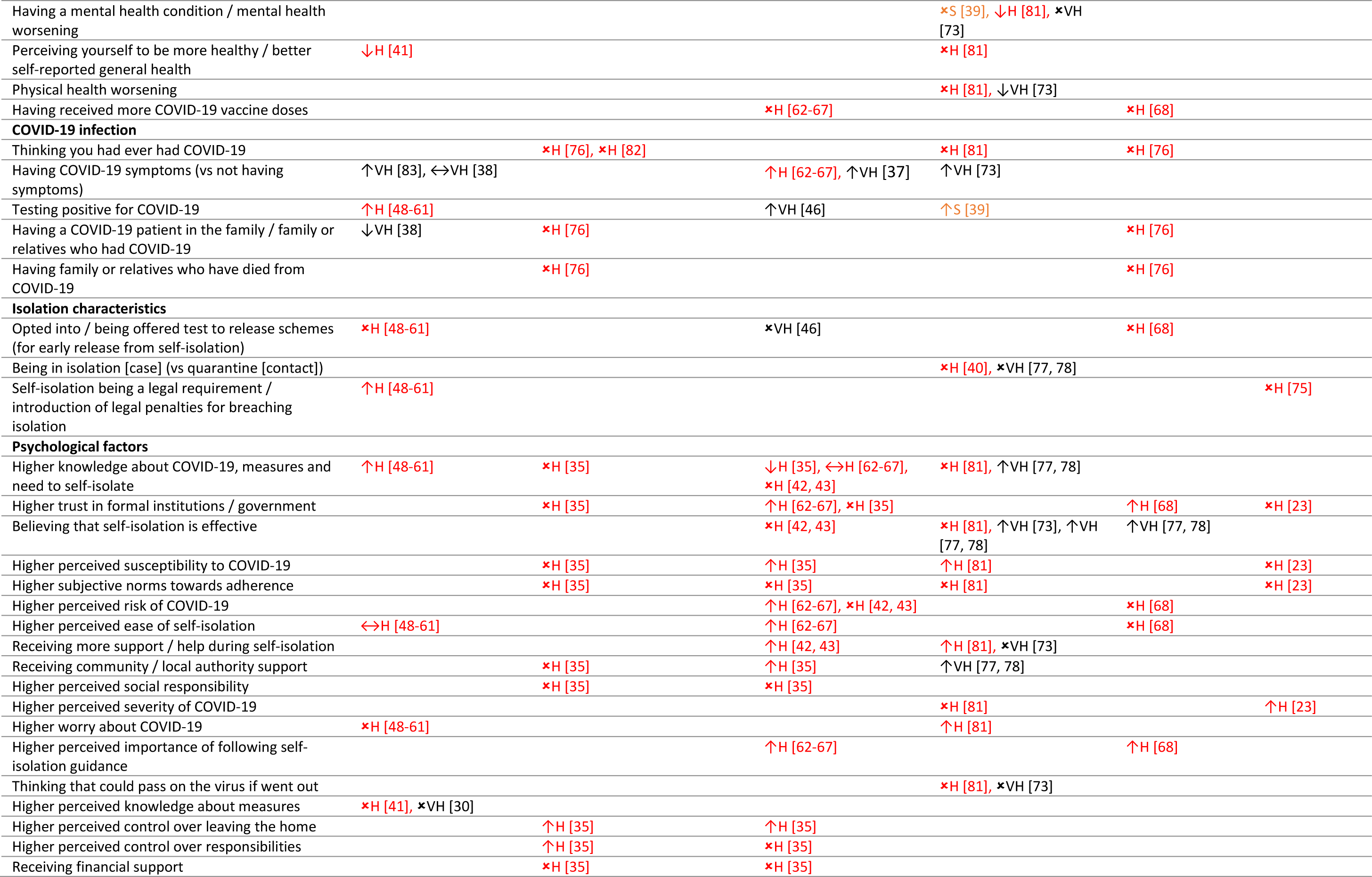
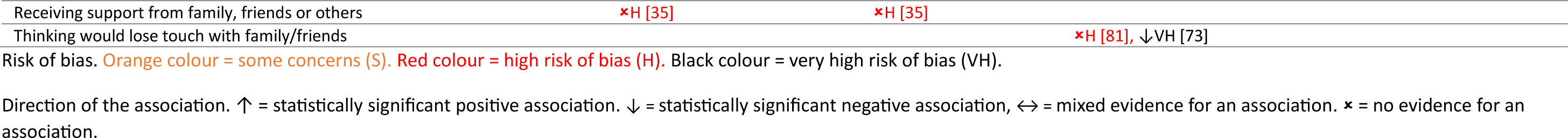
Summary table of personal and clinical characteristics associated with adherence to self-isolation by risk of bias. Colours relate to risk of bias. Studies in orange were rated at some concerns of bias (S), studies in red were rated as high risk of bias (H), and studies in black were rated as very high risk of bias (VH). Arrows indicate the direction of the association. Up arrows indicate a statistically significant positive association, down arrows indicate a statistically significant negative association, horizontal arrows indicate mixed evidence for an association, and crosses indicate that there is no evidence for an association.

There was weak evidence for an association with female gender and adherence, with six of 18 analyses finding an association. [29, 34, 41, 76–78, 81] Eleven analyses found no evidence for an association. [23, 30, 35, 36, 42, 43, 62–68, 76–78, 82] One analysis (some concerns of bias) found an association between being male and higher adherence. [39] This study investigated people experiencing homelessness or unstable housing and included a low percentage of women (24%).

Therefore, results are unlikely to be generalisable to the general population. There was weak evidence that lower education was associated with adherence with three analyses finding an association, [29, 41] and eight analyses finding no evidence for an association [30, 35, 36, 76, 81, 82]; one analysis found adherence to be associated with higher education. [37]

There was good evidence that adherence to self-isolation was associated with having COVID-19 symptoms, [37, 38, 62–67, 73, 83] and testing positive for COVID-19 (vs being a contact, testing negative; association found by one analysis at some concerns of bias; Table 4). [39, 46, 48–61] Taken together, there was some evidence that practical support may be associated with adherence to self-isolation (Table 4). The nature of an association between adherence to self-isolation and knowledge about COVID-19 and measures was unclear, with two analyses finding that higher knowledge was associated with adherence, [48–61, 77, 78] one analysis finding that lower knowledge was associated with adherence, [35] one analysis finding mixed evidence, [62–67] and three analyses finding no evidence for an association. [35, 42, 43, 81] There was weak evidence for an association between adherence to self-isolation and higher trust in the government and formal institutions, believing that self-isolation was effective, higher perceived importance of following self-isolation guidance, higher perceived susceptibility to COVID-19, higher perceived control over leaving home, and higher perceived ease of self-isolation (Table 4). There was little to no evidence for an association with adherence to self-isolation and clinical or isolation characteristics (Table 4).

## Discussion

This systematic review identified 45 studies investigating self-isolation during the COVID-19 pandemic. Most studies defined self-isolation as “staying at home”. Few studies included not receiving visitors, though this should have been an important consideration. The important parameters of self-isolation for a given illness depend on the clinical characteristics of infection. Future definitions of self-isolation, by public health agencies, policy makers and in academic research, should clearly specify behaviours that need to be enacted and for how long.

There was no standardized measure of self-isolation. Only four studies did not use self-report. [22, 39, 74, 75] While observing behaviour may result in more accurate rates of behaviour (higher external validity), observational studies may encounter other issues such as small sample sizes (e.g., n=7 in [74]), and time- and resource-consuming data collection (e.g., for door knocking). Of 41 studies using self-report data, only one reported on the reliability of the measure, [23] while another gave indirect evidence for the lack of validity of their measure, [83] highlighting the substantial risk of bias in self-isolation measurements. Studies often relied on participants’ understanding of self-isolation guidance – which was low [86] – to answer items accurately. While growing internet use means that online surveys are increasingly able to reach groups that previously did not use the internet (e.g., older adults), [87] more research is needed to quantify how self-report data from the quota samples often used in these studies match up to less biased methods. [8]

Rates of self-isolation adherence ranged between 0% and 100%. Studies with higher risk of bias often had more varied estimates and were more imprecise. Studies that used multiple rounds of data collection often reported similar prevalence estimates across rounds [48–68, 76] suggesting that study design, in particular sampling methods and phrasing of items, play a large role in adherence estimates. Using only studies with some concerns of bias, estimates of adherence were approximately 85% for COVID-19 cases (confirmed by a test), contacts, and people returning from travel, but only 16% for people with COVID-19-like symptoms who had not tested. While this may be due to individual study characteristics, [88] the next lowest risk of bias estimates for the COVID-19-like symptoms group reported 43% adherence. [82] This may be due to low knowledge of COVID-19 symptoms and the non-specific nature of cardinal COVID-19 symptoms (cough, high temperature), meaning that participants may not have attributed their symptoms to COVID-19. [89] Population-wide measures that rely on people self-isolating when symptomatic may be ineffective at controlling outbreaks, especially if symptoms are mild and non-specific, due to low adherence. Taken together, evidence suggests that public health messages should clearly specify symptoms and behaviours that should be enacted to increase adherence to self-isolation.

[3]There was good evidence that having symptoms (vs not) and having a positive COVID-19 test (vs negative or no test) were associated with adherence to self-isolation among identified cases and contacts. These associations are intuitive, with people with more signs of illness being more likely to adopt self-isolation. There was some evidence that receiving practical support is associated with adherence to self-isolation, in line with a previous systematic review. [3] Rigorously conducted randomised controlled trials of provision of support on adherence to self-isolation, investigating both practical (e.g., delivering groceries) and financial support, would strengthen the evidence base. There was little consistent evidence that socio-demographic factors were associated with adherence to self-isolation. A previous systematic review of factors associated with adherence to self-isolation also found little to no evidence for associations between adherence and socio-demographic factors. [3] Generally, evidence for associations between adherence to self-isolation and psychological factors was weak. Factors associated with adherence – higher trust in governments or formal institutions, believing self-isolation to be an effective way to prevent the spread of infection, and higher perceived susceptibility to COVID-19 – were similar to findings from a previous systematic review. [3] These factors are potentially modifiable, and could be targeted by public health campaigns in future outbreaks.

Most studies in the review were surveys using non-probabilistic sampling methods, therefore results may not be generalisable to the wider population. Most studies were cross-sectional, therefore causation is uncertain. The overall quality of included studies was low. Data were mainly self-report and may have been subject to social desirability and recall bias. Studies used a range of items to measure self-isolation and different cut-offs for what constituted “adherence”, with a significant minority of studies using items that relied on participants’ understanding of self-isolation. There is a good chance that participants misunderstood these items. There was a notable absence of randomised controlled trials investigating self-isolation adherence.

Human error means that we may have missed studies that should have been included. Studies in the review were heterogeneous in methods, materials, and definitions of self-isolation and adherence, meaning we were unable to conduct a meta-analysis. Through our grey literature search, we identified other studies that investigated adherence to self-isolation. However, these were not included as they did not investigate factors associated with adherence (an inclusion criterion for grey literature). [90–92] Other data reporting self-isolation adherence and associated factors exist but are not publicly available [93–95]; these were not included.

This systematic review describes and appraises definitions of self-isolation used by studies during the COVID-19 pandemic, measures of adherence, rates of adherence, and factors associated with adherence. Definitions and measures of self-isolation were wide-ranging, with no consensus on how self-isolation should be operationalized and no standardized measures. Only one study reported reliability data for their measure, highlighting the significant risk of bias in studies of self-isolation. More scientifically rigorous studies investigating adherence to self-isolation, especially randomized controlled trials of interventions to promote self-isolation, are needed.

## Contributors

LES, AFM, TMM and GJR conceptualised the study. AFM ran the systematic searches and curated the data. LES, AFM, SKB, RD, MVS and GJR screened citations. LES completed data extraction, risk of bias ratings and wrote the original draft. AFM, SKB, RD, MVS, RA, TMM and GJR reviewed and edited the manuscript. LES, AFM, SKB, RD, MVS, RA, TMM and GJR approved the final manuscript. LES and AFM are guarantors. The corresponding author attests that all listed authors meet authorship criteria and that no others meeting the criteria have been omitted.

## Declaration of interests

GJR advised the UK’s Office for National Statistics on its work relating to self-isolation; papers relating to this work were included as part of the review. LES, AFM, SKB, RD, MVS, RA, TMM, and GJR co-authored papers that were either included in this review or considered during the review process. RA is an employee of the UK Health Security Agency. LES, RA, TMM and GJR were participants of the UK’s Scientific Advisory Group for Emergencies or its subgroups. AFM, SKB, RD, and MVS report no competing interests.

## Data sharing

No novel data were collected as part of this study. All data are already publicly available.

## Funding statement

This study was funded by Research England Policy Support Fund 2022-23 (from the allocation to King’s College London). LES, AFB, SKB, RA and GJR are supported by the National Institute for Health and Care Research Health Protection Research Unit (NIHR HPRU) in Emergency Preparedness and Response, a partnership between the UK Health Security Agency, King’s College London and the University of East Anglia. The views expressed are those of the authors and not necessarily those of the NIHR, UKHSA, or the Department of Health and Social Care. For the purpose of open access, the author has applied a Creative Commons Attribution (CC BY) licence to any Author Accepted Manuscript version arising.

## Ethics

All data used were in the public domain, therefore ethical approval was not required.

## Data Availability

No novel data were collected as part of this study. All data are already publicly available.

## Notes

### Author Declarations

All data used were in the public domain, therefore ethical approval was not required.

