## Supplementary materials for "Measuring and increasing rates of self-isolation in the context of infectious diseases: A systematic review with narrative synthesis"

### Supplement S1. PRISMA checklist

| Section and Topic | Item # | Checklist item | Location where item is reported |
|---|---|---|---|
| **TITLE** | | | |
| Title | 1 | Identify the report as a systematic review. | P1 |
| **ABSTRACT** | | | |
| Abstract | 2 | See the PRISMA 2020 for Abstracts checklist. | P2 |
| **INTRODUCTION** | | | |
| Rationale | 3 | Describe the rationale for the review in the context of existing knowledge. | P3 |
| Objectives | 4 | Provide an explicit statement of the objective(s) or question(s) the review addresses. | P3 |
| **METHODS** | | | |
| Eligibility criteria | 5 | Specify the inclusion and exclusion criteria for the review and how studies were grouped for the syntheses. | P4, Supplement S5 |
| Information sources | 6 | Specify all databases, registers, websites, organisations, reference lists and other sources searched or consulted to identify studies. Specify the date when each source was last searched or consulted. | P4 |
| Search strategy | 7 | Present the full search strategies for all databases, registers and websites, including any filters and limits used. | Supplement S3 |
| Selection process | 8 | Specify the methods used to decide whether a study met the inclusion criteria of the review, including how many reviewers screened each record and each report retrieved, whether they worked independently, and if applicable, details of automation tools used in the process. | P4 |
| Data collection process | 9 | Specify the methods used to collect data from reports, including how many reviewers collected data from each report, whether they worked independently, any processes for obtaining or confirming data from study investigators, and if applicable, details of automation tools used in the process. | P4 |
| Data items | 10a | List and define all outcomes for which data were sought. Specify whether all results that were compatible with each outcome domain in each study were sought (e.g. for all measures, time points, analyses), and if not, the methods used to decide which results to collect. | P4, Supplement S5 |
| | 10b | List and define all other variables for which data were sought (e.g. participant and intervention characteristics, funding sources). Describe any assumptions made about any missing or unclear information. | P4, Supplement S5 |
| Study risk of bias assessment | 11 | Specify the methods used to assess risk of bias in the included studies, including details of the tool(s) used, how many reviewers assessed each study and whether they worked independently, and if applicable, details of automation tools used in the process. | P4, Supplement S4 |
| Effect measures | 12 | Specify for each outcome the effect measure(s) (e.g. risk ratio, mean difference) used in the synthesis or presentation of results. | Supplement S5 |
| Synthesis methods | 13a | Describe the processes used to decide which studies were eligible for each synthesis (e.g. tabulating the study intervention characteristics and comparing against the planned groups for each synthesis (item #5)). | P5, Supplement S5 |

| Section and Topic | Item # | Checklist item | Location where item is reported |
|---|---|---|---|
| | 13b | Describe any methods required to prepare the data for presentation or synthesis, such as handling of missing summary statistics, or data conversions. | P5, Supplement S5 |
| | 13c | Describe any methods used to tabulate or visually display results of individual studies and syntheses. | Supplement S5 |
| | 13d | Describe any methods used to synthesize results and provide a rationale for the choice(s). If meta-analysis was performed, describe the model(s), method(s) to identify the presence and extent of statistical heterogeneity, and software package(s) used. | P5, Supplement S5 |
| | 13e | Describe any methods used to explore possible causes of heterogeneity among study results (e.g. subgroup analysis, meta-regression). | P5, Supplement S5 |
| | 13f | Describe any sensitivity analyses conducted to assess robustness of the synthesized results. | N/A |
| Reporting bias assessment | 14 | Describe any methods used to assess risk of bias due to missing results in a synthesis (arising from reporting biases). | P4, Supplement S4 |
| Certainty assessment | 15 | Describe any methods used to assess certainty (or confidence) in the body of evidence for an outcome. | Supplement S5 |
| **RESULTS** | | | |
| Study selection | 16a | Describe the results of the search and selection process, from the number of records identified in the search to the number of studies included in the review, ideally using a flow diagram. | P5, Figure 1 |
| | 16b | Cite studies that might appear to meet the inclusion criteria, but which were excluded, and explain why they were excluded. | P5, Figure 1 |
| Study characteristics | 17 | Cite each included study and present its characteristics. | P6-26 |
| Risk of bias in studies | 18 | Present assessments of risk of bias for each included study. | P27, p36, Supplement S6, Supplementary Table S8 |
| Results of individual studies | 19 | For all outcomes, present, for each study: (a) summary statistics for each group (where appropriate) and (b) an effect estimate and its precision (e.g. confidence/credible interval), ideally using structured tables or plots. | P27-40 |
| Results of syntheses | 20a | For each synthesis, briefly summarise the characteristics and risk of bias among contributing studies. | P27-40 |
| | 20b | Present results of all statistical syntheses conducted. If meta-analysis was done, present for each the summary estimate and its precision (e.g. confidence/credible interval) and measures of statistical heterogeneity. If comparing groups, describe the direction of the effect. | P27-40 |
| | 20c | Present results of all investigations of possible causes of heterogeneity among study results. | N/A |
| | 20d | Present results of all sensitivity analyses conducted to assess the robustness of the synthesized results. | N/A |
| Reporting biases | 21 | Present assessments of risk of bias due to missing results (arising from reporting biases) for each synthesis assessed. | P27-40, Supplement S6 |
| Certainty of evidence | 22 | Present assessments of certainty (or confidence) in the body of evidence for each outcome assessed. | P27-40 |
| **DISCUSSION** | | | |

| Section and Topic | Item # | Checklist item | Location where item is reported |
|---|---|---|---|
| Discussion | 23a | Provide a general interpretation of the results in the context of other evidence. | P40-41 |
| | 23b | Discuss any limitations of the evidence included in the review. | P41 |
| | 23c | Discuss any limitations of the review processes used. | P31 |
| | 23d | Discuss implications of the results for practice, policy, and future research. | P40-41 |
| **OTHER INFORMATION** | | | |
| Registration and protocol | 24a | Provide registration information for the review, including register name and registration number, or state that the review was not registered. | P2, p4 |
| | 24b | Indicate where the review protocol can be accessed, or state that a protocol was not prepared. | P2, p4 |
| | 24c | Describe and explain any amendments to information provided at registration or in the protocol. | Supplement S2 |
| Support | 25 | Describe sources of financial or non-financial support for the review, and the role of the funders or sponsors in the review. | P2, p5 |
| Competing interests | 26 | Declare any competing interests of review authors. | P42 |
| Availability of data, code and other materials | 27 | Report which of the following are publicly available and where they can be found: template data collection forms; data extracted from included studies; data used for all analyses; analytic code; any other materials used in the review. | P42 |

*From:* Page MJ, McKenzie JE, Bossuyt PM, Boutron I, Hoffmann TC, Mulrow CD, et al. The PRISMA 2020 statement: an updated guideline for reporting systematic reviews. BMJ 2021;372:n71. doi: 10.1136/bmj.n71

For more information, visit: http://www.prisma-statement.org/

#### Supplement S2. Deviations from the protocol

In addition to the databases mentioned in the protocol, we also searched Embase.

In the protocol we stated that risk of bias would be assessed using the relevant NIH tool for the study design (https://www.nhlbi.nih.gov/health-topics/study-quality-assessment-tools). However, upon reviewing the tool prior to data extraction, we felt that the NIH tools did not allow for sensitivity to differentiate between studies. Furthermore, the final rating ("good", "fair" or "poor") was left up to the reviewer's discretion. We therefore consulted the Cochrane handbook for their recommended tool and came across the Risk Of Bias In Non-randomized Studies (Exposure / Intervention; ROBINS-E/I). We decided to use these tools to assess analyses of associations for factors associated with adherence.

The ROBINS tools are not appropriate to assess prevalence estimates. We performed a quick search of the literature for recommended risk of bias tools to assess prevalence estimates. Two systematic reviews recommended the Joanna Briggs Institute Prevalence Critical Appraisal Tool. [1, 2] Therefore, we decided to use an adapted version of the Joanna Briggs Institute Prevalence Critical Appraisal Tool to appraise prevalence estimates.

### Supplement S3. Full details of searches used

#### Search terms used for database searches

| # | Query |
|---|---|
| **1** | (coronavirus or covid* or sars-cov-2 or ncov2019).mp. or exp Coronavirus/ or exp COVID-19/ or exp SARS-CoV-2/ |
| **2** | (isolat* or quarantin* or confinement).mp. or exp Patient Isolation/ or exp Quarantine/ |
| **3** | 2 not "social isolation".mp. [mp=title, book title, abstract, original title, name of substance word, subject heading word, floating sub-heading word, keyword heading word, organism supplementary concept word, protocol supplementary concept word, rare disease supplementary concept word, unique identifier, synonyms] |
| **4** | (adheren* or compliance or wellbeing or well-being or "quality of life" or resilien* or coping or flourish* or "positive psychology" or "posttraumatic growth" or "post-traumatic growth" or "life satisfaction" or "personal satisfaction" or "psychosocial functioning" or "mental health" or anxiety or depress* or ptsd or trauma* or psychiatric or "psychological stress" or "social stigma" or distress* or mood* or emotion* or "substance abuse" or "substance misuse" or "substance use" or "hazardous drinking" or "alcohol use" or "alcohol abuse" or "alcohol misuse" or alcoholi* or sleep or insomnia or loneliness).mp. or exp Guideline Adherence/ or exp "Treatment Adherence and Compliance"/ or exp Compliance/ or exp Patient Compliance/ or exp "Quality of Life"/ or exp Resilience, Psychological/ or exp Psychology, Positive/ or exp Posttraumatic Growth, Psychological/ or exp Personal Satisfaction/ or exp Psychosocial Functioning/ or exp Mental Health/ or exp Anxiety Disorders/ or exp Anxiety/ or exp Panic/ or exp Panic Disorder/ or exp Depression/ or exp Stress Disorders, Post-Traumatic/ or exp Psychological Trauma/ or exp Stress, Psychological/ or exp Social Stigma/ or Psychological Distress/ or exp Emotions/ or exp Sleep/ or exp "Sleep Initiation and Maintenance Disorders"/ or exp Substance Abuse, Intravenous/ or exp Substance-Related Disorders/ or exp Alcoholism/ or exp Alcohol Drinking/ or exp Loneliness/ |
| **5** | 1 and 3 and 4 |
| **6** | limit 5 to (humans and yr="2020 -Current") |

The search functions for registers (medRxiv, PsyArXiv) are not suitable for use in systematic reviews due to multiple limitations. Specifically, confusing Boolean operators, a lack of reproducibility, and no batch export. To overcome this, preprint searches were extracted using the R package medrxivr. [3, 4] Code is available on request.

#### Grey literature searches

Websites

| N of websites | Website name | Website link(s) | Reports sought for retrieval | Reports not retrieved | Reports assessed for eligibility | Reports included | Reports excluded |
|---|---|---|---|---|---|---|---|
| **1** | Rijksinstituut voor Volksgezondheid en Milieu (Dutch National Institute for Public Health and the Environment) | https://www.rivm.nl/en/coronavirus-covid-19/research/behaviour | 21 | 0 | 21 | 0 | 21 |
| **2** | iCARE (international COVID-19 Awareness and Responses Evaluation) Study | https://icare.mbmc-cmcm.ca/results-findings/results-1/ | 1 | 0 | 1 | 0 | 1 |
| **3** | | https://www.mbmc-cmcm.ca/2021/covid19/results-findings/infographics/ | 1 | 0 | 1 | 0 | 1 |
| **4** | | https://www.mbmc-cmcm.ca/2021/covid19/results-findings/publications/ | 0 | 0 | 0 | 0 | 0 |
| **5** | CHARIS (Covid Health and Adherence Research In Scotland) Study | https://www.abdn.ac.uk/iahs/research/health-psychology/publications-documents-2174.php | 26 | 0 | 26 | 0 | 26 |
| | | TOTAL | 49 | 0 | 49 | 0 | 49 |

Organisations

| N of organisations | Organisation | Website link(s) | Reports sought for retrieval | Reports not retrieved | Reports assessed for eligibility | Reports included | Reports excluded |
|---|---|---|---|---|---|---|---|
| **1** | UK COVID-19: testing initiative evaluation programme | https://www.gov.uk/government/collections/covid-19-testing-initiative-evaluation-programme | 7 | 0 | 7 | 0 | 7 |
| **2** | Office for National Statistics | https://www.ons.gov.uk/ | 42 | 0 | 42 | 21 | 21 |
| **3** | Welsh Government | https://www.gov.wales/ | 4 | 0 | 4 | 0 | 4 |
| **4** | Public Health Wales | https://phw.nhs.wales/ | 4 | 0 | 4 | 2 | 2 |
| **5** | StatsWales | https://statswales.gov.wales/Catalogue | 0 | 0 | 0 | 0 | 0 |
| **6** | Scottish Government | https://www.gov.scot/ | 26 | 0 | 26 | 2 | 24 |
| **7** | Public Health | https://publichealthscotland.scot/ | 13 | 0 | 13 | 0 | 13 |

| | | | | | | | |
|---|---|---|---|---|---|---|---|
| | Scotland | | | | | | |
| **8** | NI Direct | https://www.nidirect.gov.uk/ | 0 | 0 | 0 | 0 | 0 |
| **9** | Department of Health, Northern Ireland | https://www.health-ni.gov.uk/covid-19-statistics | 2 | 0 | 2 | 0 | 2 |
| **10** | Northern Ireland Executive | https://www.northernireland.gov.uk/publications | 0 | 0 | 0 | 0 | 0 |
| **11** | HSC Public Health Agency (Northern Ireland) | https://www.publichealth.hscni.net/ | 1 | 0 | 1 | 0 | 1 |
| **12** | Northern Ireland Statistics and Research Agency | https://www.nisra.gov.uk/statistics/ni-summary-statistics/coronavirus-covid-19-statistics | 1 | 0 | 1 | 0 | 1 |
| **13** | Government of Ireland | https://www.gov.ie/en/ | 0 | 0 | 0 | 0 | 0 |
| **14** | Central Statistics Office Ireland | https://www.cso.ie/en/index.html | 1 | 0 | 1 | 0 | 1 |
| | | TOTAL: | 101 | 0 | 101 | 25 | 76 |

Grey literature databases

| **N of databases** | **Website name** | **Website link(s)** | **Reports sought for retrieval** | **Reports not retrieved** | **Reports assessed for eligibility** | **Reports included** | **Reports excluded** |
|---|---|---|---|---|---|---|---|
| **1** | Opengrey.eu | https://opengrey.eu | 0 | 0 | 0 | 0 | 0 |
| **2** | WHO | https://www.who.int/en/ | 0 | 0 | 0 | 0 | 0 |
| **3** | NTIS (US Department of Commerce) | https://www.ntis.gov | 0 | 0 | 0 | 0 | 0 |
| **4** | WorldCat | https://www.worldcat.org | 12 | 0 | 12 | 1 | 11 |
| **5** | Agency for Healthcare Research and Quality | https://www.ahrq.gov | 0 | 0 | 0 | 0 | 0 |
| | | TOTAL | 12 | 0 | 12 | 1 | 11 |

Studies already identified in other searches are not included here, so as not to double count them.

Google search

| N of websites | Citation | Website link | Reports sought for retrieval | Reports not retrieved | Reports assessed for eligibility | Reports included | Reports excluded |
|---|---|---|---|---|---|---|---|
| **1** | Scottish Government 2021 | https://www.gov.scot/publications/compliance-self-isolation-quarantine-measures-literature-review/pages/4/ | * | * | * | * | * |
| **2** | Scottish Government 2021 | https://www.gov.scot/publications/compliance-self-isolation-quarantine-measures-literature-review/ | * | * | * | * | * |
| **3** | Smith et al 2020 | https://www.ncbi.nlm.nih.gov/pmc/articles/PMC7474581/ | * | * | * | * | * |
| **4** | Smith et al 2021 | https://www.bmj.com/content/372/bmj.n608 | * | * | * | * | * |
| **5** | Nuffield Trust 2021 | https://www.nuffieldtrust.org.uk/news-item/to-solitude-learning-from-other-countries-on-how-to-improve-compliance-with-self-isolation-1 | 1 | 0 | 1 | 0 | 1 |
| **6** | Institute of Applied Economics and Social Value | https://iaesv.our.dmu.ac.uk/2020/11/02/lockdown-compliance-in-the-uk/ | 1 | 0 | 1 | 0 | 1 |
| **7** | Fancourt et al 2020 | https://www.thelancet.com/journals/lancet/article/PIIS0140-6736(20)31690-1/fulltext | 1 | 0 | 1 | 0 | 1 |
| **8** | Blundell et al 2020 | https://onlinelibrary.wiley.com/doi/full/10.1111/1475-5890.12232 | 1 | 0 | 1 | 0 | 1 |
| **9** | Wright et al 2021 | https://www.thelancet.com/journals/lanepe/article/PIIS2666-7762(21)00038-7/fulltext | 1 | 0 | 1 | 0 | 1 |
| **10** | Patel et al 2021 | https://www.thelancet.com/journals/lanepe/article/PIIS2666-7762(21)00066-1/fulltext | * | * | * | * | * |
| **11** | Eraso & Hills 2021 | https://www.mdpi.com/1660-4601/18/13/7015 | * | * | * | * | * |
| **12** | Welsh Government 2020 | https://research.senedd.wales/research-articles/less-than-a-third-of-people-are-fully-self-isolating-during-the-coronavirus-pandemic-what-support-is-available-to-increase-compliance/ | * | * | * | * | * |
| **13** | ScotCen 2021 | https://dera.ioe.ac.uk/id/eprint/38459/1/covid-19-support-study-experiences-compliance-self-isolation.pdf | * | * | * | * | * |
| **14** | Scientific Pandemic Insights Group on Behaviours 2020 | https://assets.publishing.service.gov.uk/government/uploads/system/uploads/attachment_data/file/888750/7b._20.04.27_SPI-B_behavioural_science_notes_on_symptom_vs_test_based_approaches_S0260.pdf | 1 | 0 | 1 | 0 | 1 |

| | | | | | | | |
|---|---|---|---|---|---|---|---|
| **15** | Smith et al 2020 | https://www.sciencedirect.com/science/article/pii/S003335062030319X | * | * | * | * | * |
| **16** | Mills et al 2021 | https://academic.oup.com/policing/article/16/4/580/6486891 | 1 | 0 | 1 | 0 | 1 |
| **17** | Almayahi & Al Lamki 2022 | https://ejnpn.springeropen.com/articles/10.1186/s41983-022-00481-x | * | * | * | * | * |
| **18** | Eraso & Hills 2021 | https://journals.plos.org/plosone/article?id=10.1371/journal.pone.0256495 | 1 | 0 | 1 | 0 | 1 |
| **19** | Bodas & Peleg 2020 | https://www.healthaffairs.org/doi/10.1377/hlthaff.2020.00382 | * | * | * | * | * |
| **20** | Eraso & Hills 2021 | https://repository.londonmet.ac.uk/6861/1/ijerph-18-07015-v2.pdf | * | * | * | * | * |
| **21** | Zabadi et al 2021 | https://www.frontiersin.org/articles/10.3389/fpubh.2021.570242/full | 1 | 0 | 1 | 0 | 1 |
| **22** | Hills & Eraso 2021 | https://bmcpublichealth.biomedcentral.com/articles/10.1186/s12889-021-10379-7 | 1 | 0 | 1 | 0 | 1 |
| **23** | Office for National Statistics 2021 | https://www.ons.gov.uk/peoplepopulationandcommunity/healthandsocialcare/conditionsanddiseases/publications?page=5 | * | * | * | * | * |
| **24** | Centers for Disease Control and Prevention 2022 | https://www.cdc.gov/coronavirus/2019-ncov/php/contact-tracing/contact-tracing-plan/contact-tracing.html | 1 | 0 | 1 | 0 | 1 |
| **25** | Kalaij et al 2022 | https://jamsa.amsa-international.org/index.php/main/article/view/398 | 1 | 0 | 1 | 0 | 1 |
| **26** | Sopory et al 2022 | https://link.springer.com/article/10.1007/s10389-021-01544-8 | 1 | 0 | 1 | 0 | 1 |
| **27** | Welsh Government 2021 | https://phw.nhs.wales/publications/publications1/self-isolation-confidence-adherence-and-challenges-behavioural-insights-from-contacts-of-cases-of-covid-19-starting-and-completing-self-isolation-in-wales/ | * | * | * | * | * |
| **28** | Sopory et al 2022 | https://research.bangor.ac.uk/portal/files/37541291/2021_Quarantine_Acceptance.pdf | † | † | † | † | † |
| **29** | Science Media Centre 2020 | https://www.sciencemediacentre.org/expert-reaction-to-preprint-reporting-on-adherence-to-the-test-trace-and-isolate-system-in-the-uk/ | 1 | 0 | 1 | 0 | 1 |
| **30** | Lucas et al 2021 | https://royalsocietypublishing.org/doi/pdf/10.1098/rstb.2020.0270 | * | * | * | * | * |
| **31** | Smith et al 2020 | https://www.researchgate.net/publication/344205680_Factors_associated_with_adherence_to_self- | * | * | * | * | * |

| | | | | | | | |
|---|---|---|---|---|---|---|---|
| | | isolation_and_lockdown_measures_in_the_UK_a_cross-sectional_survey | | | | | |
| **32** | Welsh Government 2022 | https://www.gov.wales/sites/default/files/publications/2022-02/technical-advisory-group-reduction-in-isolation-period-supported-by-LFT-for-cases-of-COVID-19.pdf | * | * | * | * | * |
| **33** | Webster et al 2020 | https://eprints.whiterose.ac.uk/159125/ | * | * | * | * | * |
| **34** | Webster et al 2020 | https://eprints.whiterose.ac.uk/159125/5/1-s2.0-S0033350620300718-main.pdf | * | * | * | * | * |
| **35** | House of Lords 2021 | https://hansard.parliament.uk/Lords/2021-03-01/debates/B44440E9-7C5F-414A-AAE5-79821C80DD86/HealthProtection(CoronavirusRestrictions)(AllTiersAndSelf-Isolation)(England)(Amendment)Regulations2021 | 1 | 0 | 1 | 0 | 1 |
| **36** | Gasperini et al 2022 | https://www.cambridge.org/core/journals/epidemiology-and-infection/article/covid19-isolationquarantine-rules-in-home-care-patients/D0E817DC6C1C69794BB7DDBFBFE61FB3 | 1 | 0 | 1 | 1 | 0 |
| **37** | Cardwell et al 2021 | https://onlinelibrary.wiley.com/doi/10.1002/rmv.2244 | * | * | * | * | * |
| **38** | Schumpe et al 2022 | https://www.nature.com/articles/s41598-021-04703-9 | 1 | 0 | 1 | 0 | 1 |
| **39** | Institute for Government 2021 | https://www.instituteforgovernment.org.uk/article/comment/government-must-step-its-support-people-told-self-isolate | 1 | 0 | 1 | 0 | 1 |
| **40** | COVID-19 Evidence Network to support Decision Making 2021 | https://www.mcmasterforum.org/docs/default-source/product-documents/living-evidence-syntheses/covid-19-living-evidence-synthesis-13.1---appendix.pdf?sfvrsn=a0fbb76c_5 | 1 | 0 | 1 | 0 | 1 |
| **41** | Webster et al 2020 | https://pesquisa.bvsalud.org/global-literature-on-novel-coronavirus-2019-ncov/resource/pt/covidwho-625063 | * | * | * | * | * |
| **42** | Almaghrabi 2021 | https://www.dovepress.com/public-awareness-attitudes-and-adherence-to-covid-19-quarantine-and-is-peer-reviewed-fulltext-article-IJGM | * | * | * | * | * |
| **43** | Cabot & Bushnik 2022 | https://www150.statcan.gc.ca/n1/pub/82-003-x/2022009/article/00001-eng.htm | * | * | * | * | * |
| **44** | European Centre for Disease Prevention and Control 2020 | https://www.ecdc.europa.eu/sites/default/files/documents/covid-19-social-distancing-measuresg-guide-second-update.pdf | 1 | 0 | 1 | 0 | 1 |
| **45** | Patel et al 2021 | https://www.pure.ed.ac.uk/ws/portalfiles/portal/212199743/Maximising_public_adherence_to_COVID_19_self_isolation_i | * | * | * | * | * |

| | | | | | | | |
|---|---|---|---|---|---|---|---|
| | | n_Europe.pdf | | | | | |
| **46** | Schnyer et al 2021 | https://smw.ch/index.php/smw/announcement/view/40 | 1 | 0 | 1 | 0 | 1 |
| **47** | Wright et al 2021 | https://discovery.ucl.ac.uk/id/eprint/10128631/1/1-s2.0-S2666776221000387-main.pdf | † | † | † | † | † |
| **48** | Newman University 2022 | https://www.newman.ac.uk/wp-content/uploads/sites/10/2022/04/Covid-19-Resilience-Document-May-2022-Final.pdf | 1 | 0 | 1 | 0 | 1 |
| **49** | Kucharski 2020 [tweet] | https://twitter.com/adamjkucharski/status/1309145690992455682 | 1 | 0 | 1 | 0 | 1 |
| **50** | Health Information and Quality Authority 2021 | https://www.hiqa.ie/sites/default/files/2021-02/Measures-to-support-self-isolation-and-ROM_Protocol.pdf | 1 | 0 | 1 | 0 | 1 |
| **51** | Webster et al 2020 | https://europepmc.org/article/MED/32334182 | * | * | * | * | * |
| **52** | Guernsey Press 2021 | https://guernseypress.com/news/uk-news/2021/01/13/lockdown-compliance-high-but-concerns-about-adherence-to-self-isolation-study/ | 1 | 0 | 1 | 0 | 1 |
| **53** | Ukonu et al 2021 | https://journals.sagepub.com/doi/full/10.1177/21582440211047243 | 1 | 0 | 1 | 0 | 1 |
| **54** | Bodas & Peleg 2020 | https://www.semanticscholar.org/paper/Self-Isolation-Compliance-In-The-COVID-19-Era-By-A-Bodas-Peleg/f6e2216725013f8d004e69721164cf8d24eb1bf7 | * | * | * | * | * |
| **55** | Dukhi et al 2021 | https://openpublichealthjournal.com/VOLUME/14/PAGE/45/FULLTEXT/ | 1 | 0 | 1 | 0 | 1 |
| **56** | Jurblum et al 2020 | https://www1.racgp.org.au/ajgp/2020/december/psychological-consequences-of-social-isolation-and | 1 | 0 | 1 | 0 | 1 |
| **57** | Washington Post 2020 | https://www.washingtonpost.com/world/2020/09/25/coronavirus-self-isolation-adherence-paid-leave/ | 1 | 0 | 1 | 0 | 1 |
| **58** | SBS News 2020 | https://www.sbs.com.au/news/article/simply-unacceptable-one-in-four-victorians-with-coronavirus-not-at-home-when-door-knocked/5wz7jxj60 | 1 | 0 | 1 | 0 | 1 |
| **59** | Rifa'i et al 2022 | https://ejurnal.ung.ac.id/index.php/gojhes/article/view/13577 | 1 | 0 | 1 | 1 | 0 |
| **60** | Gasperini et al 2022 | https://www.scienceopen.com/document_file/775aa2ad-8ea0-4e85-b05d-a9197d66f924/PubMedCentral/775aa2ad-8ea0-4e85-b05d-a9197d66f924.pdf | † | † | † | † | † |
| **61** | Van den Bergh & Hoorens 2020 | https://en.bfp-fbp.be/report-quarantine-adherence | 1 | 0 | 1 | 0 | 1 |

| | | | | | | | |
|---|---|---|---|---|---|---|---|
| **62** | Webster et al 2020 | https://www.med.uminho.pt/pt/covid19/Sade%20Mental/Webster%202020%20How%20to%20improve%20adherence%20with%20quarantine.pdf | * | * | * | * | * |
| **63** | National Mental Health Commission | https://www.google.co.uk/url?sa=t&rct=j&q=&esrc=s&source=web&cd=&ved=2ahUKEwi7u4qur4T_AhXMS8AKHUiDB9c4PBAWegQIAhAB&url=https%3A%2F%2Fwww.health.gov.au%2Fsites%2Fdefault%2Ffiles%2Fdocuments%2F2020%2F12%2Fcoronavirus-covid-19-advice-on-mental-health-screening-assessment-and-support-during-covid-19-quarantine.doc&usg=AOvVaw2OyMljg_fLGA2YlVAYeQ0m | 1 | 0 | 1 | 0 | 1 |
| **64** | Zimmermann et al 2022 | https://www.ijhpm.com/article_4035.html | 1 | 0 | 1 | 0 | 1 |
| **65** | Smith et al 2020 | https://researchportal.ukhsa.gov.uk/ws/portalfiles/portal/42932482/Smith2020FactorsAssociatedWithAdherenceToSelf_IsolationAndLockdownMeasuresInTheUKPublicHealth.pdf | * | * | * | * | * |
| **66** | Mansdorf 2020 | https://reliefweb.int/report/world/enforcing-compliance-covid-19-pandemic-restrictions-psychological-aspects-national | 1 | 0 | 1 | 0 | 1 |
| **67** | Maragakis 2020 | https://www.hopkinsmedicine.org/health/conditions-and-diseases/coronavirus/coronavirus-social-distancing-and-self-quarantine | 1 | 0 | 1 | 0 | 1 |
| **68** | Northern Echo 2021 | https://www.thenorthernecho.co.uk/news/national/19007395.lockdown-compliance-high-concerns-adherence-self-isolation---study/ | 1 | 0 | 1 | 0 | 1 |
| **69** | King's College London 2020 | https://www.kcl.ac.uk/news/adherence-to-quarantine-during-covid-19-pandemic | 1 | 0 | 1 | 0 | 1 |
| **70** | Steens et al 2020 | https://www.eurosurveillance.org/content/10.2807/1560-7917.ES.2020.25.37.2001607 | * | * | * | * | * |
| **71** | UK Government 2020 | https://www.legislation.gov.uk/uksi/2020/1045/made | 1 | 0 | 1 | 0 | 1 |
| **72** | Guillen 2021 | https://www.scirp.org/pdf/ajc_2021081310221813.pdf | 1 | 0 | 1 | 0 | 1 |
| **73** | Government of the Czech Republic 2022 | https://www.vlada.cz/en/media-centrum/aktualne/measures-adopted-by-the-czech-government-against-coronavirus-180545/ | 1 | 0 | 1 | 0 | 1 |
| **74** | Centrally Funded Technical Institutes 2022 | https://nitttrbpl.ac.in/files/Covid%20E%20Book_21_03_22.pdf | 1 | 0 | 1 | 0 | 1 |
| **75** | Republic of Slovenia 2022 | https://www.gov.si/en/news/?date=&nrOfItems=50&tag%5B0%5D=554 | 1 | 0 | 1 | 0 | 1 |

| | | | | | | | |
|---|---|---|---|---|---|---|---|
| **76** | Not reported | https://api.portal.idealagent.com/IDtrack?rackid=K35v506&FilesData=Mcq_In_Infection_Control_Mahesy.pdf | 1 | 1 | 0 | 0 | 0 |
| **77** | Ulijaszek 2017 | https://books.google.co.uk/books?id=-y47DwAAQBAJ&pg=PA140&lpg=PA140&dq=self-isolation+quarantine+adherence+compliance&source=bl&ots=dwEIDy61vX&sig=ACfU3U3S-QpecNhefl6utiQUTJ2-AE-nng&hl=en&sa=X&ved=2ahUKEwjI5qbRsIT_AhUFhlwKHXHmAIw4RhDoAXoECAQQAw#v=onepage&q=self-isolation%20quarantine%20adherence%20compliance&f=false | 1 | 0 | 1 | 0 | 1 |
| **78** | Sabate et al 2022 | https://books.google.co.uk/books?id=ZjZuEAAAQBAJ&pg=PA88&lpg=PA88&dq=self-isolation+quarantine+adherence+compliance&source=bl&ots=fUbN4S_aDZ&sig=ACfU3U3JD2DUBe2OxTSWku-zFGv4LkzYEw&hl=en&sa=X&ved=2ahUKEwjI5qbRsIT_AhUFhlwKHXHmAIw4RhDoAXoECAMQAw#v=onepage&q=self-isolation%20quarantine%20adherence%20compliance&f=false | 1 | 0 | 1 | 0 | 1 |
| **79** | Pashoja (ed) 2021 | https://books.google.co.uk/books?id=7M6aEAAAQBAJ&pg=PA51&lpg=PA51&dq=self-isolation+quarantine+adherence+compliance&source=bl&ots=y8-IhEaYpV&sig=ACfU3U3wOyzhUHf_ekzy9alcJyyftf9OvQ&hl=en&sa=X&ved=2ahUKEwjI5qbRsIT_AhUFhlwKHXHmAIw4RhDoAXoECCAQAw#v=onepage&q=self-isolation%20quarantine%20adherence%20compliance&f=false | 1 | 0 | 1 | 0 | 1 |
| **80** | Kowalski et al. 2020 | https://www.ncbi.nlm.nih.gov/pmc/articles/PMC7590639/ | * | * | * | * | * |
| **81** | Wright et al 2021 | https://www.ncbi.nlm.nih.gov/pmc/articles/PMC7907734/ | † | † | † | † | † |
| **82** | Eraso & Hills 2021 | https://www.ncbi.nlm.nih.gov/pmc/articles/PMC8297259/ | * | * | * | * | * |
| **83** | Webster et al 2020 | https://www.ncbi.nlm.nih.gov/pmc/articles/PMC7194967/ | * | * | * | * | * |
| **84** | Rojek et al 2022 | https://www.ncbi.nlm.nih.gov/pmc/articles/PMC9819452/ | 1 | 0 | 1 | 0 | 1 |
| **85** | Al Zabadi et al 2021 | https://www.ncbi.nlm.nih.gov/pmc/articles/PMC7960769/ | * | * | * | * | * |

| | | | | | | | |
|---|---|---|---|---|---|---|---|
| **86** | Steens et al 2020 | https://www.ncbi.nlm.nih.gov/pmc/articles/PMC7502884/ | * | * | * | * | * |
| **87** | Shushtari et al 2021 | https://www.ncbi.nlm.nih.gov/pmc/articles/PMC8721272/ | 1 | 0 | 1 | 0 | 1 |
| **88** | Almaghrabi 2021 | https://www.ncbi.nlm.nih.gov/pmc/articles/PMC8364964/ | * | * | * | * | * |
| **89** | Lucas et al 2021 | https://www.ncbi.nlm.nih.gov/pmc/articles/PMC8165588/ | * | * | * | * | * |
| **90** | Book et al 2022 | https://www.ncbi.nlm.nih.gov/pmc/articles/PMC9638749/ | * | * | * | * | * |
| **91** | Yu et al 2022 | https://www.ncbi.nlm.nih.gov/pmc/articles/PMC9119710/ | 1 | 0 | 1 | 0 | 1 |
| **92** | Patel et al 2021 | https://www.ncbi.nlm.nih.gov/pmc/articles/PMC7981224/ | * | * | * | * | * |
| **93** | Scientific Pandemic Insights Group on Behaviours | https://assets.publishing.service.gov.uk/government/uploads/system/uploads/attachment_data/file/966941/S0899_SPI-B_Behavioural_effects_of_reducing_duration_of_quarantine_for_contacts.pdf | 1 | 0 | 1 | 0 | 1 |
| **94** | Galende et al 2022 | https://www.ncbi.nlm.nih.gov/pmc/articles/PMC9025158/ | 1 | 0 | 1 | 0 | 1 |
| **95** | Scientific Advisory Group for Emergencies 2022 | https://www.gov.uk/government/publications/tfms-behavioural-paper-supporting-the-consensus-statement-on-mass-testing-27-august-2020/tfms-behavioural-paper-supporting-the-consensus-statement-on-mass-testing-27-august-2020 | * | * | * | * | * |
| **96** | Scottish Government 2022 | https://www.gov.scot/binaries/content/documents/govscot/publications/research-and-analysis/2021/08/covid-19-support-study-experiences-compliance-self-isolation-research-findings/documents/research-findings-no-147-2021-covid-19-support-study-experiences-compliance-self-isolation/research-findings-no-147-2021-covid-19-support-study-experiences-compliance-self-isolation/govscot%3Adocument/research-findings-no-147-2021-covid-19-support-study-experiences-compliance-self-isolation.pdf | * | * | * | * | * |
| **97** | Tarr et al 2022 | https://www.google.co.uk/url?sa=t&rct=j&q=&esrc=s&source=web&cd=&ved=2ahUKEwjY9eqKsoT_AhUVOcAKHUEcCvA4HhAWegQIGhAB&url=https%3A%2F%2Fjournals.plos.org%2Fplosone%2Farticle%2Ffile%3Fid%3D10.1371%2Fjournal.pone.0267261%26type%3Dprintable&usg=AOvVaw1RG | * | * | * | * | * |

| | | | | | | | |
|---|---|---|---|---|---|---|---|
| | | gALuBV_1-tuKayKhLlx | | | | | |
| **98** | Reicher et al 2021 | https://blogs.bmj.com/bmj/2021/04/05/why-contrasting-figures-on-adherence-to-self-isolation-show-that-support-to-self-isolate-is-even-more-important-than-we-previously-realised/ | 1 | 0 | 1 | 0 | 1 |
| **99** | Almayahi & Al Lamki 2022 | https://www.ncbi.nlm.nih.gov/pmc/articles/PMC9079213/ | * | * | * | * | * |
| **100** | de Noronha et al 2022 | https://www.ncbi.nlm.nih.gov/pmc/articles/PMC9271418/ | 1 | 0 | 1 | 0 | 1 |
| | | TOTAL: | 54 | 1 | 53 | 2 | 51 |

* Study had already been identified in a previous search (database or other grey literature) and therefore was not screened again.
† Found earlier in google search and therefore was not screened again.

#### Supplement S4. Adapted Joanna Briggs Institute Prevalence Critical Appraisal Tool

Amendments to the tool:

1. We excluded the question asking "are all important confounding factors/subgroups/differences identified and accounted for?" as this was not applicable where only a prevalence rating and no further analyses of associations were investigated.
2. We assigned scores to the response options for each question "yes" (1), "no" (0), "unclear" (0) or "not applicable" (1), and allowed an extra response option ("strong yes", scored as 2) for the question asking "were study participants recruited in an appropriate way?".
3. Scores were summed to give a total score out of 10. Studies were then categorised as having low risk of bias (scores 9 to 10), some concerns (scores 7 to 8), high risk of bias (scores 5 to 6), or very high risk of bias (scores 0 to 4).

Tool used:

| | **Question** | **Description** | **Response options (score)** | **Rater rules of thumb** |
|---|---|---|---|---|
| **1** | Was the sample representative of the target population? | This question relies upon knowledge of the characteristics of broader the population of interest. If the study is of women with breast cancer, knowledge of at least the characteristics, demographics, and medical history is needed. The term "target population" should not be taken to infer every individual from everywhere or with similar disease or exposure characteristics. Instead, give consideration to specific population characteristics in the study, including age range, gender, morbidities, medications, and other potentially influential factors. For example, a sample may not be representative of the target population if a certain group has been used (such as those working for one organisation, or one profession) and the results then inferred to the target population (i.e. working adults). | Y (1), N (0), U (0), N/A (1) | |

| | | | | |
|---|---|---|---|---|
| **2** | Were study participants recruited in an appropriate way? | Recruitment is the calling or advertising strategy for gaining interest in the study, and is not the same as sampling. Studies may report random sampling from a population, and the methods section should report how sampling was performed. What source of data were study participants recruited from? Was the sampling frame appropriate? For example, census data is a good example of appropriate recruitment as a good census will identify everybody. Was everybody included who should have been included? Were any groups of persons excluded? Was the whole population of interest surveyed? If not, was random sampling from a defined subset of the population employed? Was stratified random sampling with eligibility criteria used to ensure the sample was representative of the population that the researchers were generalizing to? | YY (2), Y (1), N (0), U (0), N/A (1) | Random sampling or all eligible invited = YY (2). Quota sampling = Y (1). Convenience sampling = N (0) |
| **3** | Was the sample size adequate? | An adequate sample size is important to ensure good precision of the final estimate. Ideally we are looking for evidence that the authors conducted a sample size calculation to determine an adequate sample size. This will estimate how many subjects are needed to produce a reliable estimate of the measure(s) of interest. For conditions with a low prevalence, a larger sample size is needed. Also consider sample sizes for subgroup (or characteristics) analyses, and whether these are appropriate. Sometimes, the study will be large enough (as in large national surveys) whereby a sample size calculation is not required. In these cases, sample size can be considered adequate. When there is no sample size calculation and it is not a large national survey, the reviewers may consider conducting their own sample size analysis using the following formula (24,25):<br>$n = Z^2P(1-P) / d^2$<br>Where:<br>n= sample size<br>Z= Z statistic for a level of confidence<br>P= Expected prevalence or proportion (in proportion of one; if 20%, P= 0.2)<br>d= precision (in proportion of one; if 5%, d= 0.05) | Y (1), N (0), U (0), N/A (1) | Z =1.96, as for 95% CI. P=50%. D=0.05. GIVES n=385. (((1.96*1.96)*0.5)*0.5) / (0.05*0.05) = 0.9604 / 0.0025 = 384.16<br><br>If a study reports two or more prevalence rates from different sample sizes, use the largest group to answer this question. |
| **4** | Were the study subjects and setting described in detail? | Certain diseases or conditions vary in prevalence across different geographic regions and populations (e.g., women vs. men, socio-demographic variables between countries). Has the study sample been described in sufficient detail so that other researchers can determine if it is comparable to the population of interest to them? | Y (1), N (0), U (0), N/A (1) | Age, gender, region |

| | | | | |
|---|---|---|---|---|
| **5** | Is the data analysis conducted with sufficient coverage of the identified sample? [LS - missing data] | A large number of dropouts, refusals or "not founds" amongst selected subjects may diminish a study's validity, as can low response rates for survey studies.<br>• Did the authors describe the reasons for non-response and compare persons in the study to those not in the study, particularly with regards to their socio-demographic characteristics?<br>• Could the not-responders have led to an underestimate of prevalence of the disease or condition under investigation?<br>• If reasons for non-response appear to be unrelated to the outcome measured and the characteristics of non-responders are comparable to those in the study, the researchers may be able to justify a more modest response rate.<br>• Did the means of assessment or measurement negatively affect the response rate (measurement should be easily accessible, conveniently timed for participants, acceptable in length, and suitable in content). | Y (1), N (0), U (0), N/A (1) | Less 5% missing data = Y. (Question 1 covers representativeness of sample and dropouts/refusals etc.) |
| **6** | Were objective, standard criteria used for measurement of the condition? | Here we are looking for measurement or classification bias. Many health problems are not easily diagnosed or defined and some measures may not be capable of including or excluding appropriate levels or stages of the health problem. If the outcomes were assessed based on existing definitions or diagnostic criteria, then the answer to this question is likely to be yes. If the outcomes were assessed using observer reported, or self-reported scales, the risk of over- or under-reporting is increased, and objectivity is compromised. Importantly, determine if the measurement tools used were validated instruments as this has a significant impact on outcome assessment validity. | Y (1), N (0), U (0), N/A (1) | Self-/observer-reported. Subjective e.g., "adherence to isolation" = N. Objective, e.g., number of times left home = Y. Measure not given = U. |
| **7** | Was the condition [ADHERENCE] measured reliably? | Considerable judgment is required to determine the presence of some health outcomes. Having established the objectivity of the outcome measurement instrument (see item 6 of this scale), it is important to establish how the measurement was conducted. Were those involved in collecting data trained or educated in the use of the instrument/s? If there was more than one data collector, were they similar in terms of level of education, clinical or research experience, or level of responsibility in the piece of research being appraised?<br>• Has the researcher justified the methods chosen?<br>• Has the researcher made the methods explicit? (For interview method, how were interviews conducted?) | Y (1), N (0), U (0), N/A (1) | Self-reported = N (as each participant could have a different interpretation of the measure). Different researchers = N. Different researchers, but they are trained = Y |

| | | | | |
|---|---|---|---|---|
| **8** | Was there appropriate statistical analysis? | As with any consideration of statistical analysis, consideration should be given to whether there was a more appropriate alternate statistical method that could have been used. The methods section should be detailed enough for reviewers to identify the analytical technique used and how specific variables were measured. Additionally, it is also important to assess the appropriateness of the analytical strategy in terms of the assumptions associated with the approach as differing methods of analysis are based on differing assumptions about the data and how it will respond. Prevalence rates found in studies only provide estimates of the true prevalence of a problem in the larger population. Since some subgroups are very small, 95% confidence intervals are usually given. | Y (1), N (0), U (0), N/A (1) | Reported multiple outcome measurements or rates from multiple different subgroups without justification = N. |
| **10** | Were subpopulations identified using objective criteria? | Objective criteria should also be used where possible to identify subgroups (refer to question 6). | Y (1), N (0), U (0), N/A (1) | |

Abbreviations: YY = strong yes, Y = yes, N = no, U = unclear, N/A = not applicable, CI = confidence interval.

#### Supplement S5. Full details of the analysis conducted for each outcome

For definitions of self-isolation, all studies were analysed together. Details of behaviours required during self-isolation were synthesised. There was no standardised metric for this outcome.

For measures of self-isolation, we first separated studies by mode of data collection. Details of the measure were synthesised only for studies using self-report measures. There was no standardised metric for this outcome.

For rates of adherence to self-isolation, studies were grouped based on reasons why participants were in self-isolation (e.g., due to a positive test, or for travel) as rates may have differed by reason for self-isolation, taking into account risk of bias rating. Some studies reported multiple rates of adherence; these were often different behaviours, such as staying at home, or wearing a face covering. Where studies reported multiple rates of adherence to different aspects of self-isolation, we chose the statistic that was most in line with "not leaving home" and selected the item that was phrased in such a way to be most permissible to report breaking the rules – that is, the item least likely to be affected by social desirability. [5] Prevalence estimates selected for synthesis are shown in Supplementary Table S8. For this outcome, we reported percentages as the standardised metric and drew a Forest plot of adherence rates. To informally investigate heterogeneity, as no meta-analysis was planned, we structured the reporting of prevalence rates by reason for self-isolation and risk of bias rating. Confidence intervals were used to assess the certainty of evidence. No transformations were applied for results reported in different formats (e.g., mean).

For analyses of factors associated with adherence to self-isolation, studies were grouped based on reasons why participants were isolating to allow patterns in associations to be identified. For this outcome, the standardised metric was the direction of the effect. To informally investigate heterogeneity, we structured the reporting of directions of effect by reason for self-isolation and risk of bias rating. Consistency of effects across studies was used to assess the certainty of synthesis findings. Sometimes, studies investigated factors associated with adherence to self-isolation in multiple sub-samples who were isolating for different reasons (e.g., because they were a COVID-19 case, or because they were a contact). We analysed these separately. Where studies reported multiple analyses, we synthesised the most rigorous analysis conducted in each group (see Supplementary Table S8). We extracted all factors associated with self-isolation, noting whether studies reported they were positively associated with adherence to self-isolation, negatively associated, had mixed results (where studies analysed the same factor across multiple waves with different results), or showed no evidence for an effect. Results were tabulated, accounting for risk of bias rating. Where studies grouped participants and the groups overlapped, we tabulated these results together (one study analysing COVID-19 cases and people returning from travel together, and another analysing cases, contacts and people returning from travel together). Where only one study investigated an outcome, findings were removed from the summary and not included in the synthesis. Where only one analysis investigated a factor and its association with adherence to self-isolation, these were removed from the summary table. There were some cases where removing studies because of their outcome left only one analysis that investigated a factor in the summary table; these were also removed from the table. Risk of bias rating and number of studies investigating a factor were taken into account when summarising the strength of evidence for associations with adherence to self-isolation.

#### Supplement S6. Full details of risk of bias ratings for studies

##### Prevalence estimates, assessed using amended Joanna Briggs Institute Prevalence Critical Appraisal Tool

| Citation | Q1 | | Q2 | | Q3 | | Q4 | | Q5 | | Q6 | | Q7 | | Q8 | | Q10 | | Overall | |
|---|---|---|---|---|---|---|---|---|---|---|---|---|---|---|---|---|---|---|---|---|
| | Ans | Score | Ans | Score | Ans | Score | Ans | Score | Ans | Score | Ans | Score | Ans | Score | Ans | Score | Answer | Score | Total | Rating |
| **Ali-Saleh & Obeid 2022 [6]** | U | 0 | N | 0 | N | 0 | N | 0 | Y | 1 | N | 0 | N | 0 | N | 0 | N/A | 1 | 2 | Very high |
| **Almaghrabi 2021 [7]** | U | 0 | U | 0 | N | 0 | N | 0 | U | 0 | N | 0 | N | 0 | Y | 1 | N/A | 1 | 2 | Very high |
| **Almayahi & Al Lamki 2022 [8]** | U | 0 | YY | 2 | N | 0 | Y | 1 | Y | 1 | Y | 1 | N | 0 | Y | 1 | N/A | 1 | 7 | Some concerns |
| **Aslaner et al 2022 [9]** | U | 0 | YY | 2 | Y | 1 | Y | 1 | Y | 1 | N | 0 | N | 0 | Y | 1 | N/A | 1 | 7 | Some concerns |
| **Bannour et al 2021 [10]** | U | 0 | YY | 2 | N | 0 | Y | 1 | Y | 1 | N | 0 | N | 0 | Y | 1 | N/A | 1 | 6 | High |
| **Bara'a et al 2021 [11]** | U | 0 | U | 0 | Y | 1 | N | 0 | Y | 1 | N | 0 | N | 0 | Y | 1 | N/A | 1 | 4 | Very high |
| **Carlsen et al 2020 [12]** | U | 0 | U | 0 | Y | 1 | N | 0 | Y | 1 | N | 0 | N | 0 | Y | 1 | N | 0 | 3 | Very high |
| **Domenghino et al 2022 [13]** | U | 0 | YY | 2 | Y | 1 | Y | 1 | Y | 1 | N | 0 | N | 0 | Y | 1 | N/A | 1 | 7 | Some concerns |
| **Dowthwaite et al 2021 [14, 15]** | U | 0 | Y | 1 | N | 0 | N | 0 | N | 0 | N | 0 | N | 0 | N | 0 | N | 0 | 1 | Very high |
| **Elaraby et al 2022 [16]** | U | 0 | N | 0 | N | 0 | N | 0 | U | 0 | N | 0 | N | 0 | Y | 1 | N/A | 1 | 2 | Very high |
| **Enticott et al 2021 [17]** | U | 0 | Y | 1 | Y | 1 | N | 0 | N | 0 | N | 0 | N | 0 | N | 0 | Y | 1 | 3 | Very high |
| **Eraso & Hills 2021 [18]** | N | 0 | N | 0 | N | 0 | Y | 1 | Y | 1 | Y | 1 | N | 0 | Y | 1 | U | 0 | 4 | Very high |
| **Eslamzadeh et al 2022 [19]** | U | 0 | N | 0 | N | 0 | Y | 1 | U | 0 | N | 0 | N | 0 | N | 0 | N/A | 1 | 2 | Very high |
| **Farooq et al 2021 [20]** | U | 0 | YY | 2 | N | 0 | Y | 1 | U | 0 | U | 0 | N | 0 | N | 0 | N | 0 | 3 | Very high |
| **Foroozanfar et al 2020 [21]** | U | 0 | YY | 2 | N | 0 | Y | 1 | Y | 1 | U | 0 | N | 0 | Y | 1 | N/A | 1 | 6 | High |
| **Fuchs et al 2021 [22]** | N | 0 | Y | 2 | Y | 1 | N | 0 | Y | 1 | Y | 1 | Y | 1 | Y | 1 | N/A | 1 | 8 | Some concerns |
| **Gasperini et al 2022 [23]** | U | 0 | U | 0 | N | 0 | N | 0 | Y | 1 | N | 0 | U | 0 | N | 1 | Y | 1 | 3 | Very high |

| | | | | | | | | | | | | | | | | | | | | |
|---|---|---|---|---|---|---|---|---|---|---|---|---|---|---|---|---|---|---|---|---|
| **Hood et al 2022 [24] [case interview]** | N | 0 | Y | 1 | Y | 1 | N | 0 | N | 0 | U | 0 | Y | 1 | N | 0 | N/A | 1 | 4 | Very high |
| **Hood et al 2022 [24] [survey]** | N | 0 | YY | 2 | N | 0 | N | 0 | U | 0 | Y | 1 | N | 0 | Y | 1 | N/A | 1 | 5 | High |
| **Kyle et al 2021 [25, 26]** | Y | 1 | Y | 1 | Y | 1 | Y | 1 | Y | 1 | Y | 1 | N | 0 | Y | 1 | N/A | 1 | 8 | Some concerns |
| **Li et al 2021 [27]** | U | 0 | U | 0 | N | 0 | Y | 1 | Y | 1 | Y | 1 | N | 0 | Y | 1 | N/A | 1 | 5 | High |
| **Lou et al 2020 [28]** | U | 0 | U | 0 | Y | 1 | Y | 1 | Y | 1 | N | 0 | N | 0 | Y | 1 | N/A | 1 | 5 | High |
| **Martin et al 2021 [29]** | U | 0 | YY | 2 | N | 0 | N | 0 | Y | 1 | Y | 1 | N | 0 | Y | 1 | N/A | 1 | 6 | High |
| **Mijovic et al 2021 [30]** | U | 0 | U | 0 | N | 0 | N | 0 | Y | 1 | Y | 1 | N | 0 | Y | 1 | N/A | 1 | 4 | Very high |
| **Office for National Statistics 2021, 2022 [31-44]** | Y | 1 | YY | 2 | Y | 1 | N | 0 | Y | 1 | Y | 1 | N | 0 | Y | 1 | N/A | 1 | 8 | Some concerns |
| **Office for National Statistics 2021 [45-50]** | Y | 1 | YY | 2 | Y | 1 | N | 0 | Y | 1 | Y | 1 | N | 0 | Y | 1 | N/A | 1 | 8 | Some concerns |
| **Office for National Statistics 2021 [51]** | Y | 1 | YY | 2 | Y | 1 | N | 0 | Y | 1 | Y | 1 | N | 0 | Y | 1 | Y | 1 | 8 | Some concerns |
| **Pinheiro et al 2022 [52, 53]** | U | 0 | U | 0 | N | 0 | Y | 1 | Y | 1 | U | 0 | N | 0 | N | 0 | Y | 1 | 3 | Very high |
| **Rifa'i et al 2022 [54]** | N | 0 | N | 0 | N | 0 | Y | 1 | Y | 1 | U | 0 | N | 0 | Y | 1 | N/A | 1 | 4 | Very high |
| **Ripon et al 2020 [55]** | U | 0 | N | 0 | Y | 1 | N | 0 | Y | 1 | U | 0 | N | 0 | N | 0 | N/A | 1 | 3 | Very high |
| **Robin et al 2022 [56]** | U | 0 | YY | 2 | N | 0 | U | 0 | Y | 1 | Y | 1 | N | 0 | Y | 1 | N/A | 1 | 6 | High |
| **Rosca et al 2020 [57]** | N | 0 | YY | 2 | N | 0 | N | 0 | N | 0 | U | 0 | N | 0 | N | 0 | Y | 1 | 3 | Very high |
| **Rubio et al 2021 [58] [survey]** | U | 0 | YY | 2 | N | 0 | N | 0 | Y | 1 | Y | 1 | N | 0 | Y | 1 | N/A | 1 | 6 | High |
| **Rubio et al 2021 [58] [random** | U | 0 | YY | 2 | N | 0 | N | 0 | N | 0 | Y | 1 | Y | 1 | Y | 1 | N/A | 1 | 6 | High |

| | | | | | | | | | | | | | | | | | | | | |
|---|---|---|---|---|---|---|---|---|---|---|---|---|---|---|---|---|---|---|---|---|
| **home visits and calls]** | | | | | | | | | | | | | | | | | | | | |
| **Ryu et al 2022 [59]** | Y | 1 | YY | 2 | Y | 1 | N | 0 | Y | 1 | Y | 1 | Y | 1 | Y | 1 | N/A | 1 | 9 | Low |
| **Sahin et al 2022 [60] – cold symptoms** | N | 0 | N | 0 | Y | 1 | N | 0 | Y | 1 | N | 0 | N | 0 | Y | 1 | N/A | 1 | 4 | Very high |
| **Sahin et al 2022 [60] – travel** | U | 0 | N | 0 | N | 0 | N | 0 | Y | 1 | N | 0 | N | 0 | Y | 1 | N/A | 1 | 3 | Very high |
| **Scottish Government 2021 [61, 62]** | N | 0 | Y | 1 | Y | 1 | N | 0 | U | 0 | Y | 1 | N | 0 | N | 0 | Y | 1 | 4 | Very high |
| **Senol & Avci 2022 [63]** | U | 0 | U | 0 | Y | 1 | Y | 1 | Y | 1 | N | 0 | N | 0 | Y | 1 | N/A | 1 | 5 | High |
| **Shewasinad Yehualashet et al 2021 [64]** | U | 0 | YY | 2 | Y | 1 | Y | 1 | Y | 1 | N | 0 | N | 0 | Y | 1 | N/A | 1 | 7 | Some concerns |
| **Smith et al 2020 [65]** | U | 0 | Y | 1 | N | 0 | N | 0 | Y | 1 | Y | 1 | N | 0 | Y | 1 | N/A | 1 | 5 | High |
| **Smith et al 2021 [66]** | U | 0 | Y | 1 | Y | 1 | Y | 1 | Y | 1 | Y | 1 | N | 0 | N | 0 | Y | 1 | 6 | High |
| **Steens et al 2020 [67]** | U | 0 | YY | 2 | Y | 1 | N | 0 | Y | 1 | U | 0 | N | 0 | N | 0 | Y | 1 | 5 | High |
| **Tseng et al 2021 [68]** | U | 0 | YY | 2 | N | 0 | Y | 1 | Y | 1 | Y | 1 | N | 0 | Y | 1 | N/A | 1 | 7 | Some concerns |
| **Yang et al 2022 [69]** | U | 0 | YY | 2 | N | 0 | Y | 1 | U | 0 | Y | 1 | N | 0 | Y | 1 | N/A | 1 | 6 | High |

Abbreviations: ans = answer, N = no, N/A = not applicable, U = unclear, Y = yes, YY = strong yes.

#### Analyses of associations, assessed using ROBINS-E tool

| Citation | Domain 1 | Domain 2 | Domain 3 | Domain 4 | Domain 5 | Domain 6 | Domain 7 | Overall |
|---|---|---|---|---|---|---|---|---|
| **Ali-Saleh & Obeid 2022 [6]** | High | Some concerns | Some concerns | Low | Low | Some concerns | High | High |
| **Carlsen et al 2020 [12]** | Low | Low | Low | Low | Low | High | High | High |
| **Domenghino et al 2022 [13]** | Low | Some concerns | High | Low | Very high | Some concerns | High | Very high |
| **Enticott et al 2021 [17]** | Low | Low | Some concerns | Low | Very high | Some concerns | Low | Very high |
| **Eraso & Hills 2021 [18]** | Low | Some concerns | Some concerns | Low | Low | Some concerns | Low | High |
| **Eslamzadeh et al 2022 [19]** | Low | Low | Some concerns | Low | High | Some concerns | High | High |
| **Farooq et al 2021 [20]** | High | Low | Some concerns | Low | High | Some concerns | High | Very high |
| **Foroozanfar et al 2020 [21]** | High | Low | Some concerns | High | Low | Some concerns | High | Very high |
| **Fuchs et al 2021 [22]** | Low | Low | Low | Low | Some concerns | Low | Low | Some concerns |
| **Gasperini et al 2022 [23]** | High | Low | Low | Low | Low | High | Low | High |
| **Kriens 2022 [70]** | Some concerns | Some concerns | Some concerns | Low | High | Some concerns | Low | High |
| **Kyle et al 2021 [25, 26]** | Low | Some concerns | Some concerns | Low | Low | Some concerns | Low | High |
| **Lou et al 2020 [28]** | Low | Low | Low | Low | Low | Some concerns | Low | Some concerns |
| **Martin et al 2021 [29] *** | High | Some concerns | Some concerns | High | Low | Some concerns | High | Very high |
| **Mijovic et al 2021 [30]** | High | Low | Some concerns | Low | Low | Some concerns | Low | High |
| **Office for National Statistics 2021, 2022 [31-44]** | High | Low | Low | Low | Low | Some concerns | High | High |
| **Office for National Statistics 2021 [45-50]** | High | Low | Low | Low | Low | Some concerns | High | High |
| **Office for National Statistics 2021 [51]** | High | Low | Low | Low | Low | Some concerns | High | High |
| **Robin et al 2022 [56]** | High | Some concerns | High | Low | Some concerns | Some concerns | High | Very high |
| **Ryu et al 2022 [59]** | High | Low | Low | Low | Low | Low | High | High |
| **Sahin et al 2022 [60]** | High | Low | Some concerns | Low | Low | Some concerns | High | High |
| **Scottish Government 2021 [61, 62]** | High | Some concerns | Some concerns | Low | High | Some concerns | High | Very high |
| **Smith et al 2020 [65]** | Low | Low | Low | Low | High | Some concerns | Low | High |
| **Smith et al 2021 [66]** | Low | Low | Low | Low | High | Some concerns | High | High |
| **Steens et al 2020 [67]** | High | Low | Low | Low | High | High | High | Very high |

*Assessed using ROBINS-I tool.

Risk of bias. Green colour = Low risk. Orange colour = some concerns (S). Red colour = high risk of bias (H). Black colour = very high risk of bias (VH).

#### Supplementary Table S7. Full table reporting details about the measure of adherence to self-isolation

| Citation | Definition of self-isolation (duration) | Group that adherence was measured in | Measure of adherence to self-isolation | Definition of adherence to self-isolation | Mode of data collection (length of time between self-isolation and measurement) | Psychometric characteristics of measure, reliability and validity | Acceptability of measure to participants |
|---|---|---|---|---|---|---|---|
| **Ali-Saleh & Obeid 2022 [6]** | Not reported (not reported) | People who had tested positive for COVID-19, had been exposed to a confirmed case, or had returned from abroad | "Asked to quantify the rate of compliance with MoH guidelines on seven components that were created according to MoH guideline" Response options: 5-point scale from 1 (not at all) to 5 (always) Mean scores calculated with higher scores representing higher adherence | Different binary cut offs used for different items Continuous (mean) outcome used for analyses of association | Self-reported – online survey (not reported) | Cronbach's α = 0.80 for seven items | From ns and percentages reported, looks like no missing data for items measuring adherence |
| **Almaghrabi 2021 [7]** | Not reported (not reported) | People who had or who had a family member who had been abroad during the last 2 weeks | "If yes, stayed at home after return back" Response options: not reported | Not reported | Self-reported – online survey (travel had to be in previous 2 weeks) | Not reported | From ns and percentages reported, looks like 13% missing data for item measuring adherence |
| **Almayahi & Al Lamki 2022 [8]** | "Self-isolate" (10 days) | People who had tested positive for COVID-19 (PCR) | "Explored the conditions of health isolation, including duration, place, conception, medical service, challenges and compliance with isolation protocols" Response options: not reported | Not reported | Self-reported – online survey (not reported, minimum of 9 days, maximum not reported) | Not reported | From ns and percentages reported, looks like no missing data for items measuring adherence |
| **Aslaner et al** | Not reported (not | People who | Not reported | Not "did not follow | Self-reported – | Not reported | From ns and |

| | | | | | | | |
|---|---|---|---|---|---|---|---|
| **2022 [9]** | reported) | had a positive COVID-19 PCR test and who were not hospitalised, or who tested negative and were in contact with a case | Response options: not reported | the rules and went out of the house while in isolation” | telephone interview (not reported) | | percentages reported, looks like no missing data for items measuring adherence |
| **Bannour et al 2021 [10]** | Not reported (not reported) | People with COVID-19 | Not reported Response options: do not remember, never, sometimes, often, all the time | Not “not respecting the quarantine” | Self-reported – telephone interview using a questionnaire (not reported) | Not reported | From ns and percentages reported, looks like no missing data for items measuring adherence |
| **Bara’a et al 2021 [11]** | Not reported (14 days) | People with symptoms suggesting COVID-19 | Not reported Response options: not reported | “Isolated themselves” | Self-reported – online survey (not reported) | Not reported | From ns and percentages reported, looks like no missing data for item measuring adherence |
| **Carlsen et al 2020 [12]** | Not reported (not reported) | People who reported they had been ill, been tested for COVID-19, had a positive COVID-19 test, or had a confirmed or suspected diagnosis of COVID-19 from a physician in the last 14 days | “Having been in quarantine or self-isolation any time during the previous 14 days” (“not differentiating between the two”) Response options: yes, no | Answering “yes” | Self-reported – online survey (reporting on behaviour in last 14 days) | Not reported | From ns and percentages reported, looks like no missing data for item measuring adherence |

| | | | | | | | |
|---|---|---|---|---|---|---|---|
| **Domenghino et al 2022 [13]** | "Isolation measures (e.g., avoiding contact with others, hygiene measures, wearing a mask) as specified by the Federal Office of Public Health (FOPH)" (not reported) | PCR confirmed cases of SARS-CoV-2 | Not reported<br>Response options: never, very rarely, rarely, occasionally, frequently, almost always, always | "Always" or "almost always" | Self-reported – online survey (1 month and 2 weeks after positive test result) | Not reported | 15% missing data (item asking about number of days in isolation had 25% missing data) |
| **Dowthwaite et al 2021 [14, 15]** | Not reported (not reported) | People who had tested positive for COVID-19; had a household member test positive; had another person close to them test positive; had been asked to self-isolate | "To what extent, if at all, did you follow any advice given to you?"<br>Response options: not at all [1], very little [2], somewhat [3], very much [4], entirely [5], no advice | Not reported | Self-reported – online survey (not reported) | Not reported | No missing data for items measuring adherence |
| **Elaraby et al 2022 [16]** | "Quarantine at home" (not reported) | Full study sample | "Do you stay at home and self-isolate even if you have minor symptoms such as cough, headache, mild fever"<br>Response options: 10-point scale from 1 to 10 (anchors not reported) | Not reported | Self-reported – telephone interview using a semi-structure questionnaire (not reported) | Not reported | Not reported |
| **Enticott et al 2021 [17]** | "Self-quarantine and self-isolation" (14 days) | "Have or you believe you have the virus"<br>"Have been in contact for over 15 min with others who are awaiting test | "Self-quarantining if you have or believe you have the virus"<br>"Self-isolating if you have been in contact for over 15min with others who are awaiting test results"<br>"Self-quarantine at home if you have | Binary cut off, adherent if answered "most of the time"<br>Ordinal outcome used for mixed effects ordinal logistic regressions | Self-reported – online survey (not reported) | Not reported | Outcome items answered by n=395 people in first wave (total n=1005) and n=343 to 416 people in second wave (total n=1051) |

| | | | | | | | |
|---|---|---|---|---|---|---|---|
| | | results" "Have symptoms and are awaiting a COVID-19 result" "Have had close contact with a confirmed case" | symptoms and are awaiting a COVID-19 result" "Self-quarantine if you have had close contact with a confirmed case" Response options: most of the time (4), some of the time (3), seldom (2), never (1) | | | | |
| **Eraso & Hills 2021 [18]** | People with COVID-19 symptoms were required to self-isolate at home and avoid all but essential contact with others (7 days) People who had a household member who had COVID-19 symptoms (14 days) | People who reported COVID-19 symptoms themselves or who had a household member who had COVID-19 symptoms | "After developing symptoms of coronavirus, how many times did you leave your house (going to your garden does not count as leaving your house) for any reason within 7 days of developing symptoms?" Response options: free text entry "After someone you live with developed symptoms of coronavirus, how many times did you have family or friends visit within 14 days of developing symptoms?" Response options: free text entry | Adherent if answered 0 | Self-reported – online survey (not reported) | Not reported | No missing data for outcome variable |
| **Eslamzadeh et al 2022 [19]** | "In accordance with the guidelines provided by Iran's Ministry of Health and Medical Education on how patients with mild to moderate symptoms should | People with diagnosed COVID-19 | 13 items, asking about changing mask every day, using facial mask at home, using personal or well-conditioned bathroom, using trash can with lid, | Continuous measure used | Self-reported – online survey (not reported, but participants were contacted about participating at the end of their | Not reported | Not reported |

| | | | | | | | |
|---|---|---|---|---|---|---|---|
| | quarantine themselves at home" (14 days) | | using separate or well-conditioned room, using personal dishes, maintaining social distance at home, staying in the room and not wandering around, staying at home all the time, not visiting relatives and friends (elsewhere in manuscript, referred to as "stop having guests"), washing hands regularly, using prescribed medications, covering mouth when coughing<br>Response options: 0 (never), 1 (sometimes), 2 (always)<br>Items were summed to give a score ranging from 0 to 26. | | quarantine period) | | |
| **Farooq et al 2021 [20]** | "Quarantine regulations" (not reported) | Paediatric contacts of adult COVID-19 patients | Not reported<br>Response options: not reported | "Compliance with quarantine regulation" | Self-reported – telephone interview (up to four months) | Not reported | Not reported |
| **Foroozanfar et al 2020 [21]** | "Isolation at home includes refusing to visit relatives during illness, avoiding close contact for 10 days and having a single room at home" (10 days) | People with positive PCR test for COVID-19 | "We assessed patients' self-care behaviors by asking questions about their isolation… refusing to visit relatives during illness, avoiding close contact for 10 days and having a single room at home"<br>Response options: not reported | "Observance of all three mentioned conditions" | Self-reported – telephone interview (not reported) | Not reported | 0.6% missing data (n=2) for outcome measure |
| **Fuchs et al 2021 [22]** | "Followed the Centers for Disease Control and | People with positive | "extracted referral sources and | No premature discontinuation of | Data extracted from four administrative | Not reported | No missing data for |

| | | | | | | | |
|---|---|---|---|---|---|---|---|
| | Prevention guidelines to define the isolation period for those with symptomatic and asymptomatic COVID-19 and the duration of quarantine among close contacts” | COVID-19 test, people under investigation and awaiting test results, close contacts to a COVID-19 case | demographic and clinical data, including reasons for leaving the isolation/quarantine hotel early” | isolation/quarantine (premature discontinuation of isolation/quarantine defined as “leaving voluntarily prior to the end of the prescribed isolation period, either against medical advice or declining isolation/quarantine stay after arrival”) | and clinical data sources (discharge reason recorded in medical records at time of person leaving isolation/quarantine hotel) | | outcome measure |
| **Gasperini et al 2022 [23]** | Staying in a separate bedroom, having access to food resources and consuming them alone, washing hands frequently, maintaining social distancing and wearing a face mask when distancing was not possible and during care procedures (not reported) | People who had COVID-19 or who had been in close contact with a COVID-19 case who were in the home care of the Nursing Home Service | Patients and their relatives were asked about “presence of a caregiver (relative or paid assistant) providing care and ensuring proper isolation was explored, presence of personal protective equipment (PPE) and antiseptic and their correct use” Response options: “the general opinion of the home care nurses about the adherence to isolation/quarantine rules was recorded at the end of the questionnaire.” | Not reported | Self-reported – in-person survey conducted by nurse, nurse then made adherence judgement (during isolation / quarantine) | Not reported | No missing data for outcome measure |
| **Hood et al 2022 [24] [case interview]** | “Adherence to isolation and quarantine guidance” | Positive COVID-19 test (PCR or antigen) | Not reported Response options: not reported | Leaving home | Self-reported – mode not reported (“typically occurred 6 days following symptom onset, with symptomatic cases seeking testing 2 to 3 days | Not reported | 23% did not answer question |

| | | | | | | | |
|---|---|---|---|---|---|---|---|
| | | | | | after symptom onset") | | |
| **Hood et al 2022 [24] [survey]** | "Adherence to isolation and quarantine guidance" | Positive COVID-19 test (PCR or antigen) | "Left home for nonmedical reason during isolation period"<br>"Shared room with any uninfected household contact(s)" [Only asked to those who reported living with one or more people who had not been diagnosed with COVID-19]<br>"A household contact left home after known exposure" [Only asked to those who reported living with one or more people who had not been diagnosed with COVID-19]<br>"All household contacts tested for COVID-19" [Only asked to those who reported living with one or more people who had not been diagnosed with COVID-19]<br>Response options: not reported | Leaving home for a nonmedical reason during isolation period | Self-reported – one-third online, remainder telephone interview (one or two days after completing isolation period) | Not reported | Not reported |
| **Kriens 2022 [70]** | "Were told to stay home" ("at least 7 days from the moment when symptoms first arise, or will last until 24 hours after being symptom free if one has been sick for more than 7 days") | People with a positive COVID-19 test | "It can be quite difficult not to head outside if suffering from mild symptoms, no one would be able to do your groceries or you'd like to get some fresh air. Have you, during the isolation period, gone outside to for | Did comply = answered "no"<br>Did not comply = answered "yes"<br>Missing = "I don't know" | Self-reported – online survey (not reported) | Not reported | Not reported |

| | | | | | | | |
|---|---|---|---|---|---|---|---|
| | | | instance go for a walk, do groceries of visit people?" Response options: "Yes, about (number of times)", "No", "I don't know" | | | | |
| **Kyle et al 2021 [25, 26]** | "Self-isolation" ("up to 14 days") | Contacts of COVID-19 cases | "Item that asked respondents to recall the number of times they left the house during their self-isolation period" Response options: 6-point scale from "none" to "every day" | Not reported | Self-reported – telephone survey (not reported) | Not reported | No missing data for outcome variable |
| **Li et al 2021 [27]** | Isolation environment [living in a well-ventilated single room and avoiding living with others; wearing surgical masks; opening window for ventilation; disinfecting furniture in bedroom daily; using separate daily household items and toilet; having a regular caregiver], condition self-monitoring [checking body temperature frequently; visiting hospital if symptoms are severe; caregivers monitor their own temperature and seek medical attention if they develop symptoms], diet and drug guidance [following drug course | People with diagnosed COVID-19 | "Did you keep living in a separate room during home isolation?" "Did you maintain daily ventilation during the isolation?" "Did you use separate toilets during isolation and disinfect them after each use?" "Did you keep your nose covered with tissue when coughing or sneezing during isolation?" "Did you wash your hands and disinfect them after contact with respiratory secretions (saliva, mucus)?" "Did you clean and disinfect your home daily as required?" "Did you maintain temperature monitoring | Answering "fully complied" | Self-reported – survey [mode not reported] (at end of 14-day isolation period) | Not reported | No missing data for items measuring adherence |

| | | | | | | | |
|---|---|---|---|---|---|---|---|
| | prescribed by doctor based on severity of presenting symptoms; increasing drinking water; ensuring energy intake and eating a balanced diet], psychological counselling [caregivers creating a good living environment; caregivers to actively encourage patients; caregivers to prepare books / electronic products; medical staff maintain regular online communication] (14 days) | | every day?"<br>"Did you wear a mask when you were in the public areas at home?"<br>"Did you take medication according to the doctor's advice during your isolation?"<br>"Did you maintain a certain amount of daily activity/ exercise?"<br>"Did you keep regular contact with your doctor?"<br>"Did you ever go out during isolation?"<br>Response options: fully complied, partially complied, not complied | | | | |
| **Lou et al 2020 [28]** | Duration: Continuous home quarantine (7 days)<br>Mask usage: wear masks properly when in close contact with family members<br>Cough etiquette: Cover coughs and sneezes with your elbow, not hands<br>Hand hygiene: wash hands frequently and do not touch mouth, nose, eyes, and other parts of the face without washing hands<br>Distance: keep 1.5 m distance from patients<br>Ventilation: Keep the rooms clean and ventilated: Half an hour | Paediatric patients | Items assessed "each measure" of home quarantine<br>Response options: no adherence (1), occasional adherence (2), basic adherence (3), full adherence (4) | Participants categorised into two groups: high adherence (score ≥ 24), and low adherence (score < 24) | Self-report – telephone interview (8 days after hospital visit) | Not reported | No missing data for any of the items measuring adherence |

| | | | | | | | |
|---|---|---|---|---|---|---|---|
| | in the morning, noon, and night. Keep warm during ventilation<br>Air conditioning: do not use air conditioning, especially central air conditioning<br>Socialising: do not gather with others who are not family<br>Disinfection: swab articles used by the patient with 75% alcohol or soak in hot water (> 56°C) for 30 min<br>Mask disposal: seal used masks in fresh bags and put them in the trash | | | | | | |
| **Martin et al 2021 [29]** | Not leave home except for a limited number of reasons e.g., for an essential medical appointment (10 days) | Contacts of PCR confirmed COVID-19 cases | Item assessed having left home<br>Response options included: "1) to go to the shops for groceries, toiletries or medicine; 2) to go to the shops for other items; 3) to go to work, school or university; 4) to help or provide care for someone; 5) to spend time indoors and in close contact (less than a meter apart and for more than 15 min) with friends or family that they did not live with; 6) to go out for a meal or to an entertainment venue; 7) to take a child to or from school; | Higher risk non-essential contacts. Left home to go to shops, work, help or provide care, out for a mean or to an entertainment venue<br>Lower risk non-essential contacts. Left home to take a child to or from school or to exercise<br>No non-essential contacts. Left home to attend a medical appointment or did not go out | Self-reported – online survey (not reported) | Not reported | No missing data for outcome variable |

| | | | | | | | |
|---|---|---|---|---|---|---|---|
| | | | 8) to exercise; 9) to attend a medical appointment; 10) or for any other reason. Participants could also select 11) 'did not go out for any reason' as an option."<br>Reported for days trying to self-isolate ("included days spent waiting for the test kit to arrive and any days after testing positive") and days with a negative test result | | | | |
| **Mijovic et al 2021 [30]** | "Pre-operative isolation" (June cohort: 14 days. September cohort: 72 hours) | Patients undergoing elective surgery | "1. Did the patient isolate pre-operatively and for how long? 2. What daily activities patients carried out during their isolation period? 3. Who patients self-isolated with and what activities they did together? 4. What activities household members took part in during the isolation period? 5. How patients travelled to and from the hospital for their operation?"<br>Response options: not reported | Green: "isolated with their household bubble or away from household members taking part in other activities. A person from their isolating bubble or hospital transport was used on their operation day"<br>Amber: "isolated at home but left the house for exercise alone or with someone from their house"<br>Red: "did not adequately isolate pre-operatively or travelled to hospital by taxi, public transport or with someone outside of their bubble. | Self-reported [mode not reported] (asked pre-operatively, at end of quarantine period) | Not reported | No missing data for outcome variable |

| | | | | | | | |
|---|---|---|---|---|---|---|---|
| | | | | Patients interacting with others who went shopping or received visitors were included in this category” | | | |
| **Office for National Statistics 2021, 2022 [31-44]** | Not leaving home during self-isolation, except to get or return a test for COVID-19 and not receiving visitors during self-isolation, except for visitors supporting one’s personal care (10 days) | People with a positive COVID-19 test | “Reasons for leaving the house” Response options included: “To go to the shops for groceries toiletries or medicine or other items; To go to their place of work school or university; For medical reasons other than getting or returning a COVID-19 test (e.g., a doctor’s appointment); For outdoor recreation or exercise (e.g., a run a walk to sit in the park); For another reason” | “Did not leave their home during self-isolation, except to get or return a test for coronavirus (COVID-19)” and “did not receive any visitors during self-isolation, except for visitors supporting their personal care” | Self-reported – telephone survey (longest time = 12 days) | Not reported | No missing data for outcome measure |
| **Office for National Statistics 2021 [45-50]** | “Not leaving your home… In addition to staying home, if you are self-isolating you should not receive visitors, unless the purpose of the visit is to provide essential care.” (10 days) | Contacts of COVID-19 cases | “Reasons for leaving the house” Response options included: “Go to the shops for groceries, toiletries or medicine or other items; For outdoor recreation or exercise (e.g., a run, a walk, to sit in the park); Go to your place of work, school or university; Medical reasons other than getting or returning a COVID-19 test (e.g., a doctor’s appointment); | “Did not leave their home during self-isolation, except to get or return a test for coronavirus (COVID-19)” and “did not receive any visitors during self-isolation, except for visitors supporting their personal care” | Self-reported – telephone survey (longest time = 10 days) | Not reported | No missing data for outcome measure |

| | | | | | | | |
|---|---|---|---|---|---|---|---|
| | | | To help or provide care for a vulnerable person; For another reason” | | | | |
| **Office for National Statistics 2021 [51]** | “When you arrive in England from an amber list country or territory, you must travel directly to the place you are staying and not leave…unless they had a job which qualified for travel exemptions or they opted into the voluntary Test to Release scheme” (10 days) | International travellers | “Between arriving at the place you quarantined and the end of your quarantine period, did you leave the house for any reason?” Response options: yes, no, don’t know, prefer not to say | “Did not leave their accommodation for the full quarantine period, except to get or return a coronavirus (COVID-19) test, or to go for emergency medical treatment or hospital attendance” and “did not receive any visitors, except for visitors supporting their personal care” | Self-reported – telephone survey (10 to 14 days after arriving in England) | Not reported | No missing data for outcome measure |
| **Pinheiro et al 2022 [52, 53]** | “Advised to follow strict home quarantine, avoid non essential travel and avoid social contact for the period of the quarantine.” “Persons in quarantine were instructed to use masks, practice hand hygiene measures and stay separated from others” (14 days) | International and domestic travellers | Not reported Response options: not reported | “Strict room quarantine” | Self-reported – semi-structured telephone interview (at end of quarantine period) | Not reported | No missing data for outcome measure |
| **Rifa’i et al 2022 [54]** | “Isolation environment that have good ventilation”. “Self-isolation…can done in private place or medical facility”. “Quarantine is separated from others due to has been exposed with the covid-19 patient and can be released once tested | People with COVID-19, people who are self-isolating for COVID-19, people who have completed self-isolation for COVID-19 | Not reported Response options: not reported | “Stay at home only during self-isolation” | Self-reported – online survey (not reported) | Not reported | No missing data for outcome measure |

| | | | | | | | |
|---|---|---|---|---|---|---|---|
| | negative COVID-19" (not reported) | | | | | | |
| **Ripon et al 2020 [55]** | "instructed to stay in obligatory quarantine and not to go away from the quarantine position…told to wash their hands repeatedly, to dress in masks while they felt any complexity with inhalation, and were told to contact the nearest hospital" (14 days) | PCR confirmed cases of COVID-19 | Not reported<br>Response options: not reported | Not reported | Self-reported – online survey (within 5 days of the end of quarantine) | Not reported | No missing data for outcome measure |
| **Robin et al 2022 [56]** | "Staying at home for the full duration when advised to do so" (not reported) | COVID-19 cases and contacts | "Respondents reported how often they left home for various reasons".<br>Response options included: shop – essential, shop – other, exercise, medical, work, childcare/school, help someone else, meet people, walk dog | Responses dichotomised as "ever" vs "never" left home for each reason<br>"Categorised reports of leaving home during the isolation period into lower and higher-contact outings, defining exercise and dog-walking as lower contact and all other reasons as higher-contact"<br>Formed three groups: no outings, low-contact outings only, high-contact outing | Self-reported – online survey (survey invitations sent on 24 July [phase 1] and 9 October 2020 [phase 2]; recruited from people in system by 12 March 2020) | Not reported | 0.4% (n=1) missing data on one item asking about out-of-home behaviour ("shop – other"). No missing data for other items |
| **Rosca et al 2020 [57]** | "Quarantined" (14 days) | Patients and staff who had come into contact with a confirmed COVID-19 | Not reported<br>Response options: not reported | Not leaving home | Not reported (during quarantine period) | Not reported | No missing data for outcome measure |

| | | | | | | | |
|---|---|---|---|---|---|---|---|
| | | case | | | | | |
| **Rubio et al 2021 [58] [survey]** | Not leaving home (last three days) | People who had a positive PCR test for COVID-19 and were symptomatic | "Over the last 3 days, how many times did you leave the house to go to buy food, to run errands, to go to work, to go to school, to go to a doctor's visit, or other?" Response options: not reported | Not leaving the house for any reason aside from seeing a health care provider | Self-reported survey [mode not reported] (during quarantine, reporting on last three days) | Not reported | No missing data for outcome measure |
| **Rubio et al 2021 [58] [random home visits and calls]** | Not leaving home (last three days) | People who had a positive PCR test for COVID-19 and were symptomatic | Random home visits and calls | Answering the phone or door when the community health worker was outside. Categorised as non-adherent if did not answer the door after 3 attempts or if work, transport, or street noise was noted while talking on the phone | Observer reported (during quarantine) | Not reported | Reported for n=7 participants (NB – 13 households selected) |
| **Ryu et al 2022 [59]** | "Self-quarantine" (14 days) | People with suspected or confirmed COVID-19, people travelling from countries with high COVID-19 infection risk | Violation cases from Korean Ministry of Interior and Safety records | Not having a violation case | "Actively monitored by a mobile application or phone-call twice a day and the public health workers sometimes randomly visit the quarantined place in person" (during quarantine) | Not reported | Not reported, used data for daily number of quarantine cases and daily number of quarantine violations |
| **Sahin et al 2022 [60]** | "If you have cold symptoms, avoid contact with people, especially the elders and those with chronic diseases, and do not go | Whole sample – general population People who had travelled abroad in first | "Do you isolate yourself when you have symptoms of a cold?" Response options: No, sometimes, yes "When you travel | Not reported Not reported | Self-reported – online survey (1 to 17 months) | Not reported | No missing data for outcome measures |

| | | | | | | | |
|---|---|---|---|---|---|---|---|
| | out without wearing a mask”<br>Staying at home on your return from travel (14 days) | period of pandemic (March to May 2020), “mid of the pandemic” (October to December 2020), and currently (May to July 2021) | abroad, did you isolate yourself in the specified time on your return?”<br>Response options: I have not travelled abroad, I did not isolate myself, I isolated for 1-5 days, I isolated for 6-9 days, I isolated for 10 days, I isolated for >10 days | | | | |
| **Scottish Government 2021 [61, 62]** | “Individuals are asked to remain at home or in managed isolation…should not leave their house/accommodation unless this is solely to get or return a COVID-19 test and should not receive visitors from outside their household. Where possible, those self-isolating should try to maintain physical distancing from others within their household who have not been advised to self-isolate” (10 days) | People with a positive PCR test for COVID-19, contacts of COVID-19 cases, international travellers | Perceived adherence. “How well they thought they managed to comply with self-isolation”<br>Response options: all of the time, some of the time, not able at all<br>Behavioural adherence. “Derived measure of compliance based on how soon an individual started to self-isolate after being advised to do so, whether or not they left their home/accommodation during this period and how long they were able to self-isolate for”<br>Response options: not reported | Behavioural adherence. Fully compliant = “a participant who complied with all of these measures”. Partially compliant = “someone who complied with some, but not all” measures. Non-compliant = “participant who did not comply with any” measures | Self-reported – online survey, with option of telephone completion (not reported) | Not reported | Not reported |
| **Senol & Avci 2022 [63]** | “Applying of isolation of the primary case” (not reported) | People who had a positive PCR test for COVID-19 and their household contacts | Primary case. “Applying of isolation of the primary case”<br>Response options: yes, no<br>Household contact. “Applying of quarantine | Not reported | Self-reported survey [mode not reported] (two weeks to three months after positive test) | Not reported | No missing data for outcome measures |

| | | | | | | | |
|---|---|---|---|---|---|---|---|
| | | | to the household contact"<br>Response options: yes, no<br>Secondary case.<br>"Applying of quarantine to the household contact"<br>Response options: yes, no | | | | |
| **Shewasinad Yehualashet et al 2021 [64]** | "Self-isolated" (not reported) | Whole sample – general population | "I have been practicing self-isolation when I have a fever, cough and headache"<br>Response options: never, rarely, sometimes, often, always | Not reported | "Data was collected by 27 health care workers. Google platform was used to collect data" (not reported) | Not reported | No missing data for outcome measures |
| **Smith et al 2020 [65]** | Not leaving home at all if suffering from a new continuous cough or fever (7 days)<br>Not leaving home at all if someone else in the household developed a cough or fever (14 days) | People who reported symptoms in their household in the last 7 days | "In the past twenty-four hours, how many times, if at all, have you left your home for each of the following reasons? (Please type your answers in the boxes below)…<br>To go to the shops for groceries, toiletries or medicine<br>To go to the shops for other items<br>For exercise<br>For a medical purpose excluding going to the shops/ pharmacy for medicine (e.g., an outpatient appointment)<br>To go to work<br>To help someone else (e.g., delivered | Had not left their home in the last 24 hours | Self-reported – online survey (last 24 hours) | Not reported | No missing data for outcome measures |

| | | | | | | | |
|---|---|---|---|---|---|---|---|
| | | | medicine or done their shopping for them)<br>To meet friends or family who do not live with you<br>Other"<br>Response options: free text entry | | | | |
| **Smith et al 2021 [66]** | Duration adjusted self-isolation. Not leaving home for any reason if developed symptoms in last ten days (10 days)<br>Full self-isolation. Not leaving home for any reason if had symptoms in last seven or ten days (since developing symptoms) | Participants who indicated that they had experienced symptoms of covid-19 (high temperature or fever, cough, or loss of sense of smell or taste) in the past seven days, excluding those who had received a negative test since symptoms developed or in the last week | "Participants were asked for what reason, if any, they had left home since the development of symptoms"<br>Response options included: to go to the shops for groceries/pharmacy. to go out to work, to go to the shops for things other than groceries/pharmacy, my symptoms did not persist / were temporary, for a medical need (other than coronavirus), to go for a walk or some other exercise, my symptoms were only mild, my symptoms got better, I don't think it is necessary for me to stay at home, I was too bored, to help or provide care for a vulnerable person, to meet up with friends and/or family, I was too depressed or anxious, I didn't think it | Duration adjusted self-isolation. Reported not leaving home for any reason in the first 10 days after symptoms developed<br>Full self-isolation. Reported not leaving home for any reason since symptoms developed | Self-reported – online survey (last 7 days) | Not reported | No missing data for outcome measures |

| | | | | | | | |
|---|---|---|---|---|---|---|---|
| | | | was that risky, I was too lonely, my symptoms got worse, to get or return a test for coronavirus, I have not left the home at all | | | | |
| **Steens et al 2020 [67]** | "Quarantine of contacts and isolation of ill people" (not reported) | People who reported that, in the previous 7 days, they had been asked to quarantine or isolate themselves or had been in quarantine or isolation for at least one day | Not reported<br>Response options: not reported | "We evaluated adherence by jointly considering whether participants, during the previous 7 days (i) had been asked to quarantine or isolate themselves, and (ii) had been in quarantine or isolation for at least 1 day." | Self-reported – online survey (last 7 days) | "However, several participants reported having received a request to self-isolate or having isolated themselves while not having been tested (11%; n = 194 / 1,704). We are, therefore, uncertain whether the definitions of quarantine and isolation were understood correctly by all and pooled these variables into one variable 'quarantine/isolation' in the analysis." | 97% reported complete information on quarantine / isolation requests and behaviour |
| **Tseng et al 2021 [68]** | "Persons with exposure to COVID-19 to quarantine" (14 days)<br>"Persons with potential symptoms of COVID-19 to quarantine…regardless of test results because results can be falsely negative" (10 days)<br>"Persons with COVID-19 should isolate" (10 days) | Patients advised to quarantine because they had been exposed to COVID-19 or had symptoms of COVID-19 | "Left quarantine or isolation?"<br>Response options: shopping for groceries, work, visit family or friends, other reasons | "Left home" | Self-reported – telephone survey (calls began on 15 April 2020) | Not reported | No missing data for outcome measure |
| **Yang et al 2022 [69]** | "Self-isolation" (not reported) | People with confirmed or | Not reported<br>Response options: not | "top reasons given for non-adherence | Self-reported survey [mode not reported] | Not reported | Not reported |

| | | | | |
|---|---|---|---|---|
| | suspected COVID-19 | reported | were access to food, medication, and fresh air … the challenge of limited space most often cited as the reason for non-adherence” | (not reported) |

#### Supplementary Table S8. Full table of reported rates of, and factors associated with, adherence to self-isolation.

| Citation | Rates of adherence | Overall risk of bias for prevalence | Factors associated with adherence | Overall risk of bias for analyses of association (tool) |
|---|---|---|---|---|
| **Ali-Saleh & Obeid 2022 [6]** | 92.8% stayed indoors*<br>92.0% "always" covered their mouth and nose when coughing<br>93.1% "frequently" washed their hands<br>90.1% avoided contact with other people in the home<br>74.4% generally stayed in a separate room | Very high | Correlations. Adherence associated with: greater personal compliance to Ministry of Health guidance, greater perceived severity of COVID-19, positive attitudes towards adherence. Adherence not associated with: perceived social network adherence, pandemic fatigue, perceived susceptibility to COVID-19, trust in formal institutions, subjective norms about adherence<br>Multiple hierarchical linear regressions (12% variance explained).† Adherence associated with: greater perceived severity of COVID-19. Not associated with: gender, age, religion, pandemic fatigue, susceptibility to COVID-19, trust in formal institutions, attitudes towards adherence, subjective norms about adherence<br>Mediation effect between pandemic fatigue and adherence. Significant indirect effect of: perceived severity of COVID-19. No significant indirect effect of: susceptibility, trust in formal institutions, attitudes towards adherence, subjective norms about adherence | High (ROBINS-E) |
| **Almaghrabi 2021 [7]** | 77.8%* | Very high | - | - |
| **Almayahi & Al Lamki 2022 [8]** | 84.2% "always sleeping in a separated room"<br>90.8% "using personal towels"<br>73.6% "using masks in the presence of other family members"<br>77.3% "putting wastes in double bags"<br>86.8% "using masks when going outdoors for necessary purposes"<br>5.0% "going out during isolation period for socializing"<br>2.1% "receiving visitors in your home"<br>21.4% "going out for important visits only"*<br>11.1% "going out for drive"<br>25.3% "taking care of children" | Some concerns | - | - |

| | | | | |
|---|---|---|---|---|
| **Aslaner et al 2022 [9]** | 97.4%* | Some concerns | - | - |
| **Bannour et al 2021 [10]** | 74.7%* | High | - | - |
| **Bara'a et al 2021 [11]** | 6%* | Very high | - | - |
| **Carlsen et al 2020 [12]** | Ill. 26% men, 33% women*<br>Tested for COVID-19. 53% men, 59% women*<br>Positive COVID-19 test. 79% men, 91% women*<br>Doctor diagnosis. 65% men, 72% women* | Very high | Adjusted logistic regression.<br>Ill.† Adherence associated with: being female, survey wave 11 (vs 12, 13, and 14), living in Innlandet, Vestfold & Telemark, Rogaland, Vestland (vs Viken). Lower adherence associated with: living in Agder (vs Viken).<br>In men.† Adherence associated with: less than high school education (vs 4 years or less of college). Lower adherence associated with: more than 4 years college (vs 4 years or less of college). Not associated with: age.<br>In women.† Adherence associated with: being aged 25 to 34 years (vs 45 to 49 years), being aged 40 to 44 years, less than high school education (vs 4 years or less of college). Lower adherence associated with: being aged 50 to 54 years (vs 45 to 49 years), more than 4 years college (vs 4 years or less of college).<br>Tested for COVID-19.† Adherence associated with: being female, survey wave 12 and 13 (vs 11), living in Innlandet or Rogaland (vs Viken). Lower adherence associated with: living in Oslo, Agder or Vestland (vs Viken).<br>In men.† Adherence associated with: being aged 35 to 39 years (vs 45 to 49 years). Lower adherence associated with: being aged 55 to 59 years (vs 45 to 49 years), being aged 60+ years. Not associated with: education.<br>In women.† Adherence associated with: being aged 25 to 34 years (vs 45 to 49 years), being aged 35 to 39 years, less than high school education (vs 4 years or less of college). Lower adherence associated with: being aged 55 to 59 years (vs 45 to 49 years), being aged 60+ years, more than 4 years college (vs 4 years or less of college).<br>Doctor diagnosis.† Adherence associated with: being | High (ROBINS-E) |

| | | | | |
|---|---|---|---|---|
| | | | female, survey wave 12 and 13 (vs 11), living in Innlandet (vs Viken).<br>In men.† Adherence associated with: missing education (vs 4 years or less of college). Not associated with: age.<br>In women.† Adherence associated with: being aged 40 to 44 years (vs 45 to 49 years). Lower adherence associated with: being aged 60+ years (vs 45 to 49 years). Not associated with: education. | |
| **Domenghino et al 2022 [13]** | 81.5%* | Some concerns | Multivariable ordinal regressions.<br>Before positive test result. Adherence associated with: being male (vs female), being older (40-64 years vs 18 to 39 years; 65+ years vs 18 to 39 years), not living with children (vs living with children). Not associated with: living situation, education, occupation, information status.<br>After positive test result. Adherence associated with: not living with children (vs living with children), higher education (vocational training and specialised baccalaureate, higher technical school or college, university; vs none or mandatory). Not associated with: sex, age, living situation, occupation, information status.<br>Total population.† Adherence associated with: not living with children (vs living with children), not living with pets (vs living with pets). Not associated with: sex, age, education, occupation, information status. | Very high (ROBINS-E) |
| **Dowthwaite et al 2021 [14, 15]** | Of all people who had been asked to self-isolate: mean =3.88 ["very much"], SD = 1.292 (N=207). 10.6% of these stated that they had not received any advice<br>People who reported that they had tested positive for COVID-19: 5.0% not at all, 7.5% very little, 15.0% somewhat, 32.5% very much, 37.5% entirely,* 2.5% no advice<br>People who reported that a close member of their family who is in my household has tested positive for COVID-19: 6.3% not at all, 6.3% very little, 12.5% somewhat, 45.3% very much, 26.6% entirely,* 3.1% no advice<br>People who reported that a nonfamily member of their household has tested positive for COVID-19: 14.9% not | Very high | - | - |

| | | | | |
|---|---|---|---|---|
| | at all, 5.7% very little, 17.2% somewhat, 18.4% very much, 29.9% entirely,* 13.8% no advice<br>People who had been asked to self-isolate: 4.7% not at all, 3.5% very little, 10.6% somewhat, 15.3% very much, 57.6% entirely,* 8.2% no advice<br>Participants with the app who had been notified to self-isolate: 1.5% not at all, 12.1% very little, 10.6% somewhat, 30.3% very much, 45.5% entirely* | | | |
| **Elaraby et al 2022 [16]** | Mean response=7.6 | Very high | - | - |
| **Enticott et al 2021 [17]** | First survey wave. 76.2%.* Split by sex: 71% men, 83% women. Split by age: 69% under 30 years, 77% over 30 years<br>Second survey wave. Self-quarantining if you have or believe you have the virus. 72.7%.* Split by sex: 64% men, 84% women. Split by age: 66% under 30 years, 75% over 30 years.<br>Self-isolating if you have been in contact for over 15min with others who are awaiting test results. 64.1%<br>Self-quarantine at home if you have symptoms and are awaiting a COVID-19 result. 71.2%<br>Self-quarantine if you have had close contact with a confirmed case. 65.4% | Very high | Univariate ordinal regressions. Adherence associated with: being female (vs male), older age. Adherence not associated with: survey wave, index of relative socioeconomic disadvantage, living in a major city, living in a major state, education.<br>Multivariate ordinal regressions.† Adherence associated with: being female (vs male), older age. Adherence not associated with: survey wave, living in a major city. | Very high (ROBINS-E) |
| **Eraso & Hills 2021 [18]** | 62%<br>Of people who experienced COVID-19 symptoms oneself. 75.6%.* "On average" participants infringed rules 1.02 times (SD=2.9)<br>Of people who lived with someone who experienced COVID-19 symptoms. 57.3%.* "On average" participants infringed rules 1.93 times (SD=2.9) | Very high | Binary logistic regression.<br>People who experienced COVID-19 symptoms oneself.† Adherence associated with: higher perceived control over leaving the home, higher perceived control over responsibilities. Not associated with: gender, age, ethnicity, religion, education, employment status, key worker status, deprivation, housing situation, number of people in household, living with a person of vulnerable health status, own vulnerable health, perceived susceptibility to COVID-19, voting in 2019 general election, trust in government, lockdown phase, COVID-19 and social distancing knowledge, social responsibility, self-interest, normative pressure, financial support, community support, support from a special person, support from family, support from friends.<br>People who lived with someone who experienced COVID-19 symptoms.† Adherence associated with: | High (ROBINS-E) |

| | | | | |
|---|---|---|---|---|
| | | | higher perceived susceptibility to COVID-19, lower COVID-19 and social distancing knowledge, higher perceived control over leaving the home, getting community support if needed. Not associated with: gender, age, ethnicity, religion, education, employment status, key worker status, deprivation, housing situation, number of people in household, living with a person of vulnerable health status, own vulnerable health, voting in 2019 general election, trust in government, lockdown phase, social responsibility, self-interest, perceived control over responsibilities, normative pressure, financial support, support from a special person, support from family, support from friends. | |
| **Eslamzadeh et al 2022 [19]** | Mean overall score = 20.11, SD = 6.01. Overall adherence = 77.3%<br>Changing mask every day: mean score = 1.23<br>Using facial mask at home: mean score = 1.28<br>Using personal or well-conditioned bathroom: mean score = 1.34<br>Using trash can with lid: mean score = 1.47<br>Using separate or well-conditioned room: mean score = 1.47<br>Using personal dishes: mean score = 1.49<br>Maintaining social distance at home: mean score = 1.55<br>Staying in the room and not wandering around: mean score = 1.56<br>Staying at home all the time: mean score = 1.68<br>Not visiting relatives and friends / stop having guests: mean score = 1.71<br>Washing hands regularly for 20 seconds: mean score = 1.73<br>Using prescribed medications: mean score = 1.77<br>Covering mouth when coughing: mean score = 1.82 | Very high | Correlations. Adherence associated with: higher cooperativeness, higher self-transcendence, higher persistence, lower harm avoidance. Not associated with: novelty seeking, self-directedness, reward dependency.<br>Multivariable linear regression.† Adherence associated with: being married (vs single), higher cooperativeness, higher persistence. Adherence not associated with: gender, age, education, being employed, harm avoidance, novelty seeking, self-directedness, reward dependency, self-transcendence. | High (ROBINS-E) |
| **Farooq et al 2021 [20]** | 49%* | Very high | χ2.† Adherence associated with: higher parent education, parent(s) work in health profession, symptomatic contact (vs asymptomatic) | Very high (ROBINS-E) |
| **Foroozanfar et al 2020 [21]** | 89.4%* | High | Mann–Whitney U-test, χ2or Fisher's exact test, independent t-test. Adherence associated with: being male, not having a COVID-19 patient in the family. Not associated with: age, number of family members, job, | Very high (ROBINS-E) |

| | | | | |
|---|---|---|---|---|
| | | | years of education, residence, history of chronic disease, smoking cigarettes, smoking hookah, signs and symptoms at admission, using public transport, history of exposure to the patient in the last 14 days, government disinfection, phone tracking by health centre, home visiting.<br>Bivariate logistic regression. Adherence associated with: being male, not having a COVID-19 patient in the family. Not associated with: chills, dry cough, shortness of breath, having muscle pain, phone tracking by health centre. [Results for age, education, job, residence, number of family members, history of chronic disease, smoking cigarettes, smoking hookah, using public transport, history of exposure to the patient in the last 14 days, government disinfection, home visiting not reported.]<br>Multiple logistic regression.† Adherence associated with: dry cough, not having shortness of breath, not having muscle pain, phone tracking by health centre, not having a COVID-19 patient in the family. [Results for gender, chills not reported.] | |
| **Fuchs et al 2021 [22]** | 81%* | Some concerns | Multivariable regression.† Completed stay associated with: older age (60 years or over vs under 40 years), being male, white ethnicity (vs Black), living in a home, apartment, recreational vehicle, or trailer (vs unsheltered homeless), being positive for COVID-19 (vs being a close contact, and vs missing). Not associated with: referral source, Elixhauser medical condition, Elixhauser mental health disorder, Elixhauser substance use disorder, jail stay in past year. | Some concerns (ROBINS-E) |
| **Gasperini et al 2022 [23]** | Adequate home isolation: 75.0% (92.3% in isolation [case]*, 63.2% in quarantine [contact]*) | Very high | $\chi$2.† Not associated with adherence: isolation (case) vs quarantine (contact) | High (ROBINS-E) |
| **Hood et al 2022 [24] [case interview]** | 58% cases reported not leaving home since symptom onset.* 85% cases reported not leaving home since date of testing* | Very high | - | - |
| **Hood et al 2022 [24] [survey]** | 81% did not leave home for nonmedical reason during isolation period*<br>57% did not share room with any uninfected household contact(s)<br>70% did not report that a household contact left home | High | - | - |

| | | | | |
|---|---|---|---|---|
| | after known exposure<br>72% all household contacts tested for COVID-19 | | | |
| **Kriens 2022 [70]** | - | - | Binary logistic regression.† Adherence associated with: being female, being older, lower education, perceiving yourself to be less healthy. Not associated with: perceived social support, knowledge, perceived quality of social relations. | High (ROBINS-E) |
| **Kyle et al 2021 [25, 26]** | 77.8% (95% CI 75.2% to 80.4%) did not leave their house*, 8.2% (95% CI 6.5% to 9.9%) left their home once, 4.1% (95% CI 2.9% to 5.3%) left every day | Some concerns | $\chi^2$. Adherence associated with: not living alone, having children in the household, living in the second and third highest Welsh deprivation quintiles. Not associated with: gender, ethnicity, age, having a household member develop symptoms, having a household member who received the shielding letter, being a key worker, level of income precarity, rurality.<br>Multivariable logistic regression.† Adherence associated with: receiving higher levels of support for self-isolation, higher confidence in ability to self-isolate. Not associated with: gender, age, Welsh index of multiple deprivation, living alone, planning for self-isolation, individual risk perception, believing that self-isolation is effective, understanding why had to self-isolate. | High (ROBINS-E) |
| **Li et al 2021 [27]** | Separate room living: 94.44% fully complied, 5.56% partially complied, 0.00% not complied<br>Ventilation daily: 96.30% fully complied, 3.70% partially complied, 0.00% not complied<br>Separate toilet, disinfected after use: 22.22% fully complied, 9.26% partially complied, 68.52% not complied<br>Cover nose and mouth with tissue when coughing or sneezing: 65.74% fully complied, 16.67% partially complied, 17.59% not complied<br>Disinfect hands after contacting respiratory secretion: 75.00% fully complied, 13.89% partially complied, 11.11% not complied<br>Daily home disinfection: 73.15% fully complied, 24.07% partially complied, 2.78% not complied<br>Monitoring body temperature: 100.00% fully complied, 0.00% partially complied, 0.00% not complied<br>Wear masks during activities in public areas of the family: 93.52% fully complied, 4.63% partially complied, 1.85% not complied | High | - | - |

| | | | | |
|---|---|---|---|---|
| | A constant caregiver: 71.30% fully complied, 23.15% partially complied, 5.56% not complied<br>Take medicine according to the doctor's instruction: 100.00% fully complied, 0.00% partially complied, 0.00% not complied<br>Maintain moderate daily exercise: 47.22% fully complied, 37.96% partially complied, 14.81% not complied<br>Regular contact with doctors: 100.00% fully complied, 0.00% partially complied, 0.00% not complied<br>Prohibition of going out during quarantine: 100.00% fully complied,* 0.00% partially complied, 0.00% not complied | | | |
| **Lou et al 2020 [28]** | Disinfection: 0.00% no, 0.81% occasional, 35.76% basic, 63.43% full. Mean = 1.43, SD = 0.58<br>Distance: 50.51% no, 32.12% occasional, 17.17% basic, 0.20% full. Mean = 1.56, SD = 0.75<br>Hand hygiene: 54.95% no, 21.21% occasional, 23.84% basic, 0.00% full. Mean = 1.65, SD = 0.83<br>Mask usage: 57.17% no, 21.21% occasional, 20.81% basic, 0.81% full. Mean = 1.67, SD = 0.76<br>Cough etiquette: 60.0% no, 24.04% occasional, 15.96% basic, 0.00% full. Mean = 1.69, SD = 0.83<br>Mask disposal: 0.00% no, 4.44% occasional, 89.29% basic, 6.26% full. Mean = 2.59, SD = 1.09<br>Ventilation: 0.00% no, 0.00% occasional, 2.42% basic, 97.58% full. Mean = 3.02, SD = 0.33<br>Socialising: 0.00% no, 0.00% occasional, 80.81% basic, 19.19% full. Mean = 3.19, SD = 0.39<br>Duration: 60.81% no, 34.95% occasional, 4.24% basic, 0.00% full.* Mean = 3.63, SD = 0.50<br>Air conditioning: 25.05% no, 12.93% occasional, 39.60% basic, 22.42% full. Mean = 3.98, SD = 0.15 | High | $\chi 2$. High adherence associated with: female patient gender, older patient age, female caregiver gender, younger caregiver age, one child in the family (vs 2 or more), having an explanation of quarantine measures by nurse. Not associated: caregiver education, place of residence (urban / rural).<br>Multivariate logistic regression.† High adherence associated with: older patient age, caregiver young (vs elderly), having an explanation of quarantine measures by nurse. Not associated with: number of children in the family. | Some concerns (ROBINS-E) |
| **Martin et al 2021 [29]** | Daily testing, at least one positive test result. 11% higher contact activity, 9% lower contact activity, 80% no non-essential activity*<br>Daily testing, only negative test results. 11% higher contact activity, 11% lower contact activity, 82% no non-essential activity*<br>Daily testing not offered. 7% higher contact activity, 9% lower contact activity, 83% no non-essential activity* | High | Higher contact activity. No association with: group.<br>Lower contact activity. No association with: group.<br>No non-essential activity.† No association with: group.<br>Within those offered daily testing. Less non-essential activities associated with: self-isolating (vs receiving a negative test result).<br>Within those offered daily testing. Higher contact activity. No association with: group.<br>Within those offered daily testing. Lower contact | Very high (ROBINS-I) |

| | | | | |
|---|---|---|---|---|
| | | | activity. Less engagement in activity: only negative test results (vs at least one positive test result).<br>Within those offered daily testing. No non-essential activity.† Less engagement in activity: at least one positive test result (vs only negative test result). | |
| **Mijovic et al 2021 [30]** | June cohort. 48.0% red, 36.0% amber, 16.0% green*<br>September cohort. 30.6% red, 16.7% amber, 52.8% green* | Very high | χ2.† Adherence associated with: 72-hour isolation period (vs 14 days). | High (ROBINS-E) |
| **Office for National Statistics 2021, 2022 [31-44]** | 1 to 13 February 2021. 86%*<br>8 to 13 March 2021. 82%*<br>12 to 16 April 2021. 84%*<br>10 to 15 May 2021. 86%*<br>7 to 12 June 2021. 79%*<br>5 to 10 July 2021. 79%*<br>27 September to 2 October 2021. 78%*<br>1 to 6 November 2021. 75%*<br>29 November to 4 December 2021. 74%*<br>4 to 8 January 2022. 79%*<br>7 to 12 February 2022. 80%*<br>28 February to 8 March 2022. 64%*<br>17 to 26 March 2022. 53%*<br>28 March to 2 April 2022. 51%* | Some concerns | 1 to 13 February 2021.† Adherence associated with: fully understanding self-isolation requirements (vs misunderstanding or being unsure). No association with: perceived ease of self-isolation.<br>8 to 13 March 2021.† Adherence associated with: agreeing that it was easy to self-isolate.<br>12 to 16 April 2021.† Once a positive test was received, adherence associated with: first 24 hours (vs remainder of the isolation period). Of those with symptoms before their test, adherence associated with: receiving positive test result (vs period before result was received).<br>10 to 15 May 2021.† Once a positive test was received, adherence associated with: first 24 hours (vs remainder of the isolation period). Of those with symptoms before their test, adherence associated with: receiving positive test result (vs period before result was received). Not associated with: index of multiple deprivation<br>7 to 12 June 2021.† Adherence associated with: earlier survey wave (10 to 15 May 2021 vs 7 to 12 June 2021), understanding self-isolation requirements (vs misunderstanding or being unsure). Not associated with: survey wave (8 to 13 March 2021 vs 7 to 12 June 2021; 12 to 16 April 2021 vs 7 to 12 June 2021). Once a positive test was received, adherence associated with: first 24 hours (vs remainder of the isolation period). Of those with symptoms before their test, adherence associated with: receiving positive test result (vs period before result was received).<br>5 to 10 July 2021.† Adherence associated with: earlier survey wave (10 to 15 May 2021 vs 5 to 10 July 2021), | High (ROBINS-E) |

younger age (18 to 34 years vs 35 to 54 years). Not associated with: survey wave (7 to 12 June 2021 vs 5 to 10 July 2021). Once a positive test was received, adherence associated with: first 24 hours (vs remainder of the isolation period). Of those with symptoms before their test, adherence associated with: receiving positive test result (vs period before result was received).

27 September to 2 October 2021.† Adherence associated with: earlier survey wave (12 to 16 April 2021 vs 27 September to 2 October 2021; 10 to 15 May 2021 vs 27 September to 2 October 2021). Not associated with: survey wave (7 to 12 June 2021 vs 27 September to 2 October 2021; 5 to 10 July 2021 vs 27 September to 2 October 2021). Once a positive test was received, adherence associated with: first 24 hours (vs remainder of the isolation period).

1 to 6 November 2021.† Adherence associated with: earlier survey wave (12 to 16 April 2021 vs 1 to 6 November 2021; 10 to 15 May 2021 vs 1 to 6 November 2021). Not associated with: survey wave (7 to 12 June 2021 vs 1 to 6 November 2021; 5 to 10 July 2021 vs 1 to 6 November 2021; 27 September to 2 October 2021 vs 1 to 6 November 2021). Of those with symptoms before their test, adherence associated with: receiving positive test result (vs period before result was received).

29 November to 4 December 2021.† Not associated with: survey wave (5 to 10 July 2021 vs 29 November to 4 December 2021; 27 September to 2 October 2021 vs 29 November to 4 December 2021; 1 to 6 November 2021 vs 29 November to 4 December 2021). Of those with symptoms before their test, adherence associated with: receiving positive test result (vs period before result was received).

4 to 8 January 2022.† Not associated with: survey wave (27 September to 2 October 2021 vs 4 to 8 January 2022; 1 to 6 November 2021 vs 4 to 8 January 2022; 29 November to 4 December 2021 vs 4 to 8 January 2022), taking an LFT on day 6 and 7, variant, worry about Omicron variant. Of those with symptoms

| | | | | |
|---|---|---|---|---|
| | | | before their test, adherence associated with: receiving positive test result (vs period before result was received).<br>7 to 12 February 2022.† Adherence associated with: fully understanding self-isolation requirements (vs misunderstanding or being unsure). Not associated with: survey wave (4 to 8 January 2022 vs 7 to 12 February 2022). Of those with symptoms before their test, adherence associated with: receiving positive test result (vs period before result was received).<br>28 February to 8 March 2022.† Adherence associated with: self-isolation being a legal requirement.<br>17 to 26 March 2022.† Adherence associated with: earlier survey wave (7 to 12 February 2022 vs 17 to 26 March 2022; 28 February to 8 March 2022 vs 17 to 26 March 2022).<br>28 March to 2 April 2022.† Adherence associated with: earlier survey wave (7 to 12 February 2022 vs 28 March to 2 April 2022; 28 February to 8 March 2022 vs 28 March to 2 April 2022). Adherence not associated with: survey wave (17 to 26 March 2022 vs 28 March to 2 April 2022) | |
| **Office for National Statistics 2021 [45-50]** | 1 to 6 March 2021. 90%*<br>15 to 20 March 2021. 94%*<br>1 to 10 April 2021. 90%*<br>19 to 24 April 2021. 92%*<br>4 to 8 May 2021. 93%*<br>1 to 5 June 2021. 87%*<br>28 June to 3 July 2021. 89%*<br>9 to 16 August 2021. 88%* | Some concerns | 1 to 10 April 2021.† Adherence not associated with: survey wave (1 to 6 March 2021 vs 15 to 20 March 2021 vs 1 to 10 April 2021).<br>19 to 24 April 2021.† Adherence not associated with: survey wave (1 to 10 April 2021 vs 19 to 24 April 2021).<br>4 to 8 May 2021.† Adherence associated with: developing COVID-19 symptoms (vs not), understanding self-isolation guidance (vs misunderstanding or being unsure), agreeing that it was important to follow self-isolation advice, agreeing that it was easy for me to self-isolate, agreeing that coronavirus posed a risk to society, agreeing that coronavirus posed a risk to one or more of my friends and family, agreeing that coronavirus posed a risk to me personally, agreeing that information from the government about coronavirus can be trusted, later survey wave (vs 1 to 10 April 2021). Not associated with: survey wave (4 to 8 May 2021 vs 19 to 24 April | High (ROBINS-E) |

| | | | | |
|---|---|---|---|---|
| | | | 2021).<br>1 to 5 June 2021.† Adherence associated with: earlier survey wave (19 to 24 April 2021 vs 1 to 5 June 2021; 4 to 8 May 2021 vs 1 to 5 June 2021), developing COVID-19 symptoms (vs not), Not associated with: age, sex, index of multiple deprivation, vaccination status, understanding self-isolation guidance.<br>28 June to 3 July 2021.† Adherence associated with: earlier survey wave (4 to 8 May 2021 vs 28 June to 3 July 2021), developing COVID-19 symptoms (vs not), understanding self-isolation guidance (vs misunderstanding or being unsure). Not associated with: survey wave (1 to 5 June 2021 vs 28 June to 3 July 2021), age, sex, index of multiple deprivation, vaccination status.<br>9 to 16 August 2021.† Adherence associated with: earlier survey wave (4 to 8 May 2021 vs 9 to 16 August 2021), understanding self-isolation guidance (vs misunderstanding or being unsure) | |
| **Office for National Statistics 2021 [51]** | 24 to 29 May 2021. 78%*<br>14 to 19 June 2021. 86%*<br>12 to 17 July 2021. 83%*<br>Usually resident in the UK: 84%. Not usually resident in the UK: 79% | Some concerns | Adherence associated with: agreeing that it was important to follow the international arrivals quarantine guidance, agreeing that information from the government about COVID-19 could be trusted, 14 to 19 June 2021 data collection (vs 24 to 29 May 2021 data collection and 12 to 17 July 2021), 12 to 17 July 2021 data collection (vs 24 to 29 May 2021). Not associated with: gender, age, number of vaccine doses, usually resident in the UK, opted into Test to Release, agreeing that it was easy for me to quarantine, agreeing that coronavirus posed a risk to society, agreeing that coronavirus posed a risk to one or more of my friends and family, agreeing that coronavirus posed a risk to me personally.† | High (ROBINS-E) |
| **Pinheiro et al 2022 [52, 53]** | 58.6% "strict room quarantine"*<br>Wearing a mask: 8.5% never, 52.0% occasionally, 39.5% always<br>Hand hygiene measures: 0.0% never, 9.2% occasionally, 90.8% always<br>Physical distancing: 9.9% never, 3.9% occasionally, 86.2% always<br>Spitting in open places: 96.1% never, 1.3% occasionally, | Very high | - | - |

| | | | | |
|---|---|---|---|---|
| | 2.6% always<br>Practising cough etiquette: 2.6% never, 7.2% occasionally, 90.1% always | | | |
| **Rifa'i et al 2022 [54]** | 91.5% stay at home adherence*<br>Compulsory daily activities for self-isolating patient: 89.70% open room window and let the sun or air ventilation coming to the room, 90.91% sunbathe for as long as 10-15 minutes during 10:00 – 13:00, 96.36% wash hands with soap or sanitiser, 46.06% exercise up to 3-5 times a week, 86.06% eat healthy food three times a day separated from family, 60.00% separate the laundry from family, 73.33% cleaning the room every day, 70.30% use a mask when at home, 70.30% wash own cutlery separated from others, 67.88% check body temperature and oxygen saturation day and night, 82.42% sleep in own room separated from family, 57.58% throw mask separated from other garbage | Very high | - | - |
| **Ripon et al 2020 [55]** | 58% "remained inside their house for the time of the quarantine period"*<br>55% wore face mask in presence of a family member<br>67% "monitored their body temperature as recommended"<br>36% "increased washing hands frequently" | Very high | - | - |
| **Robin et al 2022 [56]** | 63% no outings*, 20% low-contact outings only, 16% high-contact outings | High | Risk ratio. No outings associated with: agreeing that following advice would save lives, survey phase 2 (vs 1), not being angry about being asked to self-isolate, having fever, dry cough or difficulty breathing (key COVID-19 symptoms at the time), all non-white ethnic groups, being advised to stay in room (vs stay inside), being able to get groceries delivered if tried, not thinking would lose touch with family/friends, physical health not worsening. Not associated with: having outside help, having a room to live/sleep in, age, having help with pets if any, having outside space, mental health worsening, being very ill and needing care from family, having a pet at home, contact from Public Health England by text (vs email or phone), thinking could pass on virus if went out.†<br>Low-contact outings associated with: not agreeing that following advice would save lives, not having fever, dry cough or difficulty breathing (key COVID-19 symptoms | Very high (ROBINS-E) |

| | | | | |
|---|---|---|---|---|
| | | | at the time), not identifying as non-white ethnic groups, not being advised to stay in room (vs stay inside), older age, not having help with pets if any. Not associated with: survey phase, being angry about being asked to self-isolate, having outside help, having a room to live/sleep in, having outside space, trying but being unable to get groceries delivered, mental health worsening, thinking would lose touch with family/friends, being very ill and needing care from family, physical health worsening, having a pet at home, contact from Public Health England by text (vs email or phone), thinking could pass on virus if went out.<br>High-contact outings associated with: not agreeing that following advice would save lives, being angry about being asked to self-isolate, not having outside help, trying but being unable to get groceries delivered, mental health worsening, thinking would lose touch with family/friends, being very ill and needing care from family, physical health worsening, not having a pet at home, not having contact from Public Health England by text (vs email or phone). Not associated with: survey phase, having fever, dry cough or difficulty breathing (key COVID-19 symptoms at the time), ethnicity, having a room to live/sleep in, being advised to stay in room (vs stay inside), age, having help with pets if any, having outside space, thinking could pass on virus if went out. | |
| **Rosca et al 2020 [57]** | Patients, of those quarantining at home. 80%*<br>Patients, of those quarantining in the centre. Not reported<br>Staff. Not reported | Very high | - | - |
| **Rubio et al 2021 [58] [survey]** | 91%* | High | - | - |
| **Rubio et al 2021 [58] [random home visits and calls]** | 57%* | High | - | - |
| **Ryu et al 2022 [59]** | Median number of daily self-quarantine violations = 6. Median rate of self-quarantine violations = 1.6 per 10,000 quarantined individuals. | Low | Rate of quarantine violations.† No association with: implementation of a 1-strike out policy and increased penalty amount. | High (ROBINS-E) |

| | | | | |
|---|---|---|---|---|
| | | | Number of quarantine violations in Koreans. No association with: implementation of a 1-strike out policy and increased penalty amount.<br>Number of quarantine violations in foreigners. No association with: implementation of a 1-strike out policy and increased penalty amount. | |
| **Sahin et al 2022 [60]** | When have cold symptoms.<br>First period of pandemic. 3.7% no, 5.1% sometimes, 91.2% yes*<br>Mid of pandemic. 4.0% no, 7.2% sometimes, 88.8% yes*<br>Currently. 5.4% no, 11.3% sometimes, 83.3% yes*<br>Travelling abroad.<br>First period of pandemic. 32% did not isolate, 20% isolated for 1 to 5 days, 2% isolated for 6 to 9 days, 14% isolated for 10 days, 32% isolated for more than 10 days*<br>Mid of pandemic. 34.4% did not isolate, 14.1% isolated for 1 to 5 days, 7.8% isolated for 6 to 9 days, 10.9% isolated for 10 days, 32.8% isolated for more than 10 days*<br>Currently. 49.2% did not isolate, 25.4% isolated for 1 to 5 days, 3.4% isolated for 6 to 9 days, 6.8% isolated for 10 days, 15.3% isolated for more than 10 days* | Very high | When have cold symptoms.† Adherence associated with: earlier period in pandemic (first vs mid, first vs current).<br>First period of pandemic.† Adherence associated with: being female. Not associated with: educational status, being a health care worker, population of settlement, having had COVID-19, having family or relatives who had COVID-19, having family or relatives who died due to COVID-19.<br>Mid of pandemic.† Adherence associated with: being female. Not associated with: educational status, being a health care worker, population of settlement, having had COVID-19, having family or relatives who had COVID-19, having family or relatives who died due to COVID-19.<br>Currently.† Adherence associated with: being female. Not associated with: educational status, being a health care worker, population of settlement, having had COVID-19, having family or relatives who had COVID-19, having family or relatives who died due to COVID-19.<br>In those who went abroad.† No association with: period in pandemic.<br>First period of pandemic.† Not associated with: gender, educational status, being a health care worker, population of settlement, having had COVID-19, having family or relatives who had COVID-19, having family or relatives who died due to COVID-19.<br>Mid of pandemic.† Not associated with: gender, educational status, being a health care worker, population of settlement, having had COVID-19, having family or relatives who had COVID-19, having family or relatives who died due to COVID-19.<br>Currently.† Not associated with: gender, educational status, being a health care worker, population of | High (ROBINS-E) |

| | | | | |
|---|---|---|---|---|
| | | | settlement, having had COVID-19, having family or relatives who had COVID-19, having family or relatives who died due to COVID-19. | |
| **Scottish Government 2021 [61, 62]** | COVID-19 cases and contacts together. 74% fully compliant*, 25% partial compliance, 1% non-compliant COVID-19 cases. Wave 1, 80% fully compliant. Wave 2, 72% fully compliant. Wave 3, 74% fully compliant. Contacts. Wave 1, 78% fully compliant. Wave 2, 74% fully compliant. Wave 3, 69% fully compliant. International travellers. 70% fully compliant*, 29% partially compliant, 1% non-compliant | Very high | COVID-19 cases and contacts together.† Adherence associated with: being female, older age, accepting offer of Local Authority support (vs offered but declined), aware of 10 day isolation requirement (vs those overestimating and those underestimating), agreeing that self-isolation is an effective strategy to prevent the spread of COVID-19. Not associated with: COVID-19 cases (vs contacts).<br>COVID-19 cases.† Adherence associated with: earlier survey wave.<br>Contacts.† Adherence associated with: earlier survey wave, living with someone who tested positive (vs being in close contact with a COVID-19 case who did not live in their household).<br>International travellers.† Adherence associated with: managed isolation (vs home), older age, no previous experience of self-isolating, agreeing that self-isolation is an effective strategy to prevent the spread of COVID-19, agreeing that international travel restrictions will help reduce the spread of COVID-19 and new variants of it. Not associated with: survey wave, sex. | Very High (ROBINS-E) |
| **Senol & Avci 2022 [63]** | Primary case. 68.3%*<br>Household contact. 63.8%*<br>Secondary case. 57.5%* | High | - | - |
| **Shewasinad Yehualashet et al 2021 [64]** | 20.1% never, 23.9% rarely, 19.8% sometimes, 19.9% often, 16.4% always* | Some concerns | - | - |
| **Smith et al 2020 [65]** | 24.9%* | High | Univariable logistic regressions. Adherence associated with: being female, not having a child in the household, not working, being separated / divorced / widowed / never married, not having a pet, thinking you have had or currently have COVID-19, reporting self-isolating, worse self-reported general health, not having helped someone outside household, having received help from someone outside household, agreeing that if you leave home and meet other people you could catch COVID-19. Not associated with: age, | High (ROBINS-E) |

being clinically extremely vulnerable (self), education, index of multiple deprivation, social grade, living in a rural or urban area, living alone, household member being clinically extremely vulnerable, home having access to outside space, understanding government measures if no-one in household was symptomatic, understanding government measures if someone in household was symptomatic, worry about COVID-19, perceived social norms, perceptions about impact of lockdown on mental health, perceptions about impact of lockdown on physical health, agreeing that if you completely follow government advice you will lose touch with friends and relatives, agreeing that your friends or family will disapprove if you don't follow government advice, agreeing that you could get in trouble with the police if you don't follow government advice, agreeing that it will help save lives if you follow government advice, agreeing that it will help protect the NHS if you follow government advice, agreeing that if you catch COVID-19 you may become very ill, agreeing that if you catch COVID-19 it will have a severe impact on your family's wellbeing, agreeing that if you leave home and meet other people you could pass COVID-19 to someone else, agreeing that it will have a negative impact on how much money you have if you follow government advice, agreeing that because of the current lockdown there is more conflict between people that you live with, agreeing that you will not be able to carry out important religious activities if you follow government advice, agreeing that you are enjoying spending more time at home during the lockdown, not agreeing that you feel a sense of community with other people in your neighbourhood because of COVID-19.

Multivariable adjusted logistic regressions.† Adherence associated with: being female, reporting self-isolating, higher worry about COVID-19, not thinking the current lockdown had made mental health worse, having received help from someone outside household, agreeing that if you leave home and meet other people you could catch COVID-19, agreeing that you feel a

| | | | | |
|---|---|---|---|---|
| | | | sense of community with other people in your neighbourhood because of COVID-19. Not associated with: age, having a child in the household, being clinically extremely vulnerable (self), employment status, education, index of multiple deprivation, social grade, living in a rural or urban area, living alone, marital status, household member being clinically extremely vulnerable, home having access to outside space, pet ownership, thinking you have had or currently have COVID-19, understanding government measures if no-one in household was symptomatic, understanding government measures if someone in household was symptomatic, perceived social norms, perceptions about impact of lockdown on physical health, self-reported general health, having helped someone outside household, agreeing that if you completely follow government advice you will lose touch with friends and relatives, agreeing that your friends or family will disapprove if you don't follow government advice, agreeing that you could get in trouble with the police if you don't follow government advice, agreeing that it will help save lives if you follow government advice, agreeing that it will help protect the NHS if you follow government advice, agreeing that if you catch COVID-19 you may become very ill, agreeing that if you catch COVID-19 it will have a severe impact on your family's wellbeing, agreeing that if you leave home and meet other people you could pass COVID-19 to someone else, agreeing that it will have a negative impact on how much money you have if you follow government advice, agreeing that because of the current lockdown there is more conflict between people that you live with, agreeing that you will not be able to carry out important religious activities if you follow government advice, agreeing that you are enjoying spending more time at home during the lockdown. | |
| **Smith et al 2021 [66]** | Duration adjusted self-isolation. 42.5%* (25 to 27 January 2021: 51.8%)<br>Full self-isolation. 20.2% (25 to 27 January 2021: 31.3%) | High | Multivariable logistic regression.<br>Duration adjusted self-isolation.† Not associated with: survey wave, region, gender, age, quadratic age term, having a dependent child in the household, being | High (ROBINS-E) |

| | | | | |
|---|---|---|---|---|
| | | | clinically vulnerable to COVID-19, having a household member who has a chronic illness, being employed, highest earner works in a manual occupation, index of multiple deprivation, education, ethnicity, living alone, working in a key sector, being self-employed, marital status, thinking you had ever had COVID-19, attributing current symptoms to COVID-19, financial hardship.<br>Full self-isolation. Adherence associated with: being female, being older, highest earner not working in a manual occupation, lower education, not working in a key sector, thinking you have not had COVID-19, lower financial hardship. Not associated with: survey wave, region, quadratic age term, having a dependent child in the household, being clinically vulnerable to COVID-19, having a household member who has a chronic illness, employment status, index of multiple deprivation, ethnicity, living alone, being self-employed, marital status, attributing current symptoms to COVID-19. | |
| **Steens et al 2020 [67]** | 42% "at least once" adhered to quarantine/isolation request* | High | Adherence associated with: COVID-19 compatible symptoms (vs those without symptoms), younger age (18 to 29 years vs 30 to 49 years, 18 to 29 years vs 50 to 69 years, 18 to 29 years vs 70 to 89 years), earlier survey wave.† | Very high (ROBINS-E) |
| **Tseng et al 2021 [68]** | 54%* | Some concerns | - | - |
| **Yang et al 2022 [69]** | 56%* | High | - | - |

* Statistic reported in analyses of adherence to self-isolation.

† Results reported in analyses of factors associated with adherence to self-isolation.

#### Supplementary Figure S9. Forest plot showing rates of adherence to self-isolation in individual studies investigating different reasons for self-isolating by risk of bias rating. Error bars are 95% confidence intervals. All studies were rated as very high risk of bias.

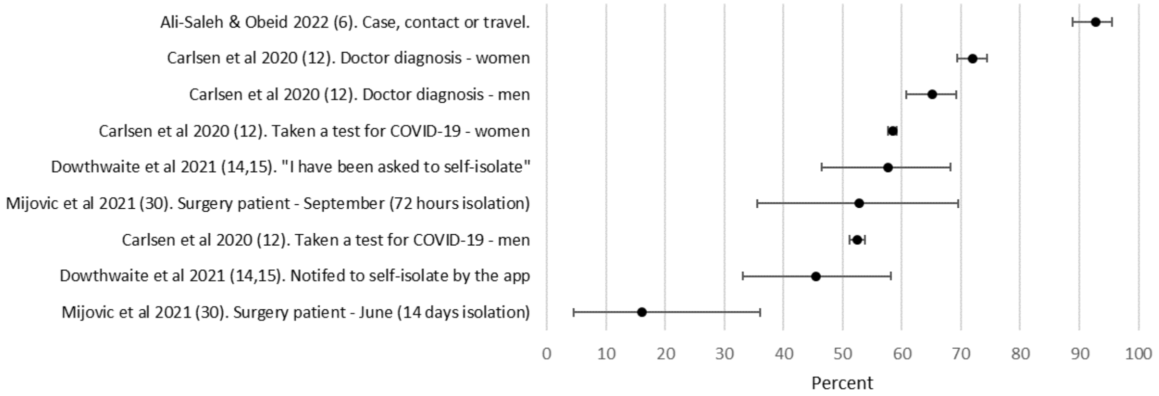

Individual studies investigated other reasons for self-isolation, with adherence ranging from 16% in surgery patients required to self-isolate for 14 days, [30] to 92.8% in COVID-19 cases, contacts, and people returning from travel (all analysed together). [6] All studies were rated as high risk of bias and most had wide confidence intervals.

### Supplement S10. Full narrative description of factors associated with adherence to self-isolation

#### Personal characteristics

Overall, there was little evidence that socio-demographic factors were associated with adherence to self-isolation (Table 4, see Supplementary Table S8 for full results). There was mixed evidence for an association between age and adherence to self-isolation, with five analyses finding an association between older age and adherence (one some concerns of bias, one high risk of bias, three very high risk of bias), [17, 22, 61, 62, 70] three analyses finding an association between younger age and adherence (two high risk of bias, one very high risk of bias), [12, 31-44, 67] and 12 analyses finding no evidence for an association (ten high risk of bias, two very high risk of bias). [6, 12, 13, 18, 19, 25, 26, 45-51, 56, 65, 66] There was weak evidence for an association with female gender and adherence to self-isolation, with six of 18 analyses finding an association (four high risk, two very high risk). [12, 60, 70] [17, 61, 62, 65] Eleven analyses found no evidence for an association (nine high risk, two very high risk). [6, 13, 18, 19, 25, 26, 45-51, 60-62, 66] One analysis (some concerns of bias) found an association between being male and higher adherence. [22] This study investigated people experiencing homelessness or unstable housing and included a low percentage of females (24%) and therefore results are unlikely to be generalisable to the general population. There was mixed evidence for timepoint in the pandemic and adherence to self-isolation, with five analyses finding associations between earlier timepoint and increased adherence (two high risk of bias, three very high risk of bias), [12, 60-62, 67], three studies finding mixed evidence (all high risk of bias) [31-51], and four analyses finding no evidence for an association (two high risk of bias, two very high risk of bias). [17, 60-62, 66] There was weak evidence that lower education was associated with adherence to self-isolation with three analyses finding an association (all high risk of bias), [12, 70] eight analyses finding no evidence for an association (seven high risk of bias, one very high risk of bias), [13, 18, 19, 60, 65, 66] and one analysis finding an association between adherence and higher education (very high risk of bias). [20] There was mixed evidence for an association and adherence to self-isolation with ethnicity, with one study (some concerns of bias) finding an higher adherence in people with white ethnicity (vs Black), [22] and another study (very high risk of bias) finding higher adherence in people from all non-white ethnic groups (vs white). [56] Three studies found no evidence for an effect (all high risk of bias). [18, 66] As the study at some concerns of bias investigated people experiencing homelessness, these results are unlikely to be generalisable.

There was little evidence (supported by one analysis) for adherence to self-isolation and: working in a key sector (including healthcare; association found by one very high risk of bias analysis, no evidence for an association in four high risk analyses); being partnered (association found by one high risk of bias analysis, no evidence for an association in two high risk analyses); not having a dependent child in the household (association found by one very high risk of bias analysis, no evidence for an association in two high risk analyses); and not having a pet (association found by one very high risk of bias analysis, no evidence for an association in one high risk and one very high risk of bias analysis; Table 4). There was no evidence for associations between adherence to self-isolation and living in a more deprived area, employment status, living alone, living in an urban area, religion, social grade, housing situation, having access to outdoor space, or lockdown phase (Table 4).

#### Clinical characteristics

There was little evidence that adherence to self-isolation was negatively impacted by: mental health worsening (association found by one high risk of bias analysis, no evidence for an association in one analysis with some concerns of bias and one analysis and very high risk of bias); better self-reported general health (association found by one high risk of bias analysis, no evidence for an association in one high risk analysis); and physical health worsening (association found by one very high risk of bias analysis, no evidence for an association in one high risk analysis; Table 4). There was no evidence for associations between adherence to self-isolation and being clinically vulnerable to COVID-19, having a household member who had a chronic illness, and number of COVID-19 vaccine doses received (Table 4).

#### COVID-19 infection

There was good evidence that having COVID-19 symptoms was associated with adherence to self-isolation, with four analyses (one high risk of bias, three very high risk of bias) finding an association. [20, 45-50, 56, 67] Another very high risk of bias analysis found mixed evidence for an association. [21] There was also good evidence that testing positive for COVID-19 (vs being a contact, testing negative) was associated with adherence to self-isolation (association found by one analysis at some concerns of bias, one high risk analysis, and one very high risk of bias analysis). [22, 29, 31-44] There was little evidence that having friends or relatives who had COVID-19 was associated with adherence to self-isolation (association found in one very high risk analysis, no evidence for an association in two high risk analyses; Table 4).There was no evidence for associations between adherence and thinking you had ever had COVID-19, or having family or relatives who had died from COVID-19 (Table 4).

#### Isolation characteristics

There was little evidence that adherence to self-isolation was associated with it being a legal requirement (one high risk analysis found an association, one high risk analysis did not find an association; Table 4). There was no evidence for associations between adherence and being involved in test-to-release schemes, being in isolation (as a COVID-19 case) or in quarantine (as a contact; Table 4). Where only one analysis investigated a factor, adherence to self-isolation was found to be associated with: shorter self-isolation period [30]; being in the first 24 hours of the isolation period [31-44]; phone tracking by health centre (vs not) [21]; being advised to stay in a separate room (vs staying inside) [56]; isolating in supported isolation (vs at home) [61, 62]; and being in contact with a COVID-19 case that lives in your household (vs outside household). [61, 62] Previous experience of self-isolation was associated with decreased adherence to self-isolation. [61, 62] There was no evidence for associations between adherence to self-isolation and the mode of contact from public health authority (text vs email vs phone), [56] or of having a separate room to live and sleep in (vs not). [56]

#### Psychological factors

The nature of any association between adherence to self-isolation and knowledge about COVID-19, measures and the need to self-isolate was unclear, with two analyses finding that higher knowledge was associated with adherence (one high risk of bias, one very high risk of bias), [31-44, 61, 62] one high risk analysis finding that lower knowledge was associated with adherence, [18] one high risk analysis finding mixed evidence, [45-50] and three high risk analyses finding no evidence for an association. [18, 25, 26, 65] There was no evidence for an association with higher perceived knowledge about measures. [13, 70] Having an explanation of quarantine measures by a nurse was associated with adherence to self-isolation in the one analysis that investigated it (some concerns of bias). [28] There was weak evidence for an association between higher trust in the government and formal institutions and adherence to self-isolation (two high risk analyses found an association, three high risk analyses found no evidence for an association; Table 4). There was weak evidence that believing that self-isolation was effective was associated with adherence to self-isolation with three high risk analyses finding an association and two high risk analyses not finding an association (Table 4). Higher perceived importance of following self-isolation guidance was associated with adherence to self-isolation in two analyses (both high risk of bias; Table 4).

Taken together, there is some evidence that practical support may be associated with adherence to self-isolation. Receiving more support or help was associated with adherence to self-isolation in two high risk analyses (no evidence for an association in one very high risk of bias analysis; Table 4). Receiving community or local authority support was also associated with adherence in two analyses (one high risk of bias, one very high risk of bias; no evidence for an association in one high risk of bias analysis; Table 4). Being able to get groceries delivered if you tried was also associated with adherence to self-isolation in the only analysis that investigated it (very high risk of bias). [56] However, there was no evidence for an association between adherence and receiving support from family, friends or others in two high risk analyses, or with receiving financial support in two high risk analyses (Table 4).

There is weak evidence for an association between adherence to self-isolation and higher perceived susceptibility to COVID-19 with two high risk analyses finding an association and two high risk analyses finding no evidence for an association (Table 4). There was little evidence for an association between adherence to self-isolation and higher perceived risk of COVID-19 (one high risk analysis found an association, no evidence for an association in two high risk analyses), higher worry about COVID-19 (one high risk analysis found an association, no evidence for an association in one high risk analysis), and higher perceived severity of COVID-19 (one high risk analysis found an association, no evidence for an association in one high risk analysis; Table 4). There was no evidence for an association with thinking that you could pass on the virus if you went out (one high risk of bias, one very high risk of bias; Table 4). There was no evidence for an association between adherence to self-isolation and higher subjective norms for self-isolation, and higher perceived social responsibility (Table 4).

There is some evidence for an association between adherence to self-isolation and higher perceived control over leaving the home (two high risk analyses found an association; Table 4). Only one of two high risk analyses found an association between adherence to self-isolation and higher perceived control over responsibilities (Table 4). Higher perceived ease of self-isolation was associated with adherence in one high risk analysis, with one high risk analysis finding mixed evidence for an association, and one high risk analysis finding no evidence for an association (Table 4). Higher confidence in your ability to self-isolate was associated with adherence to self-isolation in the only analysis that investigated it (high risk of bias). [25, 26] However, there was no evidence for an association with planning for self-isolation (high risk of bias). [25, 26] Thinking that you would lose touch with your family or friends was associated with decreased adherence to self-isolation in one analysis (very high risk of bias), but there was no evidence for an association in another analysis (high risk of bias; Table 4). One analysis each investigated the association between adherence to self-isolation and higher perceived social support (high risk of bias [70]) and quality of social relations (high risk of bias [70]), neither of which found evidence for an association.

32. Office for National Statistics. Coronavirus and self-isolation after testing positive in England: 8 March to 13 March 2021. 15 April 2021.

https://www.ons.gov.uk/peoplepopulationandcommunity/healthandsocialcare/healthandwellbeing/bulletins/coronavirusandselfisolationaftertestingpositiveinengland/8to13march2021 (accessed 30 June 2023).

etins/coronavirusandselfisolationaftertestingpositiveinengland/28februaryto8march2022 (accessed 30 June 2023).

53. Pinheiro C. Quarantine of Travellers during the Initial Phase of the COVID-19 Pandemic-Experience from a Rural Setting in Kerala, India. Personal communication.

66. Smith LE, Potts HWW, Amlôt R, Fear NT, Michie S, Rubin GJ. Adherence to the test, trace, and isolate system in the UK: Results from 37 nationally representative surveys. BMJ 2021; 372.

67. Steens A, De Blasio BF, Veneti L, et al. Poor self-reported adherence to COVID-19-related quarantine/isolation requests, Norway, April to July 2020. Eurosurveillance 2020; 25(37).

68. Tseng CW, Roh Y, DeJong C, Kanagusuku LN, Soin KS. Patients' Compliance With Quarantine Requirements for Exposure or Potential Symptoms of COVID-19. Hawaii Journal of Health and Social Welfare 2021; 80(11): 276-82.

69. Yang L, Mitchell D, Clayton F, et al. Self-isolation among discharged emergency department patients with suspected COVID-19. CJEM 2022; 24(1): 97-8.

70. Kriens W. Individual, social and structural factors underlying compliance to COVID-19 related self-isolation: Utrecht University; 2022. https://studenttheses.uu.nl/bitstream/handle/20.500.12932/43026/Kriens_6014690_Master%27s%20Thesis.pdf?sequence=1&isAllowed=y (accessed 5 June 2023).
